# A Mendelian randomization-based drug repurposing pipeline with integrated AI-facilitated prioritization: application to lipid traits and coronary artery disease

**DOI:** 10.64898/2026.02.28.26347341

**Authors:** Sergio Mundo, Monika E. Grabowska, Alyson L. Dickson, Yi Xin, Mojgan Babanejad, Sevim Serley, Alex J. Chen, Bingshan Li, Lang Li, C. Michael Stein, Wei-Qi Wei, QiPing Feng

## Abstract

Drug repurposing can efficiently identify promising therapeutic targets using existing data; however, current approaches have limitations. This calls for high-throughput approaches that combine versatility and rigor. We developed a flexible, high-throughput, Mendelian randomization (MR)-based drug repurposing pipeline with three stages: 1) MR-based protein target identification, 2) MR-based validation, and 3) drug target mapping and AI-assisted drug prioritization. In Stage 1, the pipeline conducts MR analyses to identify proteins with putative causal effects on a specified trait. In Stage 2, targets with significant Stage 1 associations are evaluated using MR for either the same outcome in an external cohort or a related outcome. Targets with consistent directions of association in Stages 1 and 2 are then assessed in Stage 3, which queries a database of druggable targets and then uses large-language models to prioritize repurposing candidates. To demonstrate the utility of this pipeline, we applied it to atherosclerotic cardiovascular disease.

## INTRODUCTION

Drug development is a costly, time-consuming enterprise that frequently ends in failure ([1]-[3]). Drug repurposing offers an efficient opportunity to utilize approved medications for additional conditions; however, historically, repurposing has relied largely upon candidate approaches that yield limited advances ([4], [5]). The advent of high-throughput approaches has expanded the scale of repurposing efforts, but there is also a need to rigorously assess the biological relevance and actionability of any results.

Human genetic research offers a powerful tool for advancing drug development and repurposing efforts ([6], [7]), particularly since naturally occurring genetic variation can mimic the effects of therapeutic modulation on disease pathways. Mendelian randomization (MR) analysis leverages genetic markers as “instruments,” or proxies, for modifiable exposures (e.g., protein levels) to infer causal effects of those exposures on disease outcomes [8]. Since germline genetic variants are randomly allocated at gamete conception and fixed throughout life, MR analyses are less susceptible to confounding factors from environmental variables and reverse causation [9], making MR an attractive approach for identifying and prioritizing drug repurposing targets.

The emergence of large-scale proteomics studies has enabled the identification of protein quantitative trait loci (pQTLs, genetic variants influencing protein abundance) and offers an efficient way to expand the application of human genetics to drug repurposing by assessing the causal relationships between plasma proteins and disease risk factors [10], [11]. The UK Biobank Pharma Proteomics Project (UKB-PPP) is one of the most comprehensive large-scale initiatives for understanding the effects of genetic variation on circulating protein levels [12], currently providing pQTL mapping and GWAS summary statistics for 2,923 proteins. These data provide a unique opportunity for proteome-wide MR that can be applied to identifying repurposing drug candidates with clinical relevance.

To fully exploit these resources for drug repurposing, there is a need for high-throughput, flexible, reproducible pipelines that can systematically: (i) screen the proteome for proteins with putative causal effects on a wide range of clinically relevant traits or diagnoses, (ii) perform rigorous sensitivity analyses and validation to mitigate bias, and (iii) prioritize proteins connected with existing or emerging therapeutics. Such pipelines are critical for moving from ad hoc, target-by-target analyses to a comprehensive “drug repurposing engine” that can rapidly generate, refine, and triage candidate targets using big data.

We introduce a high-throughput, MR-based drug repurposing pipeline (MRDRP) with three steps: 1) MR-based identification, 2) MR-based validation, and 3) drug-target mapping and AI-assisted drug prioritization. This pipeline can be applied to a broad range of clinical characteristics and diagnoses, including binary and continuous traits. Along with this flexibility, it offers rigorous quality control and validation. In Stage 1, after an optional preliminary step to optimize significance thresholds, the pipeline conducts MR analyses to identify proteins as potential drug targets (i.e., exposures) for a trait or condition (i.e., outcomes). The MR analysis includes quality control steps, comprising testing for heterogeneity, horizontal pleiotropy, and Bayesian colocalization. In Stage 2, MR analysis with quality control is conducted with the proteins yielding significant results from Stage 1 (exposures) for either the same (external cohort only) or a related outcome (e.g., lipids levels and related cardiovascular outcomes). Drug targets with a consistent and significant direction of association in Stages 1 and 2 are then assessed in Stage 3, which queries a database to identify druggable targets and uses large-language models (LLMs) to prioritize drug candidates based on clinical relevance. To demonstrate the utility and flexibility of this pipeline, we applied it to atherosclerotic cardiovascular disease (ASCVD), given its wide range of associated traits and diagnoses as well as its persistent global burden on human health despite existing therapies ([13], [14]). Specifically, we used cis-acting pQTLs from the UKB-PPP to test the causal effects of circulating proteins on low-density lipoprotein cholesterol (LDL-C) and triglycerides (TGs) in Stage 1. In Stage 2, we further validated the identified proteins for their association with coronary artery disease (CAD), using summary statistics from a large GWAS meta-analysis. In Stage 3, for the proteins consistently demonstrating significance in Stages 1 and 2, we queried the Drug-Gene Interaction Database (DGIdb) for interactions with approved drugs to identify promising drug repurposing opportunities for CAD. We then performed AI-assisted prioritization using three large-language models (LLMS; GPT-5.5 [15], Claude Sonnet 4.6 [16], Gemini 3.1 Preview [17]) to rank drugs based on their potential effectiveness for CAD repurposing to facilitate expert review for potential subsequent clinical trials.

## METHODS

### Study design

The general study design is depicted in **Fig. 1**. (The code to run Stages 1 and 2 of this pipeline is available at https://github.com/smundo/MRDRP. The DGIdb API tool used in Stage 3 can be found here: https://dgidb.org/api).

**Fig. 1.**
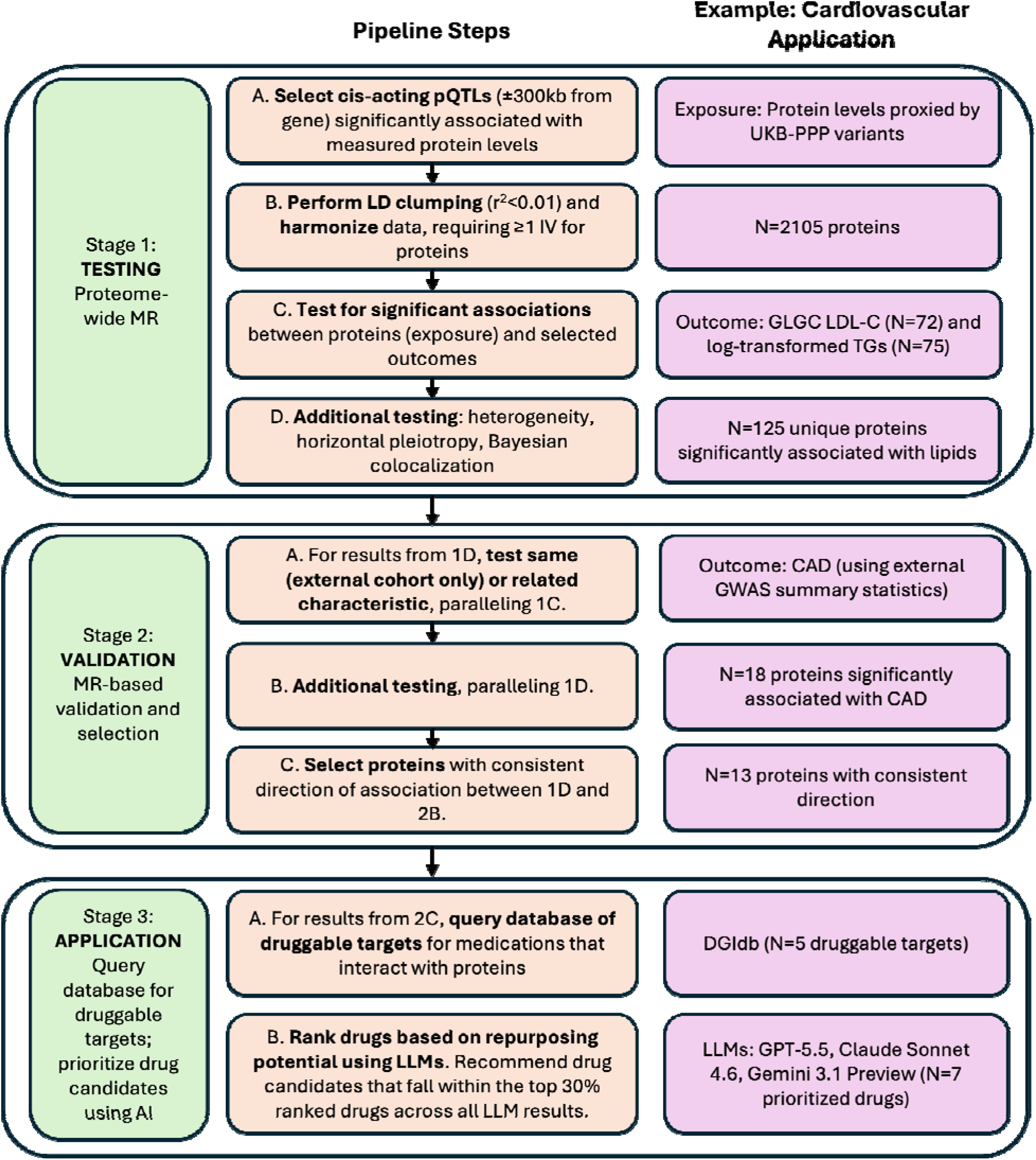
Flowchart of the general study design. The study is divided into 3 stages, with LDL- C and TGs as the outcomes in the first stage and CAD as the outcome in the second. MR=Mendelian randomization; pQTL=protei quantitative trait loci; UKB-PPP=UK Biobank Pharma Proteomics Project; GLGC=Global Lipids Genetics Consortium; LD=linkage disequilibrium; LDL-C=low-density lipoprotein cholesterol; TG=triglyceride; CAD=coronary artery disease; DGIdb=Drug-Gene Interaction Database; AI=artificial intelligence; LLMs=large-language models.

### Data Sources

We tested the pipeline using multiple data sources, including OmicsPred [18], Vanderbilt University Medical Center’s BioVU biobank [19], the UK Biobank Pharma Proteomics Project (UKB-PPP) [12], the Global Lipids Genetics Consortium (GLGC) [20], a large CAD GWAS meta-analysis [21], and the Drug-Gene Interaction Database (DGIdb) [22] (see **Supplementary Methods §1**). OmicsPred and BioVU data were used to optimize the significance threshold for variants in an optional preliminary step before Stage 1 (see **Supplementary Methods §2**). UKB-PPP data were used as the exposure, and GLGC data [after excluding UKB participants; GWAS Catalog: GCST90239655 (LDL-C), GCST90239661 (TGs)] were used as the outcome for the MR analysis in Stage 1 due to their robust and comprehensive GWAS results. A CAD meta-analysis (GWAS Catalog: GCST90132314) was used as outcome data for the MR analysis in Stage 2. The DGIdb was used in Stage 3 to identify currently approved medications that interact with proteins consistently identified in Stages 1 and 2. Given that UKB-PPP and OmicsPred data were derived predominantly from populations of European ancestry, analyses in this study were restricted primarily to individuals of European ancestry (EA) to minimize bias from ancestry mismatch across datasets.

### Stage 1: Proteome-wide Mendelian Randomization (MR)

For each protein in the UKBB PPP dataset (*N*=2,923), we first selected cis-acting pQTLs within 300 kilobase pairs (kb) of any gene that was significantly associated with the protein levels at the optimized significance threshold (*p*<10^-5^; see **Supplementary Methods §2**). We then performed LD clumping at r^2^<0.01 to select independent pQTLs for analysis. We required all instruments to have an *F*-statistic > 10, indicating adequate instrument strength [8]. We also harmonized the exposure and outcome data to ensure that the effect of an instrumental variable (IV) on the exposure and outcome was measured using the same allele in both datasets.

We then utilized the TwoSampleMR [23] package to iterate through individual exposure-outcome pairings to obtain causal estimates between each exposure (i.e., protein levels) and GLGC outcomes (i.e., LDL-C and log-transformed TGs levels). For proteins with only one IV, the Wald ratio was used to calculate the causal estimate, which was interpreted as the change in lipid levels (or log-transformed lipid levels for TGs) per standard deviation increase of circulating protein levels, as proxied by IVs. For proteins with more than one IV, we used the inverse-variance weighted (IVW) method as the primary estimator. Additional MR methods (i.e., simple mode, weighted median, weighted mode, MR-Egger regression) were also calculated for comparison with the main method. When using MR to test qualifying proteins against LDL-C levels and TGs levels (Stage 1C in **Fig. 1**), we considered MR estimates with *p*<0.05 as having a suggestive level of significance, and those with *p*<0.05/2105 (Bonferroni-corrected threshold) as significant.

### Sensitivity analyses for MR

To assess the reliability of the MR estimates, we focused on three main sensitivity analyses: Cochran’s *Q*-test for heterogeneity, MR-Egger for horizontal pleiotropy, and Bayesian colocalization.

*(1) Heterogeneity test.* The heterogeneity test determines whether there is significant evidence of heterogeneity between the IVs based on Cochran’s *Q*-test statistic, which assumes that genetic IVs are uncorrelated. Significant evidence for heterogeneity (denoted as *p_Q_*_-test_<0.05) is found when this statistic is much larger than its degrees of freedom (*N*_IV_ – 1). A significant heterogeneity result suggests that there may be a violation of MR assumptions, potentially invalidating some IVs.

*(2) MR-Egger test for horizontal pleiotropy*. This sensitivity analysis is based on MR-Egger regression, which uses both slope and intercept parameters to estimate a causal effect adjusted for directional pleiotropy. The pleiotropy test specifically uses the statistical significance of the intercept parameter to indicate the likely presence (*p*_MR-Egger_<0.05) of directional horizontal pleiotropy, with a significant result indicating a bias in the IVW estimate. When this was the case, we preferentially reported the MR-Egger estimate.

*(3) Bayesian colocalization*. To control for a potential bias in MR estimates that may arise due to confounding by linkage disequilibrium, we performed Bayesian colocalization using the “coloc” package [24] to assess whether two associated signals (in our Stage 1 example, proteins and lipid levels) were consistent with a shared causal variant. For each protein with a significant MR estimate, we included variants within ±300 kb of the gene. Default parameters were used to perform colocalization, with *p1*=1×10^−4^ (prior probability a variant is associated with protein), *p2*=1×10^−4^ (prior probability a variant is associated with the outcome), and *p12*=1×10^−5^ (prior probability a variant is associated with both protein and the outcome). A posterior probability (PP4) for the main hypothesis (both traits are associated and share a single causal variant) of ≥80% was considered strong evidence of colocalization and therefore supported an MR association with evidence of causal effects between a protein and an outcome when combined with a significant MR estimate.

When one or more of these sensitivity analyses failed for a particular protein with a significant MR estimate, the protein was still considered for the next stage, but we emphasize caution in the interpretation of its results.

### Stage 2: Validation and Selection

#### MR testing and sensitivity analyses

In Stage 2, we tested proteins that yielded significant results in Stage 1 (*N*=125) against CAD risk, using a large CAD GWAS meta-analysis ([21]; *N*=1,165,690 participants; 20,073,070 SNPs). The analytic framework paralleled the methods described in Stage 1. Specifically, the MR estimates represented the log odds change in CAD risk per standard deviation increase of circulating protein levels. We considered MR estimates with *p*<0.05/125 as significant. We then repeated the 3 sensitivity analyses (heterogeneity, horizontal pleiotropy, and Bayesian colocalization), as described above, for the CAD outcome.

#### Consistency checks and selection

To select targets for downstream translation, we additionally confirmed the direction of association between significant results in Stages 1 and 2. For proteins that showed significant associations with both LDL-C and TGs in Stage 1, we further required a consistent direction with both lipids. Only proteins with a consistent direction of effect were assessed further in Stage 3.

### Stage 3: DGIdb Query and AI-based screening

The DGIdb amasses and categorizes drug and gene data from publications, drug databases, and other resources to streamline the search for druggable therapeutic targets, containing over 10,000 genes and 20,000 drugs involved in over 70,000 drug-gene interactions [22]. In Stage 3, to determine the druggability of the resultant proteins from Stage 2, we utilized the DGIdb’s API tool to query for drug-gene interactions for those proteins. We used the direction of the MR estimate from Stage 2 and the DGIdb-reported interaction type to consider those drugs whose mechanism of action on the target protein would imply a reduction of CAD risk as plausible candidates, although we also included drugs whose interactions with the protein are either unclear or unavailable from the DGIdb. After identifying drugs with repurposing potential for CAD prevention, we performed an AI-based screening step to facilitate assessment of whether these drugs should be recommended for further evaluation (e.g., target trial emulation using real-world clinical data). We queried three current state-of-the-art LLMs (GPT-5.5 [15], Claude Sonnet 4.6 [16], Gemini 3.1 Preview [17]) to identify drug repurposing candidates for CAD, using the web search tool in the API to retrieve the most up-to-date information. Specifically, for each drug, we systematically evaluated: (1) the strength of known biological mechanisms in relation to CAD; (2) the long-term safety profile; (3) the availability of potential active comparator drugs; (4) the expected level of confounding introduced by the original indication of the drug; (5) analytic feasibility (if the candidate drug is widely used); (6) availability of completed or ongoing retrospective cohort studies already examining the drug-CAD association; and (7) availability of ongoing or completed clinical trials investigating this drug specifically for the treatment or prevention of CAD. To further validate the potential for repurposing, we also verified the accuracy of the DGIdb-reported indications listed for the drugs. Based on these criteria, candidates were ranked by predicted effectiveness for repurposing using one prompt and validated using a second prompt. We compared the top 30% ranked drugs from each LLM, and prioritized drugs candidates shared by all three in this range. If the LLMs reported an interaction type or effect of a drug on protein levels with a direction that, according to our MR results, was inconsistent with reduced CAD risk, we excluded the drug from the final list of shared top 30% ranked drug candidates.

### Computational tools and software

All statistical analyses were conducted using Python version 3.11.5 and R version 4.4.0. LLM rankings were performed by leveraging API tools for each of 3 LLMs: GPT-5.5 [15], Claude Sonnet 4.6 [16], and Gemini 3.1 Preview [17].

## RESULTS

### Stage 1: Proteome-wide MR analysis to identify candidate protein targets

We applied cis-pQTL selection at *p*<10 and performed proteome-wide MR across all 2,923 proteins in the UKB-PPP. After IV selection and LD clumping, 2,105 proteins were left for MR analysis for both LDL-C and TGs. Using Bonferroni correction (*p*<2.4x10^-5^, 0.05/2105), we found 72 proteins that were significantly associated with LDL-C and 75 with TGs (**Tables 1 & 2**). In total, there were 125 unique significant proteins (22 were associated with both LDL-C and TG levels).

In sensitivity analyses, 30 proteins for LDL-C and 26 proteins for TGs showed evidence of colocalization (PP4 ≥ 0.8). At the nominal threshold (*p_β_* < 0.05), 396 proteins were associated with LDL-C, and 456 proteins were associated with TGs, suggesting the potential for an MR association (see **Supplementary Files 1 & 2**). **Fig. 2** illustrates examples of these findings for PCSK9 and MSR1, including scatter plots depicting different methods, as well as forest plots comparing the IVW estimates with the Wald ratios of individual variants. Scatter plots for the remaining proteins can be found in **Supplementary Folders 1 & 2**, with forest plots found in **Supplementary Folders 3 & 4**.

**Fig. 2:**
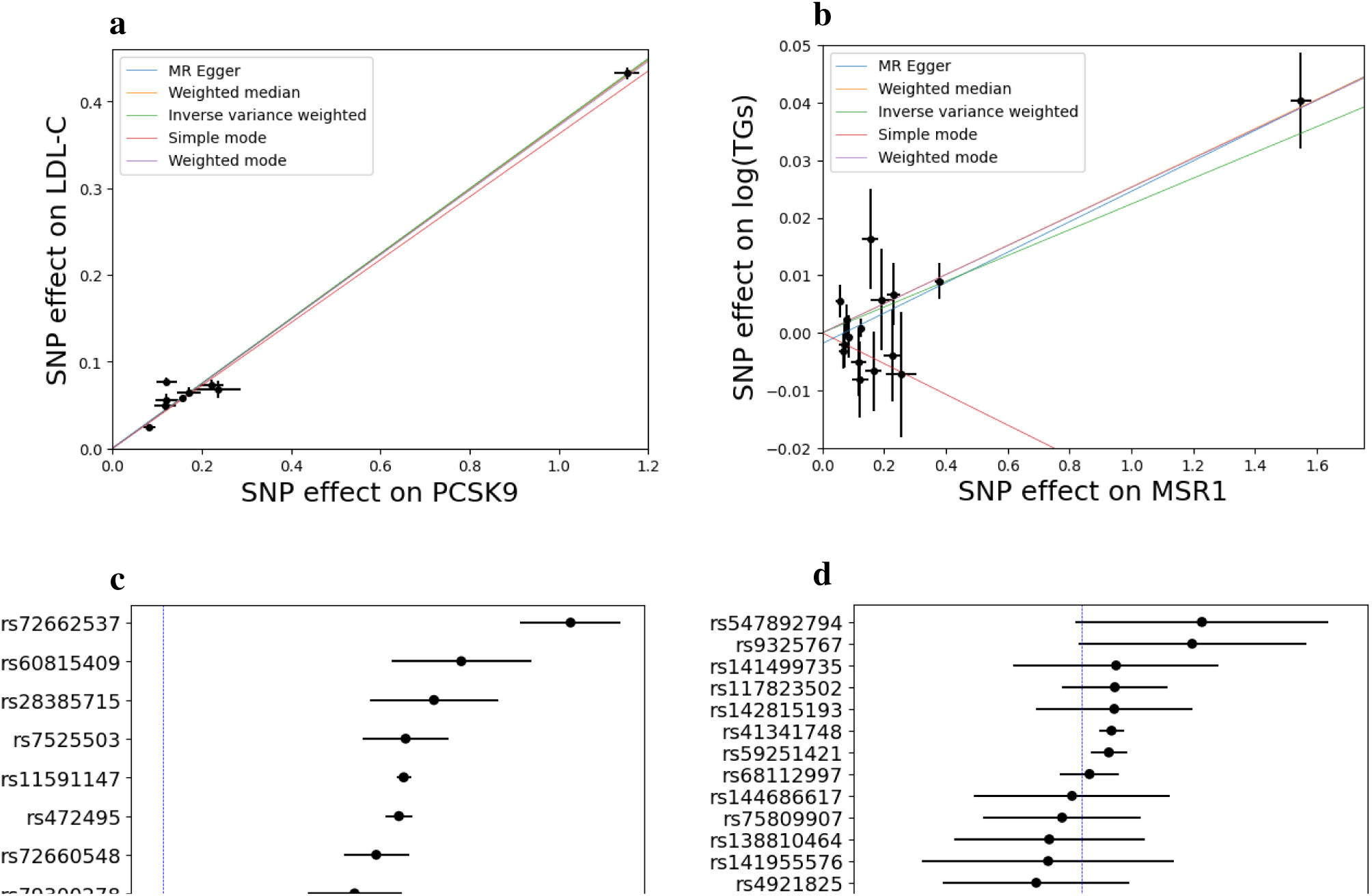
Mendelian randomization (MR) results for PCSK9 levels tested against low-density lipoprotein cholesterol (panels **a** and **c**) and MSR1 levels tested against triglycerides (panels **b** and **d**), using a variant selection threshold of *p*<10^-5^. The first row visualizes the MR analysis methods. The second row shows forest plots with the inverse-variance weighted (IVW) meta-analysis estimate in red and MR estimates for individual IVs. Numerical results for the IVW estimates are shown in Tables 1 and 2. While the simple mode MR method is shown here for MSR1, its estimate is not statistically significant.

**Table 1.**
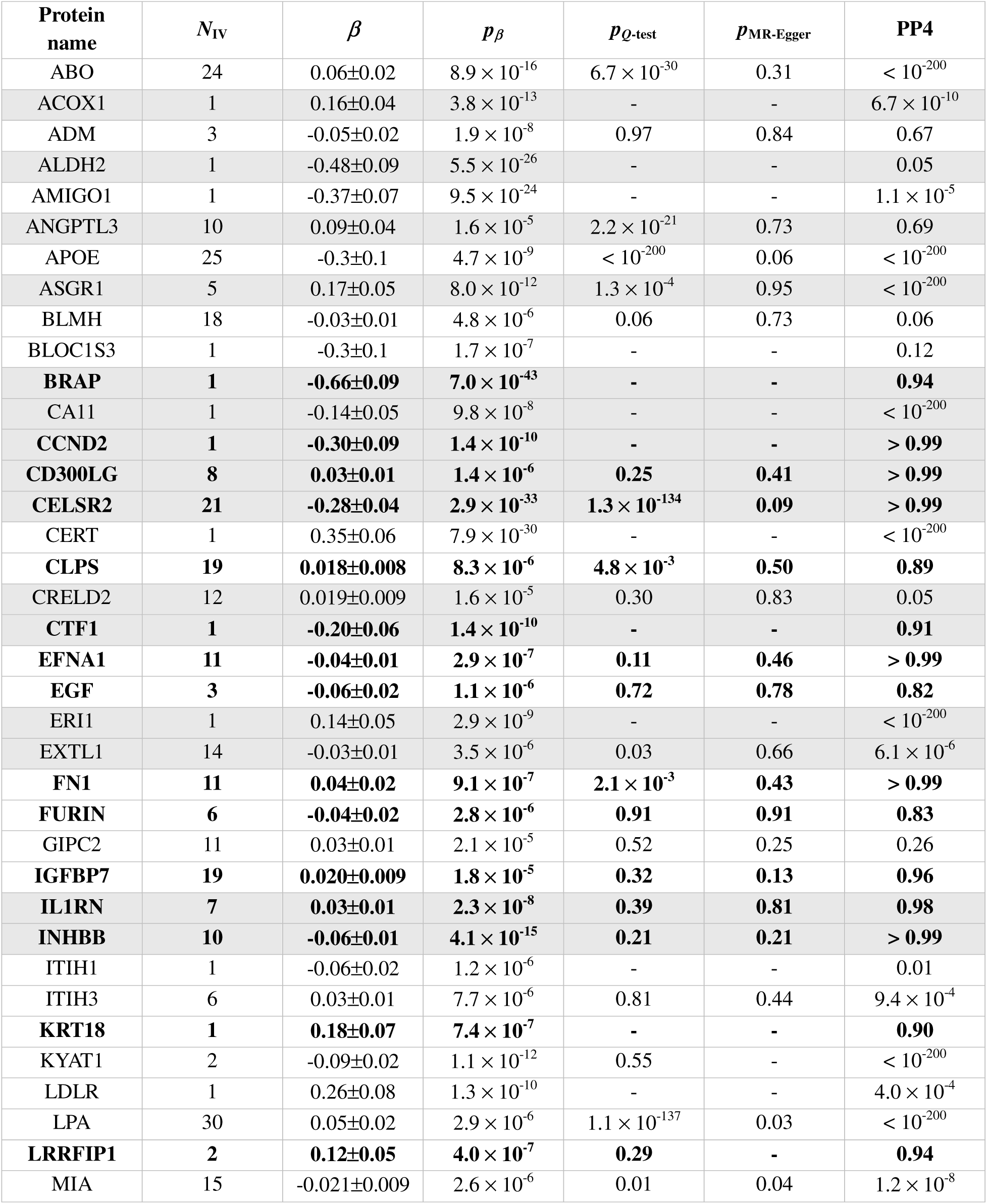
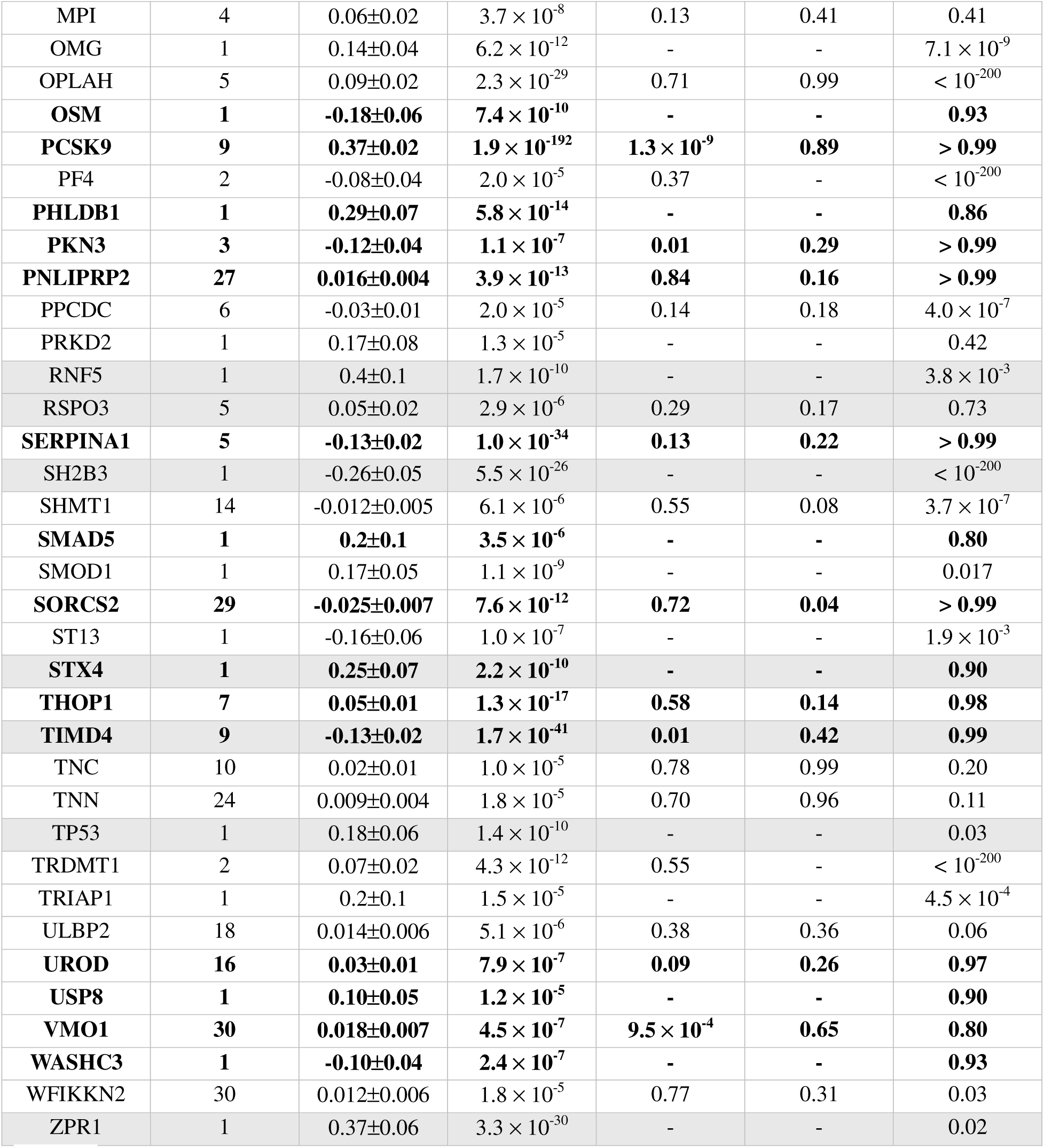
Proteome-wide MR results with LDL-C. Uncertainties are reported at the 95% confidence level. Proteins with only one IV in their analysis do not have results for the heterogeneity or pleiotropy tests because at least two variants are required for these sensitivity analyses. Results with significant evidence for colocalization are highlighted in bold. Proteins that appear in both the LDL-C and TGs results are shown with a gray background.

**Table 2.** Proteome-wide MR results with TGs. Uncertainties are reported at the 95% confidence level. Results with significant evidence for colocalization are highlighted in bold. Proteins that appear in both the LDL-C and TGs results are shown with a gray background. \**The UKB-PPP includes multiple GWAS for TNF, based on different disease/biological process-focused panels in the Olink assay (see* https://olink.com/products/olink-explore-series *for more information). We find 3 significant MR estimates corresponding to 3 of these panels for TNF, but the results are virtually identical within uncertainties. We only report the most significant estimate of the three (corresponding to the Neurology panel), which also features in our CAD results*.

| Protein name | $N_{IV}$ | $\beta$ | $p\beta$ | $p_{Q\text{-test}}$ | $p_{MR\text{-Egger}}$ | PP4 |
| --- | --- | --- | --- | --- | --- | --- |
| AARSD1 | 1 | 0.07±0.03 | $7.3 \times 10^{-7}$ | - | - | $2.8 \times 10^{-6}$ |
| **ACE** | **22** | **0.023±0.008** | **$1.8 \times 10^{-9}$** | **0.39** | **0.28** | **0.86** |
| ACOX1 | 1 | 0.11±0.04 | $9.6 \times 10^{-7}$ | - | - | 0.42 |
| **AKR1C4** | **1** | **0.28±0.06** | **$8.3 \times 10^{-19}$** | - | - | **0.92** |
| ALDH2 | 1 | 0.23±0.09 | $1.9 \times 10^{-7}$ | - | - | 0.15 |
| AMN | 20 | -0.017±0.007 | $1.9 \times 10^{-7}$ | 0.54 | 0.48 | 0.05 |
| **ANGPTL3** | **10** | **0.18±0.07** | **$5.4 \times 10^{-7}$** | **$1.6 \times 10^{-62}$** | **0.41** | **0.94** |
| APOC1 | 14 | 0.31±0.06 | $3.5 \times 10^{-23}$ | $2.1 \times 10^{-84}$ | 0.04 | $< 10^{-200}$ |
| **APOH** | **14** | **0.034±0.007** | **$7.1 \times 10^{-19}$** | **0.23** | **0.25** | **> 0.99** |
| ASGR1 | 5 | 0.05±0.02 | $6.2 \times 10^{-7}$ | 0.30 | 0.24 | $2.6 \times 10^{-4}$ |
| **ATP5IF1** | **1** | **0.07±0.03** | **$6.0 \times 10^{-6}$** | - | - | **0.91** |
| AXL | 12 | 0.05±0.02 | $3.5 \times 10^{-7}$ | 0.03 | 0.51 | 0.02 |
| BOLA1 | 1 | 0.12±0.03 | $1.0 \times 10^{-13}$ | - | - | 0.20 |
| BRAP | 1 | 0.24±0.09 | $4.3 \times 10^{-7}$ | - | - | $5.2 \times 10^{-4}$ |
| CA11 | 1 | -0.14±0.05 | $3.0 \times 10^{-8}$ | - | - | $8.8 \times 10^{-7}$ |
| CASP10 | 2 | 0.04±0.02 | $4.0 \times 10^{-6}$ | 0.84 | - | 0.14 |
| **CCND2** | **1** | **-0.70±0.09** | **$9.9 \times 10^{-53}$** | - | - | **> 0.99** |
| **CD300LG** | **8** | **-0.17±0.02** | **$1.7 \times 10^{-54}$** | **$1.2 \times 10^{-3}$** | **0.42** | **> 0.99** |
| **CELSR2** | **21** | **-0.019±0.009** | **$1.8 \times 10^{-5}$** | **0.10** | **0.38** | **0.99** |
| CFI | 8 | 0.04±0.01 | $8.3 \times 10^{-8}$ | 0.31 | 0.16 | $< 10^{-200}$ |
| CKMT1A | 1 | -0.06±0.03 | $2.5 \times 10^{-6}$ | - | - | $< 10^{-200}$ |
| **CRELD2** | **12** | **0.03±0.01** | **$1.2 \times 10^{-6}$** | **$2.7 \times 10^{-3}$** | **0.40** | **0.95** |
| CTF1 | 1 | -0.22±0.06 | $4.5 \times 10^{-12}$ | - | - | $6.5 \times 10^{-6}$ |
| CXCL16 | 9 | 0.04±0.02 | $1.9 \times 10^{-5}$ | 0.59 | 0.99 | $< 10^{-200}$ |
| DDX58 | 14 | 0.024±0.009 | $5.9 \times 10^{-7}$ | 0.50 | 0.21 | 0.25 |
| DUSP13 | 6 | -0.03±0.01 | $1.8 \times 10^{-6}$ | 0.44 | 0.74 | 0.30 |
| ERI1 | 1 | -0.15±0.05 | $1.6 \times 10^{-10}$ | - | - | $< 10^{-200}$ |
| EXTL1 | 14 | -0.03±0.01 | $5.2 \times 10^{-6}$ | 0.05 | 0.34 | 0.52 |
| **F2** | **1** | **-0.18±0.03** | **$1.7 \times 10^{-25}$** | - | - | **> 0.99** |
| **FABP4** | **1** | **0.11±0.04** | **$8.6 \times 10^{-9}$** | - | - | **0.85** |
| FSHB | 2 | 0.08±0.03 | $8.8 \times 10^{-7}$ | 0.56 | - | $3.7 \times 10^{-3}$ |
| FSTL3 | 8 | -0.06±0.03 | $7.7 \times 10^{-6}$ | 0.15 | 0.65 | 0.02 |
| **GALNT2** | **14** | **-0.12±0.02** | **$2.2 \times 10^{-44}$** | **$6.5 \times 10^{-4}$** | **0.91** | **0.99** |
| GCHFR | 1 | 0.24±0.09 | $6.1 \times 10^{-8}$ | - | - | $6.8 \times 10^{-8}$ |
| **GSTA1** | **8** | **-0.05±0.02** | **$2.3 \times 10^{-10}$** | **0.07** | **0.43** | **0.90** |
| **GSTA3** | **9** | **-0.05±0.01** | **$5.3 \times 10^{-15}$** | **0.30** | **0.21** | **0.98** |
| GSTT2B | 33 | 0.010±0.004 | $9.0 \times 10^{-7}$ | 0.46 | 0.64 | 0.24 |
| HEBP1 | 3 | -0.06±0.02 | $3.5 \times 10^{-7}$ | 0.56 | 0.90 | 0.23 |
| HYOU1 | 4 | 0.05±0.02 | $2.2 \times 10^{-5}$ | 0.14 | 0.24 | 0.72 |
| IGHMBP2 | 1 | 0.17±0.04 | $7.5 \times 10^{-17}$ | - | - | 0.75 |
| **IL1RN** | **7** | **0.04±0.02** | **$5.4 \times 10^{-6}$** | **0.10** | **0.36** | **0.99** |
| INHBB | 10 | -0.05±0.02 | $5.9 \times 10^{-6}$ | $4.5 \times 10^{-3}$ | 0.08 | $< 10^{-200}$ |
| **INHBC** | **21** | **0.033±0.008** | **$3.5 \times 10^{-14}$** | **0.09** | **0.74** | **0.97** |
| INSR | 2 | 0.15±0.05 | $1.4 \times 10^{-7}$ | 0.46 | - | $1.4 \times 10^{-5}$ |
| ITGAL | 1 | 0.15±0.05 | $1.6 \times 10^{-9}$ | - | - | $1.7 \times 10^{-8}$ |
| LDLRAP1 | 2 | 0.09±0.04 | $2.2 \times 10^{-5}$ | 0.70 | - | 0.33 |
| **MEGF9** | **10** | **0.03±0.01** | **$1.4 \times 10^{-8}$** | **0.52** | **0.46** | **0.94** |
| **MERTK** | **7** | **0.04±0.01** | **$7.4 \times 10^{-9}$** | **0.33** | **0.78** | **0.93** |
| MLN | 18 | 0.023±0.008 | $1.9 \times 10^{-8}$ | 0.56 | 0.42 | $1.8 \times 10^{-10}$ |
| **MSR1** | **15** | **0.022±0.009** | **$2.6 \times 10^{-6}$** | **0.20** | **0.26** | **> 0.99** |
| PARP1 | 1 | -0.07±0.03 | $5.3 \times 10^{-7}$ | - | - | $5.1 \times 10^{-3}$ |
| PLEKHO1 | 1 | -0.14±0.06 | $1.6 \times 10^{-6}$ | - | - | $2.1 \times 10^{-10}$ |
| **PLTP** | **10** | **-0.092±0.009** | **$5.5 \times 10^{-95}$** | **0.60** | **0.42** | **> 0.99** |
| PLXNB2 | 28 | 0.019±0.007 | $1.1 \times 10^{-7}$ | 0.17 | 0.69 | $8.1 \times 10^{-5}$ |
| PRKAR2A | 1 | -0.25±0.07 | $2.7 \times 10^{-14}$ | - | - | $< 10^{-200}$ |
| PSMD5 | 1 | -0.18±0.08 | $1.3 \times 10^{-5}$ | - | - | 0.01 |
| **RALY** | **1** | **0.17±0.05** | **$2.9 \times 10^{-13}$** | - | - | **0.81** |
| RNF5 | 1 | 0.8±0.1 | $1.2 \times 10^{-44}$ | - | - | $7.5 \times 10^{-3}$ |
| RSPO3 | 5 | 0.12±0.03 | $1.6 \times 10^{-19}$ | 0.06 | 0.42 | $1.4 \times 10^{-4}$ |
| SEMA6C | 1 | -0.08±0.03 | $1.7 \times 10^{-6}$ | - | - | 0.12 |
| **SERPINF2** | **4** | **-0.08±0.02** | **$4.9 \times 10^{-18}$** | **0.57** | **0.35** | **> 0.99** |
| **SF3B4** | **1** | **0.27±0.06** | **$3.0 \times 10^{-17}$** | - | - | **0.86** |
| SH2B3 | 1 | 0.15±0.05 | $1.3 \times 10^{-7}$ | - | - | $5.8 \times 10^{-5}$ |
| **SHBG** | **15** | **-0.04±0.02** | **$1.6 \times 10^{-5}$** | **$2.7 \times 10^{-4}$** | **0.50** | **> 0.99** |
| SLC4A1 | 1 | 0.15±0.06 | $8.3 \times 10^{-7}$ | - | - | $< 10^{-200}$ |
| SPRED2 | 1 | 0.12±0.04 | $9.2 \times 10^{-8}$ | - | - | $< 10^{-200}$ |
| STX4 | 1 | 0.29±0.08 | $2.6 \times 10^{-13}$ | - | - | $4.2 \times 10^{-3}$ |
| **SULT2A1** | **7** | **0.04±0.01** | **$8.6 \times 10^{-9}$** | **0.25** | **0.25** | **0.89** |
| SV2A | 1 | 0.08±0.02 | $4.5 \times 10^{-16}$ | - | - | 0.75 |
| **TIMD4** | **9** | **-0.14±0.01** | **$8.0 \times 10^{-50}$** | **0.01** | **0.37** | **0.99** |
| TNF* | 5 | -0.14±0.02 | $1.1 \times 10^{-32}$ | 0.61 | 0.75 | $< 10^{-200}$ |
| TP53 | 1 | 0.20±0.03 | $5.0 \times 10^{-13}$ | - | - | $1.8 \times 10^{-3}$ |
| TXNDC15 | 17 | 0.014±0.006 | $2.6 \times 10^{-6}$ | 0.09 | 0.90 | $3.9 \times 10^{-3}$ |
| TYRO3 | 20 | 0.018±0.006 | $1.1 \times 10^{-8}$ | 0.24 | 0.97 | 0.73 |
| ZPR1 | 1 | 2.28±0.06 | $< 10^{-200}$ | - | - | 0.02 |

Additional sensitivity analyses found no evidence for horizontal pleiotropy (non-significant MR-Egger intercept; *p*_MR-Egger_>0.05) and limited heterogeneity for most of the significant associations for which these two tests could be performed (i.e., requiring *N*_IV_ ≥ 2; 29 out of 44 such proteins for LDL-C, and 34 out of 43 such proteins for TGs). Twelve proteins in the LDL-C analysis and eight proteins in the TGs analysis showed no evidence of horizontal pleiotropy but did suggest significant heterogeneity (see **Tables 1 & 2**). This result could be attributed to other forms of pleiotropy, population stratification, or differences in environmental exposures, but does not necessarily invalidate the MR estimate, as consistency across different MR methods was also used to assess reliability (**Supplementary Files 3 & 4**). Only a small number of proteins showed evidence of both horizontal pleiotropy and heterogeneity (*p*_MR-Egger_<0.05 and *p_Q_*_-test_<0.05), including two proteins (LPA and MIA; see **Table 1**) in the LDL-C analysis and one protein (APOC1; see **Table 2**) in the TG analysis, although the MR estimates for LPA across different methods were consistent. Notably, 2 proteins (APOE and CELSR2) showed inconsistent MR estimates between multiple methods and indicated significant heterogeneity.

### Stage 2: MR between significant proteins and coronary artery disease (CAD)

We further tested the 125 unique proteins that were significant in Stage 1 (across LDL-C and TGs analyses; *p_β_* <0.05/2105, see **Tables 1 & 2**) with CAD to assess their plausibility as drug targets for cardiovascular risk reduction. At the nominal threshold (*p*_OR_<0.05), 42 proteins showed associations with CAD (see **Supplementary File 5**). We identified 18 proteins that met Bonferroni significance (*p*_OR_<0.05/125) for MR estimates with CAD; each of these 18 proteins showed consistent significant estimates across different MR methods (see e.g., PCSK9 in **Fig. 3a**, as well as **Supplementary Folder 5** and **Supplementary File 6** for scatter plots and results with different methods, respectively). Forest plots for these proteins can be found in **Supplementary Folder 6**. 6 proteins showed strong evidence of colocalization (i.e., PP4 ≥ 0.8). Only two, APOE and FURIN, showed signs of both horizontal pleiotropy and heterogeneity (*p*_MR-Egger_<0.05 and *p_Q_*_-test_<0.05; see **Table 3**). Thirteen proteins (AARSD1, ABO, ACOX1, APOE, ASGR1, CELSR2, GALNT2, INHBC, LPA, PCSK9, TIMD4, TNF, ZPR1) showed a consistent direction of association between their results for the lipids and CAD analyses; we used these proteins as inputs for the drug database query in Stage 3.

**Fig. 3:**
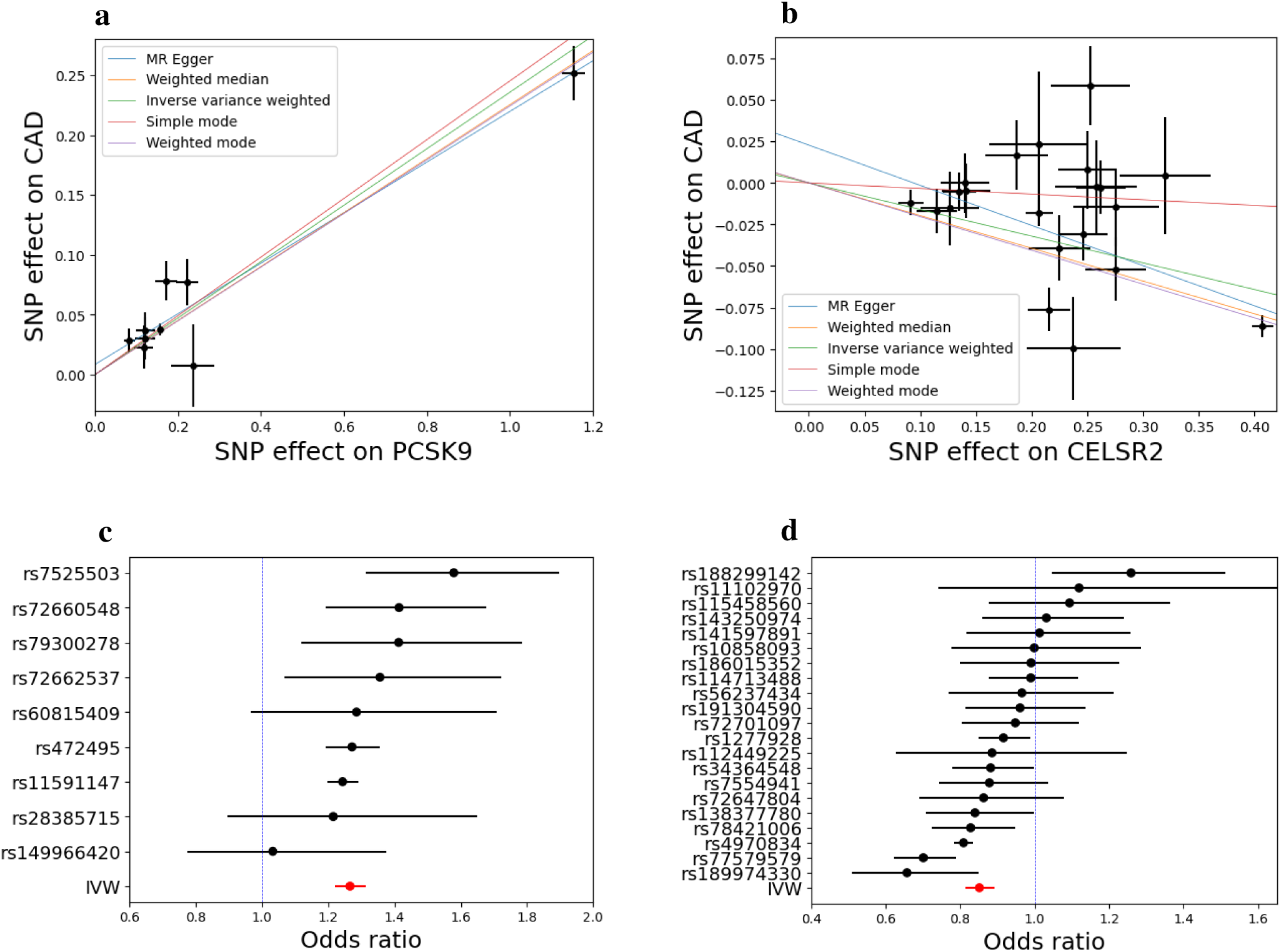
Representative Stage 2 Mendelian randomization (MR) results for coronary artery disease (CAD): PCSK9 levels tested against CAD (panels **a** and **c**) and CELSR2 levels tested against CAD (panels **b** and **d**), using a variant selection threshold of *p*<10^-5^. Panels in the top row display method-specific scatter plots; panels in the bottom row show forest plots for single-variant and inverse-variance weighted (IVW) estimates. Numerical results for the IVW estimates can be found in Table 3. While the simple mode MR method is included for CELSR2, it does not yield a statistically significant MR estimate.

**Table 3.**
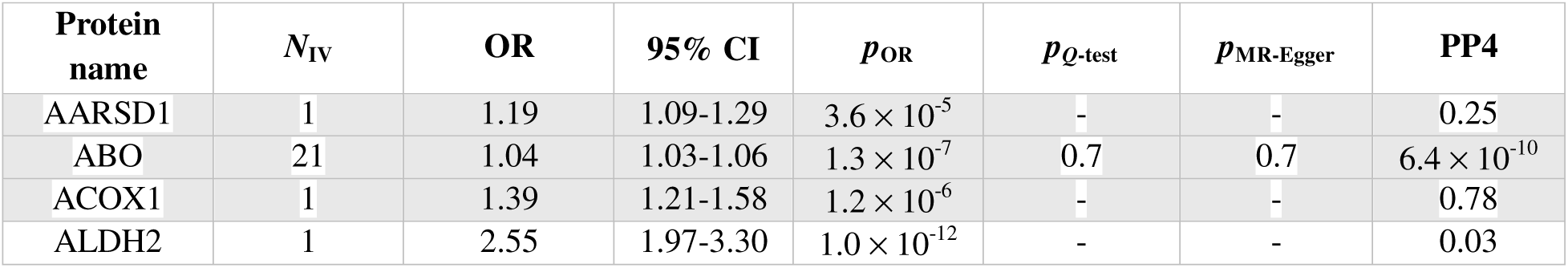
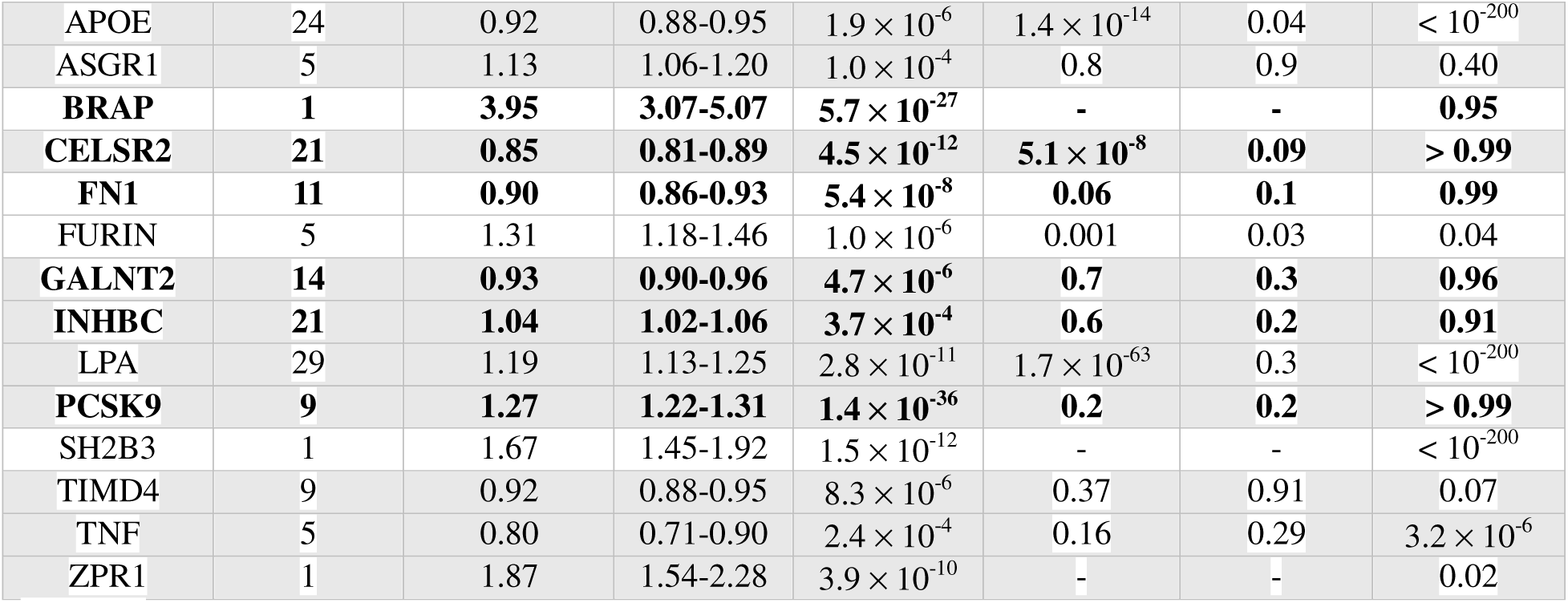
Stage 2 resultant proteins tested against CAD risk. Uncertainties are reported at the 95% confidence level (CI). Results with significant evidence for colocalization are highlighted in bold. Shaded rows correspond to the 13 proteins that showed a consistent direction of association between the results for the lipids and CAD analyses.

### Stage 3: Drug identification for proteins associated with CAD risk

We searched the Drug-Gene Interaction Database (DGIdb) [22] to assess translational potential of 13 proteins that maintained a consistent direction of the MR estimate between the lipids analysis and the CAD analysis. Of these, 6 (APOE, ASGR1, GALNT2, LPA, PCSK9, TNF) were present in the DGIdb as clinically relevant drug targets with drug interactions that could potentially reduce CAD risk, and 5 contained interactions with FDA-approved drugs (interactions with ASGR1 only list unapproved drugs with experimental or investigational status). Associated drug interactions with approved drugs are listed in **Supplementary File 7-Table S1**. After filtering out drugs with low practical feasibility for repurposing for CAD (e.g., tretinoin, which is administered via cream/lotion), as well as those that are already used to manage lipid levels and CAD, we input the remaining drugs into AI-assisted queries using 3 LLMs (GPT-5.5 [15], Claude Sonnet 4.6 [16], and Gemini 3.1 Preview [17]; see **Supplementary File 8** for full prompts), with the requested output designed to facilitate consideration for further testing (e.g., target trial emulation in retrospective cohort studies or clinical trials). Seven drug candidates appeared within the top 30%-ranked drugs across all three LLMs. **Table 4** shows results for two example drug candidates with repurposing potential (see **Supplementary File 9-Table 4** in Supplementary Material folder for full list of 7 drugs). The LLM outputs, while largely consistent, did demonstrate occasional discrepancies due to each model’s particularities, highlighting the need for further validation of LLM-generated results by medical experts. Full LLM results are available in **Supplementary Files 10-12**.

**Table 4.** Representative results of 2 drugs from our AI-assisted screening with CAD repurposing potential (full table available in Supplementary File 9 in Supplementary Material folder).

| **Apixaban** (DGIdb-reported indication: antithrombotic; protein: APOE) |  |  |  |
| --- | --- | --- | --- |
|  | **Claude Sonnet 4.6** | **GPT-5.5** | **Gemini 3.1 Preview** |
| **Known mechanisms in CAD-related pathways** | Apixaban is a direct factor Xa inhibitor that reduces fibrin clot formation on vulnerable atherosclerotic plaques, a central mechanism in acute coronary syndromes. APOE variants influence both lipid metabolism and thrombotic risk. | Direct factor Xa inhibition is mechanistically relevant to coronary thrombosis and ischemic events, although chronic anticoagulation is only beneficial in selected high-thrombotic-risk CAD contexts. | Apixaban is a direct Factor Xa inhibitor. While mapped to APOE in this database, its primary clinical value in CAD is preventing thrombotic complications (MI, stroke) and systemic embolism. |
| **Safety Profile** | Generally well tolerated for chronic use. | Suitable for chronic use when indicated. | Generally better tolerated than warfarin for chronic administration. |
| **Active comparator availability** | Yes—warfarin, rivaroxaban, dabigatran, edoxaban (all anticoagulants with overlapping indications) | High: warfarin, rivaroxaban, dabigatran, edoxaban, low-molecular-weight heparins. | Warfarin, Rivaroxaban, Edoxaban, Dabigatran |
| **Expected bias from indication** | Moderate. Apixaban is prescribed in AF and VTE patients who often have elevated cardiovascular risk, creating confounding by indication. However, the true indication (antithrombotic) is mechanistically directly relevant to CAD, and the bias is manageable with appropriate active comparators. | High: atrial fibrillation and VTE are strongly associated with cardiovascular risk and antithrombotic co-treatment; DGIdb is broadly correct and does not materially change this assessment. | High. Prescribed for atrial fibrillation or VTE, which are significant independent risk factors for MACE and mortality in CAD patients. |
| **Population use** | Widely prescribed | Widely prescribed | Widely prescribed |
| **Retrospective cohort studies** | Multiple retrospective and real-world analyses using Medicare and commercial insurance databases have examined MACE in AF patients with concurrent CAD receiving apixaban | Multiple retrospective comparative-effectiveness studies in AF/VTE populations include MI or MACE outcomes, but these evaluate anticoagulant | Extensive retrospective and registry data (e.g., ARISTOTLE sub-analyses) show its efficacy in preventing cardiovascular events in high-risk patients. |
|  | vs. warfarin, generally demonstrating comparable or reduced MI rates with apixaban. | safety/effectiveness in indicated populations rather than CAD repurposing. |  |
| **Clinical trials** | No large CAD primary prevention trial for apixaban currently identified beyond the anticoagulation context. | No coronary artery disease-specific repurposing trial identified. | AUGUSTUS (NCT02415400) evaluated apixaban in patients with atrial fibrillation and recent acute coronary syndrome or PCI. |
| **Key references** | PMID 21870978, PMID 21780946, PMID 28844192 | PMID 21870978, PMID 21780946 | PMID 21870978, PMID 30883055 |
| **Donepezil** (DGIdb-reported indication: for treatment of Alzheimer's disease; protein: APOE) |  |  |  |
|  | **Claude Sonnet 4.6** | **GPT-5.5** | **Gemini 3.1 Preview** |
| **Known mechanisms in CAD-related pathways** | APOE4 is a shared genetic risk allele for both Alzheimer's disease and coronary artery disease. Donepezil increases synaptic acetylcholine, which activates the cholinergic anti-inflammatory pathway, potentially reducing TNF-alpha and other pro-inflammatory cytokines relevant to atherosclerosis through the vagus nerve. | Cholinesterase inhibition may engage cholinergic anti-inflammatory pathways and has observational cardiovascular signals, but CAD-specific evidence is indirect. | Acetylcholinesterase inhibitor. Observational data suggest these agents are associated with a lower risk of MI and mortality, possibly through a cholinergic anti-inflammatory pathway and improved autonomic balance. |
| **Safety Profile** | Generally well tolerated for chronic use. | Chronic use is common in dementia. | Suitable for chronic use |
| **Active comparator availability** | Yes. Rivastigmine, galantamine, memantine (other Alzheimer's drugs). | High: rivastigmine, galantamine, memantine, and anti-amyloid antibodies for selected patients. | Rivastigmine, Galantamine. |
| **Expected bias from indication** | Moderate. Donepezil users are typically elderly AD patients with advanced age and associated cardiovascular comorbidities. APOE4 genotype links AD and CAD risk biologically. | High: Alzheimer disease, age, frailty, and care patterns strongly confound CAD outcomes. | High. Alzheimer's patients are typically older with significant baseline vascular risk. |
| **Population use** | Widely prescribed among Alzheimer's patients. | Moderate among older adults with dementia. | Widely used in elderly populations. |
| **Retrospective cohort studies** | No large dedicated CAD-specific cohort study identified. | Some dementia cohorts have reported lower MI or mortality among cholinesterase-inhibitor users versus nonusers, but CAD-specific causal inference is weak. | A large Swedish study found a significant reduction in MI and death in AD patients taking cholinesterase inhibitors. |
| **Clinical trials** | No coronary artery disease-specific clinical trials for donepezil identified. | No coronary artery disease-specific clinical trials identified. | No CAD-specific prevention trials identified. |
| **Key references** | 'FDA prescribing information for Aricept (donepezil hydrochloride), Eisai and Pfizer', 'No specific reference identified' | PMID 23616405 | PMID 23739311 |

## DISCUSSION

In this study, we developed a MR-based drug-repurposing pipeline that integrates proteome-wide screening, outcome-based validation, and drug-target mapping. We further demonstrated its utility for identifying druggable targets relevant to lipid lowering and CAD prevention using large-scale GWAS and proteomics data. After optimizing the cis-pQTL IV selection threshold, a proteome-wide MR analysis between all available UKB-PPP proteins and LDL-C/TGs identified 72 and 75 proteins with putative causal effects on LDL-C and TGs, respectively. Thirteen potential drug targets for cardiovascular drug development were identified when testing these proteins against CAD. Our approach successfully replicated established causal associations—in particular, PCSK9, ANGPTL3, and LPA—and revealed previously unreported protein-lipid relationships, offering promising avenues for therapeutic exploration.

In addition to identifying known and novel drug candidates, our study also contributed to methodology. **(1) A flexible, high-throughput pipeline:** MRDRP was constructed with the intent of streamlining the overall drug target identification process via MR. For this reason, this high-throughput pipeline is flexible enough to be used for any exposure-outcome pairings of interest (whether continuous or binary traits) in the context of many diseases, with minor caveats regarding data file organization. The pipeline is also easily re-runnable at any stage forward for updates (e.g., DGIdb output updates) or regular re-assessments. This study served to test the pipeline in the context of cardiovascular risk factors and to provide initial insight into such an analysis. **(2) Translational potential:** We used the DGIdb to assess the druggability of potential drug targets identified in Stages 1 and 2. Of the 13 proteins from our Stage 2 analysis that had a consistent direction of association with lipids, we found 6 listed in DGIdb. These identified proteins in turn yielded drugs with CAD repurposing potential, which we ranked using 3 LLMs based on, among other factors, the long-term safety profiles, the availability of potential active comparator drugs, and analytic feasibility; such results can facilitate clinical expert review and accelerate efficient progression to validation. Notably, although one of the DGIdb-identified proteins, ASGR1, did not show any interactions with approved drugs, one of its associated compounds, arachidonic acid, is an eicosanoid precursor involved in lipid-related pathways ([25]-[27]). There is also evidence suggesting that ASGR1 haploinsufficiency is associated with reduced levels of non-HDL cholesterol and a reduced risk of CAD, which is consistent with our results from Stages 1 and 2 [28]. **(3) Optimization for IV selection**: Rather than defaulting to the genome-wide significance level (*p*<5x10^-8^), we employed a data-driven approach to optimize the *p*-value cutoff for selecting pQTL IVs in the optional preliminary step. We selected *p*<10^-5^ to balance between IV yield and robustness, avoiding significant winner’s curse bias [29].

Our findings suggest that MR, when combined with large-scale pQTL data, can effectively identify proteins with putative causal effects on disease and prioritize drug targets, consistent with previous applications of this approach. For example, Sun *et al*. [30] performed a proteome-wide MR analysis in colorectal cancer and identified 13 potential causal proteins, four of which had already been selected for drug development in other cancers. Other studies have already proposed similar frameworks for cardiovascular drug development (e.g. [6], [31]). For instance, Yang *et al*. [31] introduced a procedure that utilizes genoproteomic data and MR for cardiovascular drug discovery, with an emphasis on translating results from such an analysis into clinical applications. Later stages of their framework implemented an exploration of drug target side effects, as well as an emulation of therapeutic efficacy and safety via a hypothetically randomized controlled trial. Together, these studies support a MR-based proteome scan as a promising strategy for drug discovery and repurposing. However, these studies were not designed to provide the same level of high-throughput scalability or broad generalizability as our pipeline.

The results from this study generally yielded both familiar and lesser-known proteins that may have previously reported associations with lipids and/or ASCVD risk. Evaluating the pipeline’s results for known protein drug targets was designed to help demonstrate its strengths and identify any limitations, as well as confirm established relationships; assessing its results for “new” targets allowed for the consideration of additional drug targets to influence lipids and cardiovascular risk, which could serve as a springboard towards drug discovery. We discuss examples of these targets in the next two subsections.

### Known targets

(1) **PCSK9**. Inhibition of PCSK9, as noted, is an approved and widely used method for lowering LDL-C levels and mitigating the risk of ASCVD. Our pipeline replicated previous results, showing that higher PCSK9 levels may raise circulating LDL-C levels and increase CAD risk (see e.g. [32] and references therein). Our search results in DGIdb included 3 established PCSK9 inhibitors, demonstrating the pipeline’s translational relevance.

(2) **LPA**. Our MR results also replicated a positive association between LPA and cardiovascular traits (namely, LDL-C and CAD). While the LDL-C analysis for LPA indicated heterogeneity and pleiotropy signals, the causal estimates across different MR methods were consistent directionally and within error bars, lending some credibility to the main IVW estimate. The lack of evidence of pleiotropy (i.e., zero average pleiotropic effect) in the CAD analysis further supported the reliability of the estimate in assessing cardiovascular risk, although there was no evidence of colocalization. DGIdb lists 4 approved drugs that interact with LPA, with 3 statins showing indications as lipid-related anti-cholesterolemic agents, as expected. While these medications may modify LPA levels, they do not target LPA directly (see e.g., [33]). As a lipid biomarker that binds cholesterol, LPA is actively being investigated as a potential drug target to reduce residual cardiovascular risk (see [34] and references therein), and the genetic evidence supports continued development.

(3) **LDLR**. The pipeline generated a positive MR estimate between LDLR and LDL-C, despite LDLR’s canonical role in clearing LDL-C. Two factors could impact these results: (i) UKB-PPP quantifies soluble protein in plasma, and LDLR on the surface of hepatocytes, not soluble circulating LDLR (sLDLR), is the main mediator of plasma LDL-C clearance; sLDLR levels do not necessarily proxy hepatocyte-surface LDLR levels. In addition, sLDLR can track processes (e.g., receptor shedding/inflammation) that positively associate with elevated LDL-C levels, as reported previously ([35]-[38]). (ii) Furthermore, the MR estimate relies on a single instrument, which may not be enough to accurately predict this relationship, and there is no evidence for colocalization. As more data becomes available, we will conduct a multi-IV analysis.

(4) **ANGPTL3**. ANGPTL3 is a well-known drug target that yielded a significant MR estimate in our analysis with TGs. Vupanorsen (an ANGPTL3 inhibitor) is an example of a drug that significantly reduced plasma TGs levels in two phase 1 trials in healthy adults, as well as in a phase 2 trial in type 2 diabetes patients with hepatic steatosis and hypertriglyceridemia [39]. Aligning with these observations, our MR results found that lower levels of ANGPTL3 were associated with reduced levels of TGs (*β* > 0); however, ANGPTL3 did not have a significant MR estimate with CAD, even though the direction of association was consistent (OR > 1) with the TGs result. Further investigation is needed to confirm whether ANGPTL3 influences CAD risk.

(5) **CELSR2/SORT1 region**. Our pipeline showed that CELSR2 showed a significant protective effect across LDL-C, TGs, and CAD, with consistent direction (*β* < 0, OR < 1). This result is consistent with prior work that has shown that genetic variants in the CELSR2-PSRC1-SORT1 gene cluster have strong associations with circulating lipid levels and CAD [40]. However, the significant heterogeneity between IVs and inconsistent MR estimates across multiple methods could be indicative of locus complexity. Tan *et al*. [40] found that lower CELSR2 expression in hepatocytes (liver cells) reduces lipid accumulation—contrasting with the protective effect the pipeline identified—suggesting the need for further locus-specific functional analysis.

(6) **TNF**. Studies with clinical and preclinical data have provided mixed results on whether drugs targeting TNF definitively reduce cardiovascular risk (e.g., [41]). For instance, one randomized controlled trial in psoriasis suggested that TNF inhibitors lowered IL6, which is causal for atherosclerosis and CVD events [42]; by contrast, evidence from previous clinical trials indicated worsening cardiac function upon inhibition of TNF in patients with preexisting heart failure [43], suggesting that the effects may be patient/cohort dependent. A recent meta-analysis further found that anti-TNF-a antibodies were associated with higher risk of major cardiovascular events (MACE) compared to placebo [44]. Our pipeline showed that for both TGs and CAD, increased TNF levels may have a protective effect against cardiovascular risk factors, consistent with the latter two studies. While we emphasize caution in the interpretation of our results due to a lack of evidence for colocalization in both the TGs and CAD analyses, they can contribute to the ongoing debate regarding the benefits and risks associated with TNF inhibition.

### New targets

(1) **BRAP**. While the analysis between BRAP and CAD yielded only one IV, this IV had a significant positive MR estimate with strong evidence for colocalization. Our observations were consistent with previous evidence linking BRAP to myocardial infarction, carotid atherosclerosis, and lipid metabolism (see e.g., [45]), but we encourage caution in the interpretation of this result, as the MR estimate for CAD relies on only one instrument and its direction is inconsistent with that of the estimate for LDL-C in Stage 1.

(2) **FURIN**. FURIN was also identified as a potential target in our Stage 1 and Stage 2 analyses, although there was no evidence for colocalization and the pipeline detected signs of heterogeneity and pleiotropy in Stage 2. FURIN has only recently been identified as having a potential causal association with ischemic heart disease, as well as other ASCVD-related phenotypes, in populations of European and East Asian ancestry [46]. It may also play a role in regulating lipid metabolism and may be associated with lipid levels in the bloodstream ([47]-[49]). In addition, elevated levels of FURIN may influence the progression of atherosclerosis ([46], [50]); while our results with CAD seem to be in alignment with these findings, our analysis with LDL-C suggested that higher FURIN levels would reduce levels of LDL-C, denoting an inconsistent direction of association as compared to CAD.

(3) **INHBC**. Another recent study that utilized MR found that circulating INHBC is causally linked with multiple cardiometabolic diseases and traits such as reduced lower-body fat, dyslipidemia, and increased risk of CAD and nonalcoholic fatty liver disease [51], part of which is in almost direct agreement with our MR results from analysis with TGs levels and CAD risk.

In sum, our study highlighted several possible new avenues of investigation for the development of drugs for cardiovascular risk factors.

### Limitations of the study

Our new drug repurposing pipeline identified a significant number of drug target candidates when applied to ASCVD, but there were some limitations. Firstly, our study was limited by the UKB-PPP study; if a protein did not show a valid IV in the UKB-PPP, it was not tested in our example application. Incorporating multiple proteomics databases (such as deCODE, ARIC, etc.) would increase the number of tested proteins and therefore the number of potential drug targets (the UKB-PPP itself will increase the number of tested proteins that are publicly available to ∼5,400 in the next year). Secondly, since the UKB-PPP measures soluble proteins from plasma, caution should be used when interpreting the observations from proteins with distinct functions between soluble and membrane-bound forms, just as we observed with LDLR. Thirdly, our study only used summary statistics from cohorts of European ancestry. Performing a similar analysis across multiple ancestries is important to obtain a more complete view on which investigative paths to pursue regarding drug development for cardiovascular risk. Fourthly, our study only focused on cis-acting pQTLs within or close to the gene region. While trans-acting variants (i.e., variants positioned outside the gene region of interest) can increase power, they are usually considered to be less robust proxies due to their potential influence on other genes and biological pathways [7]. Therefore, pleiotropy-robust modeling would be needed for a polygenic MR analysis (see e.g. MR-GRAPPLE [52]). Our pipeline can in fact expand the selection of variants to include those that are trans-acting, and applying such an analysis in conjunction with other methods may be appropriate in future studies. Finally, in the future, we intend to implement robust methods such as target trial emulation as an additional step for validation of identified drug candidates using real-world data.

### Conclusions

We have introduced a novel, high-throughput MR-based pipeline for drug repurposing and demonstrated its value in the context of lipid lowering and cardiovascular risk using GWAS and large-scale proteomics data. We identified MR associations for 72 protein drug targets for lowering LDL-C levels, and 75 such targets for lowering TGs levels; among them, 18 proteins showed potential causal associations with CAD risk, with 13 showing consistent directions of association between the lipids and CAD analyses. Further query in the DGIdb found 5 proteins associated with CAD risk listed as having interactions with approved drugs, which could possibly be repurposed to influence lipid levels and cardiovascular risk factors. Role of the Funder/Sponsor: The funders had no role in design and conduct of the study; collection, management, analysis, and interpretation of the data; preparation, review, or approval of the manuscript; and decision to submit the manuscript for publication.

## DECLARATIONS AND ACKNOWLEDGMENTS

### Ethics approval and consent to participate

This study was approved by the Institutional Review Board at VUMC and utilized data from the BioVU biobank at Vanderbilt University Medical Center, which was approved by the VICTR-BioVU Review Committee. All necessary participant consents were obtained, and data was de-identified.

### Consent for publication

Not applicable.

### Data and code availability

- Omicspred data are available at https://www.omicspred.org/
- BioVU data were derived from the <u>Vanderbilt University Medical Center (VUMC) BioVU database</u>, which is a controlled-access resource. Access is managed by the BioVU governance committee and can be requested by contacting.
- UKB-PPP data are publicly available at https://www.synapse.org/Synapse:syn51364943/wiki/622119
- GLGC data are publicly available at https://csg.sph.umich.edu/willer/public/glgc-lipids2021/
- CAD GWAS meta-analysis data are available at https://www.ebi.ac.uk/gwas/studies/GCST90132314
- MRDRP (pipeline) code: Available at https://github.com/smundo/MRDRP.
- Bayesian colocalization code: Available at https://chr1swallace.github.io/coloc/

### Declaration of Interests

The authors have declared no competing interests for this work.

### Funding acknowledgements

R01HL133786 (W.Q.W.), R01AG069900 (W.Q.W.), R01HL163854 (Q.F.), R01AG084550 (W.Q.W., Q.F.), R01HL171809 (W.Q.W., Q.F.), and the Vanderbilt Faculty Research Scholar Fund (Q.F.).

Dataset(s) were obtained from Vanderbilt University Medical Center’s BioVU, which is supported by institutional funding, 1S10RR025141-01, and CTSA grants UL1TR002243, UL1TR000445, and UL1RR024975; the UKB-PPP via the Synapse client; and the GWAS catalog.

### Contributors

Conceptualization (S.M., Q.F., W.Q.W.); data curation (S.M., Y.X., S.S.); formal analysis (S.M., Y.X., S.S.); investigation (S.M., Q.F.); funding acquisition (W.Q.W., Q.F.); methodology (S.M., Q.F., A.J.C.); validation (S.M., Q.F.); project administration (Q.F., W.Q.W.); visualization (S.M., A.L.D.); supervision (Q.F., W.Q.W.); resources (Y.X., S.S., W.Q.W., Q.F.); writing – original draft (S.M., Q.F.); and writing - review and editing (S.M., Q.F., A.L.D., C.M.S., M.G., B.L., L.L., W.Q.W.)

The first and corresponding authors had full access to all the data in the study and take responsibility for the integrity of the data and the accuracy of the data analysis. All authors read and approved the final version of the manuscript.

## Notes

### Author Declarations

Ethics committee/IRB of Vanderbilt University Medical Center gave ethical approval for this work. The study utilized data from the BioVU biobank at Vanderbilt University Medical Center, which was approved by the Vanderbilt Institute for Clinical and Translational Research-BioVU Review Committee. All necessary participant consents were obtained, and data was de-identified

### Summary of Updates

Updated title and abstract, as well as including an AI component to the study (reflected mostly in results and methods sections). Minor additional edits throughout.

