## Supplementary material for "A Mendelian randomization-based drug repurposing pipeline with integrated AI-facilitated prioritization: application to lipid traits and coronary artery disease": Supplementary File 8-CAD_prompt_new.docx

**Prompt 1:**

You are provided with the following list of 63 drugs identified from DGIdb as candidates for repurposing in the treatment of coronary artery disease. Each drug is associated with a protein target, an interaction type (where available), and an original indication from DGIdb (where available).

Note: DGIdb-reported indications are drawn from heterogeneous, sometimes noisy source databases and may not reflect the drug's actual FDA-approved indication(s). Part of this task is to independently verify each DGIdb_Indication against the drug's real, currently FDA-approved indication(s).

Note: Where a field below asks for supporting references, only include a citation, PMID, DOI, or NCT number if you are confident it is real. Do not fabricate references to fill a field — an honest "no specific reference identified" is preferable to an invented one.

Drug list (in order as provided):

Drug: donanemab | Target_Protein: APOE | Interaction_Type: unknown | DGIdb_Indication: not available

Drug: norethindrone acetate | Target_Protein: APOE | Interaction_Type: unknown | DGIdb_Indication: not available

Drug: lorazepam | Target_Protein: APOE | Interaction_Type: unknown | DGIdb_Indication: Anti-anxiety Agents; Anticonvulsants; Hypnotics and Sedatives

Drug: choriogonadotropin alfa | Target_Protein: APOE | Interaction_Type: unknown | DGIdb_Indication: Fertility Agents, antineoplastic agent

Drug: lecanemab | Target_Protein: APOE | Interaction_Type: unknown | DGIdb_Indication: not available

Drug: galantamine | Target_Protein: APOE | Interaction_Type: unknown | DGIdb_Indication: for treatment of alzheimer's disease

Drug: ritonavir | Target_Protein: APOE | Interaction_Type: unknown | DGIdb_Indication: not available

Drug: aducanumab | Target_Protein: APOE | Interaction_Type: unknown | DGIdb_Indication: for treatment of alzheimer's disease

Drug: donepezil | Target_Protein: APOE | Interaction_Type: unknown | DGIdb_Indication: for treatment of alzheimer's disease

Drug: lutein | Target_Protein: APOE | Interaction_Type: unknown | DGIdb_Indication: not available

Drug: nicotine polacrilex | Target_Protein: APOE | Interaction_Type: unknown | DGIdb_Indication: Central Nervous System Stimulants

Drug: troglitazone | Target_Protein: APOE | Interaction_Type: unknown | DGIdb_Indication: not available

Drug: triamcinolone | Target_Protein: APOE | Interaction_Type: unknown | DGIdb_Indication: for treatment of diabetic macular edema

Drug: ganciclovir | Target_Protein: APOE | Interaction_Type: unknown | DGIdb_Indication: not available

Drug: bupropion hydrochloride | Target_Protein: APOE | Interaction_Type: unknown | DGIdb_Indication: smoking-cessation agent, appetite suppressant, antidepressant

Drug: apixaban | Target_Protein: APOE | Interaction_Type: unknown | DGIdb_Indication: antithrombotic

Drug: prednisone | Target_Protein: APOE | Interaction_Type: unknown | DGIdb_Indication: corticosteroid, antiinflammatory agent

Drug: rivastigmine | Target_Protein: APOE | Interaction_Type: unknown | DGIdb_Indication: for treatment of Alzheimer's disease

Drug: midostaurin | Target_Protein: TNF | Interaction_Type: unknown | DGIdb_Indication: antineoplastic agent

Drug: rutin | Target_Protein: TNF | Interaction_Type: unknown | DGIdb_Indication: not available

Drug: naproxen sodium | Target_Protein: TNF | Interaction_Type: unknown | DGIdb_Indication: NSAID, antimigraine agent

Drug: bupivacaine hydrochloride | Target_Protein: TNF | Interaction_Type: unknown | DGIdb_Indication: not available

Drug: vitamin b6 | Target_Protein: TNF | Interaction_Type: unknown | DGIdb_Indication: not available

Drug: procarbazine hydrochloride | Target_Protein: TNF | Interaction_Type: unknown | DGIdb_Indication: not available

Drug: pentoxifylline | Target_Protein: TNF | Interaction_Type: unknown | DGIdb_Indication: for treatment of amyotrophic lateral sclerosis (ALS)

Drug: masoprocol | Target_Protein: TNF | Interaction_Type: unknown | DGIdb_Indication: prostate cancer, antineoplastic agent

Drug: fluocinolone acetonide | Target_Protein: TNF | Interaction_Type: unknown | DGIdb_Indication: glucocorticoid, antiinflammatory agent

Drug: glimepiride | Target_Protein: TNF | Interaction_Type: unknown | DGIdb_Indication: antidiabetic

Drug: miltefosine | Target_Protein: TNF | Interaction_Type: unknown | DGIdb_Indication: not available

Drug: mycophenolate mofetil | Target_Protein: TNF | Interaction_Type: unknown | DGIdb_Indication: immunosuppressant

Drug: hydroxychloroquine | Target_Protein: TNF | Interaction_Type: unknown | DGIdb_Indication: antirheumatic agent

Drug: gentamicin | Target_Protein: TNF | Interaction_Type: unknown | DGIdb_Indication: not available

Drug: propylthiouracil | Target_Protein: TNF | Interaction_Type: unknown | DGIdb_Indication: Antithyroid Agents

Drug: methimazole | Target_Protein: TNF | Interaction_Type: unknown | DGIdb_Indication: Antithyroid Agents

Drug: carbamazepine | Target_Protein: TNF | Interaction_Type: unknown | DGIdb_Indication: for treatment of bipolar disorder

Drug: rabeprazole | Target_Protein: TNF | Interaction_Type: unknown | DGIdb_Indication: Proton pump inhibitor

Drug: cromolyn sodium | Target_Protein: TNF | Interaction_Type: unknown | DGIdb_Indication: not available

Drug: amphotericin b liposomal | Target_Protein: TNF | Interaction_Type: unknown | DGIdb_Indication: not available

Drug: meropenem anhydrous | Target_Protein: TNF | Interaction_Type: unknown | DGIdb_Indication: not available

Drug: stavudine | Target_Protein: TNF | Interaction_Type: unknown | DGIdb_Indication: not available

Drug: didanosine | Target_Protein: TNF | Interaction_Type: unknown | DGIdb_Indication: not available

Drug: insulin, regular, human | Target_Protein: TNF | Interaction_Type: unknown | DGIdb_Indication: for treatment of diabetic foot ulcers, antidiabetic

Drug: indomethacin | Target_Protein: TNF | Interaction_Type: unknown | DGIdb_Indication: NSAID

Drug: gemcitabine | Target_Protein: TNF | Interaction_Type: unknown | DGIdb_Indication: antineoplastic agent

Drug: docetaxel anhydrous | Target_Protein: TNF | Interaction_Type: unknown | DGIdb_Indication: antineoplastic agent

Drug: cyclosporine | Target_Protein: TNF | Interaction_Type: unknown | DGIdb_Indication: immunosuppressant, opthalmological agent

Drug: thalidomide | Target_Protein: TNF | Interaction_Type: unknown | DGIdb_Indication: antineoplastic agent

Drug: methylene blue | Target_Protein: TNF | Interaction_Type: unknown | DGIdb_Indication: not available

Drug: diclofenac sodium | Target_Protein: TNF | Interaction_Type: unknown | DGIdb_Indication: for treatment of glaucoma, analgesic, NSAID

Drug: midazolam hydrochloride | Target_Protein: TNF | Interaction_Type: unknown | DGIdb_Indication: Anesthesia; Hypnotics and Sedatives, Adjuvants

Drug: rifampin | Target_Protein: TNF | Interaction_Type: unknown | DGIdb_Indication: not available

Drug: ketorolac tromethamine | Target_Protein: TNF | Interaction_Type: unknown | DGIdb_Indication: antimigraine agent, NSAID

Drug: sorafenib | Target_Protein: TNF | Interaction_Type: unknown | DGIdb_Indication: antineoplastic agent

Drug: lactulose | Target_Protein: TNF | Interaction_Type: unknown | DGIdb_Indication: not available

Drug: ibuprofen, sodium salt | Target_Protein: TNF | Interaction_Type: unknown | DGIdb_Indication: NSAID

Drug: cefotaxime sodium | Target_Protein: TNF | Interaction_Type: unknown | DGIdb_Indication: not available

Drug: pyrazinamide | Target_Protein: TNF | Interaction_Type: unknown | DGIdb_Indication: not available

Drug: carboplatin | Target_Protein: TNF | Interaction_Type: unknown | DGIdb_Indication: not available

Drug: omeprazole | Target_Protein: TNF | Interaction_Type: unknown | DGIdb_Indication: Proton pump inhibitor, antiulcer agent

Drug: ethambutol hydrochloride | Target_Protein: TNF | Interaction_Type: unknown | DGIdb_Indication: not available

Drug: risperidone | Target_Protein: TNF | Interaction_Type: unknown | DGIdb_Indication: Antipsychotic Agents, antipsychotic agent

Drug: prazosin hydrochloride | Target_Protein: TNF | Interaction_Type: unknown | DGIdb_Indication: not available

Drug: cisplatin | Target_Protein: TNF | Interaction_Type: unknown | DGIdb_Indication: not available

Using this fixed list, rank all 63 drugs in descending order of their potential effectiveness for coronary artery disease repurposing (Rank 1 = highest potential).

For each drug, provide the following information in JSON format using the specified keys:

Rank: Integer rank (1–63), with 1 representing the highest potential effectiveness

Drug: Generic name of the drug

Target_Protein: Protein target as listed above (APOE or TNF)

Interaction_Type: Interaction type as listed above (inhibitor or unknown)

DGIdb_Indication: Original indication as listed above, verbatim

DGIdb_Indication_Accuracy: Assessment of whether DGIdb_Indication accurately reflects a real, currently FDA-approved indication for this drug. Use one of: "Accurate" (matches an actual FDA-approved indication), "Partially Accurate" (broadly correct but imprecise, outdated, or incomplete), "Inaccurate" (does not correspond to any FDA-approved indication for this drug), or "Not Verifiable" (DGIdb_Indication is "not available" or too vague to assess)

FDA_Approved_Indications: The drug's actual, currently FDA-approved indication(s), stated independently of what DGIdb reported. If the drug is not FDA-approved (e.g., approved only outside the US, or investigational), state that explicitly.

Disease: Broader original indication(s) for which the drug was developed or approved (may expand on FDA_Approved_Indications)

Reasons_to_List: Scientific or clinical rationale supporting its potential repurposing for coronary artery disease (e.g., mechanism of action, pathway relevance, preclinical or clinical evidence)

Safety_Profile: Summary of the toxicity profile and suitability for chronic administration (typically ≥6 months), including key safety considerations

Active_Comparator_Availability: Availability of a credible active comparator, i.e., other approved drugs sharing the same primary indication as the candidate drug

Expected_Bias_From_Indication: Expected level of confounding or bias introduced by the original indication of the drug when studying its effect on coronary artery disease outcomes (minimal/moderate/high), with a brief explanation. Note explicitly if an inaccurate DGIdb_Indication would materially change this assessment (i.e., bias should be evaluated against the true FDA-approved indication, not the possibly-erroneous DGIdb_Indication).

General_Population_Use: Estimate or qualitative description of how commonly the drug is used in the general population (e.g., widely prescribed, moderate use, rare use; optionally include approximate annual prescription volume if known)

Retrospective_Cohort_Studies: Summary of any published or ongoing retrospective cohort studies examining the association between this drug and coronary artery disease outcomes (e.g., MACE, MI, revascularization, CAD progression). Include study population, data source, key findings, and approximate publication year where known. If no such studies are identified, state "No retrospective cohort studies on coronary artery disease outcomes identified."

Clinical_Trials: Summary of any ongoing or recently completed clinical trials investigating this drug specifically for the treatment or prevention of coronary artery disease. Include trial phase, NCT number, and brief description where known. If no coronary artery disease-specific trials are identified, state "No coronary artery disease-specific clinical trials identified."

Key_References: 2–4 of the most important references supporting the information above (not an exhaustive bibliography) — e.g., the FDA prescribing information, a pivotal retrospective cohort or trial paper, or a key mechanistic study. For each, give enough detail to locate it (first author, journal, year, and PMID or DOI if known; NCT number for trials; "FDA prescribing information" for label-derived facts). If no specific reference can be confidently identified, state "No specific reference identified" rather than inventing one.

Present the output strictly in valid JSON format as an array of 63 ranked objects, with no additional commentary or markdown.

**Prompt 2:**

Note: Where a field requires a citation, PMID, DOI, or NCT number (see Key_References), only treat it as satisfying the criteria below if it follows the required format — you cannot independently confirm a reference actually exists unless you have a retrieval tool available, so verification here is at the format level, not a guarantee of factual accuracy.

Please verify whether the previously generated list satisfies all required criteria:

(1) The list contains exactly the 63 drugs from the original list provided earlier, with no additions, omissions, or duplicates by generic name;

(2) For each drug, the Target_Protein, Interaction_Type, and DGIdb_Indication values in the generated JSON exactly match those in the original list provided earlier, verbatim and unaltered;

(3) None of the drugs were originally developed specifically for coronary artery disease, and all are appropriate candidates for coronary artery disease repurposing consideration;

(4) Each JSON object contains all required keys: Rank, Drug, Target_Protein, Interaction_Type, DGIdb_Indication, DGIdb_Indication_Accuracy, FDA_Approved_Indications, Disease, Reasons_to_List, Safety_Profile, Active_Comparator_Availability, Expected_Bias_From_Indication, General_Population_Use, Retrospective_Cohort_Studies, Clinical_Trials, and Key_References;

(5) DGIdb_Indication_Accuracy is populated with exactly one of "Accurate", "Partially Accurate", "Inaccurate", or "Not Verifiable", and FDA_Approved_Indications states either a real FDA-approved indication or an explicit non-approval statement — neither field is left blank;

(6) Key_References contains no more than 4 entries per drug, each entry follows the required format (first author, journal, and year plus a PMID or DOI; an NCT number for trials; or "FDA prescribing information" for label-derived facts), and the field states "No specific reference identified" rather than being left blank where no reference applies;

(7) The drugs are clearly ranked in descending order of potential effectiveness for coronary artery disease (Rank 1 = highest potential, Rank 63 = lowest);

(8) The output is presented strictly in valid JSON format as an array of 63 objects, with no additional commentary or markdown.

If the input JSON fully satisfies all specified requirements, return it exactly as provided without any modifications. If the input JSON does not meet any of the requirements, regenerate a corrected list that fully complies with all criteria and output only the revised valid JSON.
