## Supplementary material for "A Mendelian randomization-based drug repurposing pipeline with integrated AI-facilitated prioritization: application to lipid traits and coronary artery disease": Supplementary_Material_new.docx

**Supplementary Methods §1. Data Sources**

**OmicsPred**

OmicsPred is a resource that uses genotypes to directly predict multi-omics data. As part of its data production pipeline, it uses the INTERVAL cohort (*N*=50,000 participants) and plasma protein levels available from the SomaScan (*N*=3,175) and Olink (*N*=4,822) platforms to produce proteome-wide GWAS summary statistics [18]. We extracted these summary statistics for each protein for individuals of European ancestry and used them to calculate protein-level polygenic risk scores (PRS) as part of optimizing Mendelian randomization (MR) significance levels for application of this pipeline to cardiovascular traits (see Supplementary Methods §2 for additional details).

**VUMC BioVU biobank**

The Vanderbilt University Medical Center (VUMC) BioVU biobank is a biorepository of de-identified samples of plasma and DNA that are linked to de-identified electronic health records (EHRs) [19]. BioVU offers genome-wide genotyping data for ~95,000 BioVU patients on the Illumina Infinium Expanded Multi-Ethnic Genotyping Array plus custom content platform (MEGA). For the patients with EHR-reported White race who have been genotyped on this platform, we gathered age, sex, medication use (i.e., statins, fibrates, niacin, bile acid sequestrants, and omega-3 fatty acids), low-density lipoprotein cholesterol (LDL-C), triglycerides (TGs) from the EHRs. From these, we further excluded individuals without genetically determined majority European ancestry and constructed two cohorts as part of optimizing MR significance levels for application of this pipeline to cardiovascular traits: one for individuals with LDL-C measurements, and a second for with TGs measurements (see Supplementary Methods §2 for additional details).

**UKB-PPP**

The UK Biobank Pharma Proteomics Project (UKB-PPP) is a biopharmaceutical consortium with the plasma proteomic profiles of 54,219 UKB participants and has measured proteomic associations with genetics and health in the UKB [12]. This data includes pQTL mapping and GWAS summary statistics for 2,923 proteins. The project reports 14,287 significant associations across all proteins; however, these results only include the most significant associations at each locus. Indeed, the total number of potentially significant variants per protein number in the hundreds. This database offers breadth and robustness for pQTL discovery, given the sheer number of significant variants it could yield per protein. As such, we used the UKB-PPP summary statistics as the source for the exposure data in our MR analysis Stages 1 and 2, focusing on the European ancestry GWAS for the application of this pipeline to cardiovascular traits.

**GLGC**

The Global Lipids Genetics Consortium (GLGC) [20] is a worldwide collaboration that studies the genetic basis of quantitative lipid traits, including total cholesterol (TC), LDL-C, high-density lipoprotein cholesterol (HDL-C), and TGs. One of its recent studies involves a comprehensive, multi-ancestry GWAS meta-analysis of lipid levels in approximately 1.65 million individuals. For the application of this pipeline to cardiovascular traits, we utilized summary statistics from European-ancestry GWAS for LDL-C and TGs (*N*=1,320,016 individuals) as the source for the main outcome data in the MR analysis, excluding individuals from the UKB (*N*=369,130) to avoid sample overlap between testing (Stage 1) and external validation (Stage 2).

**CAD GWAS meta-analysis**

We used large GWAS meta-analysis (*N*=1,165,690) for CAD by Aragam *et al*. [21], with 181,522 cases among participants of predominantly European ancestry from at least 10 different countries for Stage 2 of the application of this pipeline to cardiovascular traits. This meta-analysis identified 241 genetic associations out of 20,073,070 variants included in the analysis.

**Supplementary Methods §2. Evaluation and optimization of instrument selection threshold for proteome-wide MR**

There are multiple considerations for determining optimized significance level for Mendelian randomization studies. For example, applying a genome wide significance threshold (*p*<5x10^-8^) can be overly punitive for the selection of cis-acting pQTLs in much smaller gene regions, as is the case in this pipeline. As such, we tested several thresholds for our example of cardiovascular disease, derived in part from data independent of the pipeline. In short, we first used summary statistics from OmicsPred, including Somascan and Olink, to calculate polygenic risk scores for proteins in cohorts of BioVU patients (one cohort of patients with TG measures and one with LDL-C measures). We then used linear regression to test these Somascan- and Olink-derived PRS for associations with measured LDL-C and TGs (*p*<0.05) in BioVU. We next used summary statistics for those significant proteins (UKB-PPP) and lipids (GLGC) to perform MR between protein levels and lipids across 3 different variant selection significance thresholds (*p*<5x10^-8^, 10^-5^, and 10^-4^) and identified the optimal threshold for use in the ensuing proteome-wide MR analysis (Stage 1 of the pipeline).

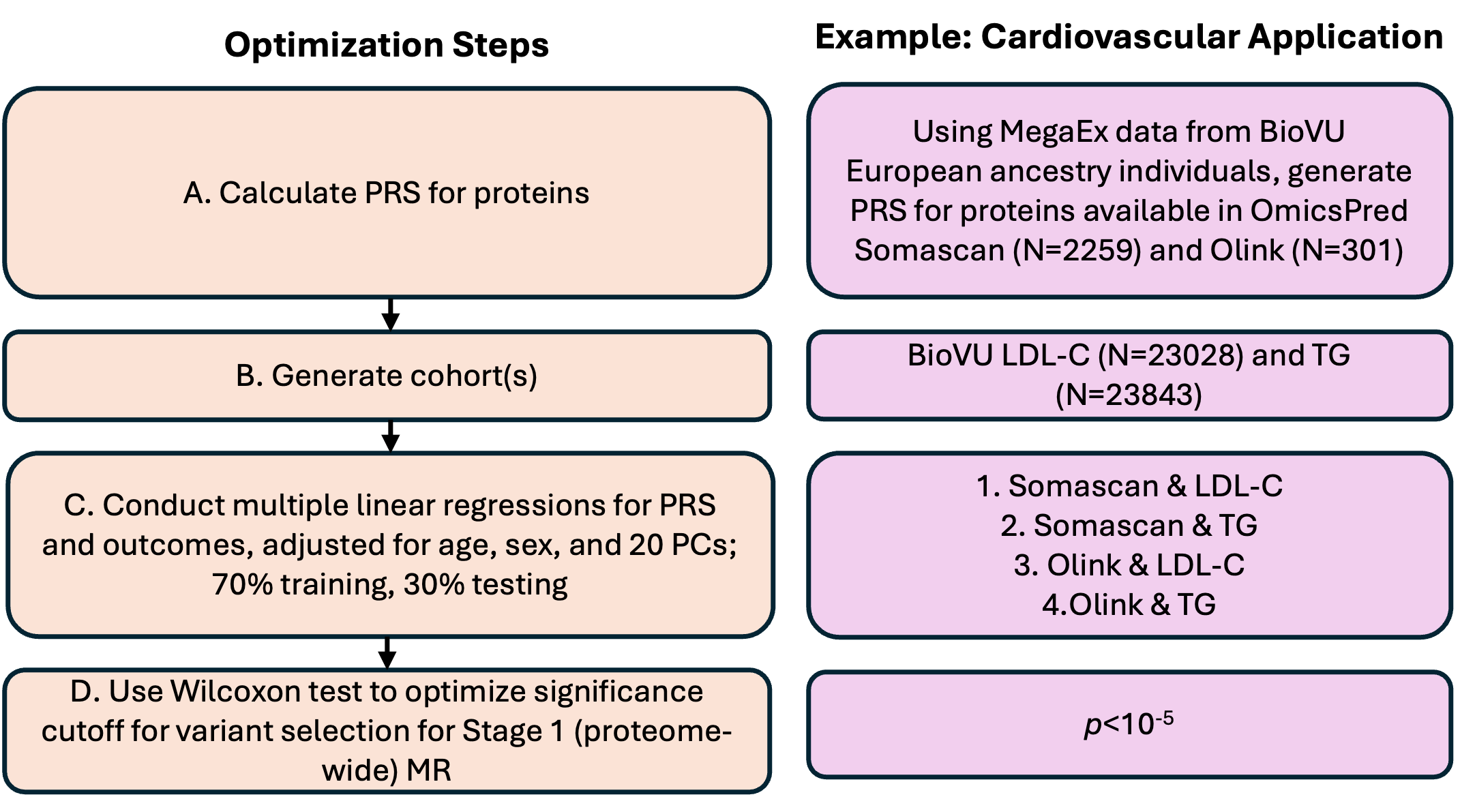

**Supplementary Figure 1. Steps to optimize significance threshold for MR.** PRS=polygenic risk score; BioVU=Vanderbilt University Medical Center BioVU biobank; LDL-C=low-density lipoprotein cholesterol; TG= triglyceride; PC=principal component.

**A. Calculate the PRS for proteins.**

We first selected all individuals with EHR-reported white race in BioVU with available genotyping data from the Illumina Infinium Expanded Multi-Ethnic Genotyping Array (MegaEx; *N*=94,863); we then imputed these data as previously reported [53]. We calculated principal component (PC) scores and prediction of ancestry memberships for the patients using the FRAPOSA pipeline [54], which utilizes a computationally efficient singular value decomposition algorithm to predict the ancestry of study samples via a principal component analysis (PCA) with a reference panel (see e.g. [55] and references therein). We restricted subsequent analyses to individuals with majority European ancestry (see below). We then used the imputed MegaEx genetic data from BioVU and OmicsPred’s effect sizes from the protein-level GWAS summary statistics produced with the INTERVAL cohort, including SomaScan (N=3,175) and Olink (N=4,822), to calculate PRS for proteins with the pgsc_calc pipeline [18]. We successfully calculated PRS for 2,259 proteins in the SomaScan platform and 301 proteins in the Olink platform [18].

**B. Generate cohorts for association testing.**

We generated 2 cohorts of EA patients from BioVU with available MegaEx genotyping data and who had at least one measurement of LDL-C or TGs, respectively, in their EHRs. For both cohorts, we extracted individuals’ cohort-related lipid measurements (i.e., LDL-C or TGs) with adjustment for lipid-lowering medication usage following the criteria described in [56]. Specifically, if any patient that had taken medication (i.e., statins, fibrates, niacin, and/or bile acid sequestrants for LDL-C; statins, fibrates, niacin, and/or omega-3 fatty acids for TGs) and had laboratory measurements after that, we adjusted the lipid level by adding a constant according to the type of medication, as in [56]. If a patient had taken more than one type of medication, we chose the largest constant for measurement adjustment. We then removed any measurements that were more than three times the interquartile range (IQR) above the median of the full distribution of measurements for all patients and calculated the median lipid measurements (LDL-C and TGs, respectively) for each patient in the LDL-C (*N*=23,028) and TGs (*N*=23,843) cohorts (see demographics summary below and **Supplementary Files 13-14** for lists of basic demographics per de-identified patient, for each cohort).

| **Cohort** | **Age [years] (mean±SD)** | **Sex [*N* (%)]** |
| --- | --- | --- |
| LDL-C (*N*=23,028) | 68±20 | Male: 10784 (47%); Female: 12244 (53%) |
| Triglycerides (*N*=23,843) | 68±20 | Male: 11217 (47%); Female: 12626 (53%) |

**Table S2.** Summary of basic demographics for each lipid cohort used for association testing. Both cohorts consisted of patients who reported white race and had genetically predicted European ancestry. SD=standard deviation.

**C. Conduct multiple linear regression between protein PRS and measured lipids (TGs and LDL-C)**

To optimize the variant selection threshold for MR, we first created a subset of proteins by performing multiple linear regressions to test the associations between protein PRS and the lipids measurements. Because SomaScan and Olink quantify proteins using distinct chemistries, scales, and pQTL architectures, we analyzed them in separate multiple linear regressions (adjusted for age, sex, and 20 PCs). This avoids cross-platform scale/assay confounding, reduces collinearity, and preserves interpretability, in addition to avoiding double counting of proteins. For each regression, we randomly selected 70% of the data to use as a training set, with the remaining 30% to be used as a testing set to evaluate the performance of the model. We also added a constant to each of the predictor data sets to allow for the possibility of an intercept. We used R^2^ (the proportion of the variance in the dependent variable explained by the independent variable) and the root mean squared error (RMSE) as the metrics to probe the fit from the training set and model performance from the testing set, respectively.

For LDL-C, SomaScan-based PRS explained 22% of the variance (R^2^=0.22, *p*-value of *F*-statistic=2.79x10^-131^; **Table S3**). Using this model, we predicted levels of LDL-C from the predictor testing set and calculate RMSE=38.8 mg/dL between predicted values of LDL-C levels and values in the outcome testing set. This result accounts for only 14% of the full range of observed LDL-C measurement values, consistent with an adequate performance by the model.

Similarly, Olink-based PRS explained 12% of variation (R^2^=0.12, p-value 1.28x10^-255^, RMSE=33.07 mg/dL). Across both platforms, we identified 154 distinct proteins whose PRS had a significant (*p*_coefficient_<0.05) association with LDL-C levels, 85 of which overlapped with the UKB-PPP.

For log-transformed TGs, SomaScan-based PRS explained 10% of variance (see **Table S3**, R^2^=0.10, P-value=5.02x10^-151^, RMSE = 0.45), accounting for 13.5% of the full range of observed log(TGs) measurement values; Olink-based PRS explained 8% of the variance (R^2^=0.08, p-value=2.79x10^-131^, RMSE=0.51). These analyses identified 147 distinct proteins whose PRS had a significant association (*p*_coefficient_<0.05) with TGs levels, with 68 of them appearing in the UKB-PPP.

| **Regression** | **R^2^** | ***p*-value (*F*-Statistic)** | **RMSE** | **Significant proteins (*p*<0.05)** |
| --- | --- | --- | --- | --- |
| **LDL-C & SomaScan PRS** | 0.22 | 3.06x10^-125^ | 38.8 mg/dL | 154 |
| **LDL-C & Olink PRS** | 0.12 | 1.28x10^-255^ | 33.07 mg/dL |  |
| **log(TGs) & SomaScan PRS** | 0.10 | 5.02x10^-151^ | 0.45 | 147 |
| **log(TGs) & Olink PRS** | 0.08 | 2.79x10^-131^ | 0.51 |  |

**Table S3.** Results for regression between lipids and protein PRS. R^2^ probes the fit from the training set, while RMSE evaluates the model performance.

**D. Threshold optimization via MR on protein subsets**

In the BioVU LDL-C cohort, we identified 154 distinct proteins whose PRS had a significant association with LDL-C levels. At the genome-wide significance threshold (*p*<5×10⁻⁸), 72 proteins from the UKB-PPP were available for MR analysis; we found nine suggestive associations, five of which were significant **(Table S4).** NCAN showed evidence of heterogeneity and pleiotropy, suggesting caution in the interpretation and reliability of its results. In the BioVU TG cohort, our regression analysis identified 147 distinct proteins whose PRS were significantly associated with TGs levels. 60 proteins from the UKB-PPP were available for analysis at the genome-wide significance threshold, and we found six significant MR estimates (see **Table S5**), with no evidence of heterogeneity or pleiotropy.

We next compared these results with those from two additional thresholds for cis-pQTL selection (*p*<10⁻⁵ and *p*<10⁻⁴). We used a one-sided Wilcoxon signed-rank test to determine whether there were significant directional differences (*p*_Wilcoxon_ < 0.05) in the distributions of instrumental variable (IV) counts for significant proteins as the MR variant selection threshold was relaxed. For the LDL-C analysis, performing the Wilcoxon test on results from the first two variant selection thresholds (genome-wide significance vs *p*<10^-5^) yielded *p*_Wilcoxon_=0.034, suggesting a significant difference in IV count between the two. Performing the Wilcoxon test between *p*<10^-5^ and *p*<10^-4^ yielded *p*_Wilcoxon_=0.054, suggesting there was no significant difference in IV count between these two thresholds. The same test provided similar results for the TGs analysis, indicating that among the three thresholds tested, *p*<10^-5^ provided the most significant gain in IV count, while results at the most relaxed threshold (*p*<10^-4^) provided marginal additional gain (average increase of only ~1 IV per protein for both the LDL-C and TGs analyses), with higher risk of winner’s curse bias due to the less stringent threshold [29]. We therefore selected *p*<10^-5^ as the significance threshold for selection of the variants to be used in the proteome-wide MR analysis.

**Selection threshold: *p* < 5 × 10^-8^**

(72 proteins tested)

| **Protein name** | ***N*_IV_** | ***β*** | ***p_β_*** | ***p_Q_*_-test_** | ***p*_MR-Egger_** |
| --- | --- | --- | --- | --- | --- |
| LDLR | 1 | 0.26±0.08 | 1.3 × 10^-10^ | - | - |
| NCAN | 5 | 0.19±0.09 | 3.9 × 10^-5^ | 1.2 × 10^-19^ | 0.02 |
| OSM | 1 | -0.18±0.06 | 7.5 × 10^-10^ | - | - |
| PCSK9 | 5 | 0.37±0.01 | < 10^-200^ | 0.15 | 0.23 |
| RSPO3 | 3 | 0.05±0.03 | 4.1 × 10^-4^ | 0.11 | 0.30 |

**Selection threshold: *p* < 10^-5^**

(76 proteins tested)

| CD40 | 12 | 0.03±0.02 | 10^-4^ | 0.004 | 0.37 |
| --- | --- | --- | --- | --- | --- |
| LDLR | 1 | 0.26±0.08 | 1.3 × 10^-10^ | - | - |
| NCAN | 6 | 0.18±0.09 | 3.8 × 10^-4^ | 2.1 × 10^-29^ | 0.03 |
| OSM | 1 | -0.18±0.06 | 7.5 × 10^-10^ | - | - |
| PCSK9 | 9 | 0.37±0.02 | 1.9 × 10^-192^ | 1.3 × 10^-9^ | 0.89 |
| RSPO3 | 5 | 0.05±0.02 | 2.9 × 10^-6^ | 0.17 | 0.29 |

**Selection threshold: *p* < 10^-4^**

(78 proteins tested)

| CD40 | 12 | 0.03±0.02 | 10^-4^ | 0.004 | 0.37 |
| --- | --- | --- | --- | --- | --- |
| LDLR | 1 | 0.26±0.08 | 1.3 × 10^-10^ | - | - |
| NCAN | 7 | 0.18±0.08 | 7.9 × 10^-5^ | 6.9 × 10^-29^ | 0.03 |
| OSM | 1 | -0.18±0.06 | 7.5 × 10^-10^ | - | - |
| PCSK9 | 12 | 0.37±0.02 | < 10^-200^ | 7.5 × 10^-10^ | 0.68 |
| RSPO3 | 7 | 0.05±0.02 | 5.7 × 10^-8^ | 0.49 | 0.19 |

**Table S4.** MR results with LDL-C for 3 variant selection thresholds. Corresponding Bonferroni-corrected p-values for MR estimate significance were *p*<0.05/72, *p*<0.05/76, and *p*<0.05/78. Uncertainties are reported at the 95% confidence level.

**Selection threshold: *p* < 5 × 10^-8^**

(60 proteins tested)

| **Protein name** | ***N*_IV_** | ***β*** | ***p_β_*** | ***p_Q_*_-test_** | ***p*_MR-Egger_** |
| --- | --- | --- | --- | --- | --- |
| ADAMTS4 | 1 | 0.06±0.03 | 5.2 × 10^-5^ | - | - |
| CD14 | 10 | 0.03±0.01 | 9.0 × 10^-6^ | 0.39 | 0.33 |
| ITGAL | 1 | 0.15±0.06 | 1.6 × 10^-9^ | - | - |
| MB | 1 | 0.16±0.09 | 3.2 × 10^-4^ | - | - |
| MERTK | 3 | 0.04±0.01 | 1.7 × 10^-9^ | 0.33 | 0.94 |
| MSR1 | 8 | 0.024±0.008 | 1.5 × 10^-8^ | 0.47 | 0.55 |

**Selection threshold: *p* < 10^-5^**

(62 proteins tested)

| ADAMTS4 | 3 | 0.05±0.02 | 2.9 × 10^-5^ | 0.63 | 0.51 |
| --- | --- | --- | --- | --- | --- |
| CD14 | 14 | 0.03±0.01 | 1.3 × 10^-4^ | 0.07 | 0.26 |
| ITGAL | 1 | 0.15±0.06 | 1.6 × 10^-9^ | - | - |
| MB | 1 | 0.16±0.09 | 3.2 × 10^-4^ | - | - |
| MERTK | 7 | 0.04±0.01 | 7.4 × 10^-9^ | 0.78 | 0.33 |
| MSR1 | 15 | 0.02±0.01 | 2.6 × 10^-6^ | 0.26 | 0.20 |

**Selection threshold: *p* < 10^-4^**

(63 proteins tested)

| ADAMTS4 | 3 | 0.05±0.02 | 2.9 × 10^-5^ | 0.63 | 0.51 |
| --- | --- | --- | --- | --- | --- |
| CD14 | 15 | 0.03±0.01 | 6.7 × 10^-5^ | 0.09 | 0.29 |
| MB | 1 | 0.16±0.09 | 3.2 × 10^-4^ | - | - |
| MERTK | 10 | 0.04±0.01 | 1.7 × 10^-7^ | 0.64 | 0.24 |
| MSR1 | 17 | 0.02±0.01 | 1.5 × 10^-6^ | 0.39 | 0.19 |

**Table S5.** MR results with TGs for 3 variant selection thresholds. Corresponding Bonferroni-corrected p-values for MR estimate significance were *p*<0.05/60, *p*<0.05/62, and *p*<0.05/63. Uncertainties are reported at the 95% confidence level.
