## Supplementary figures and images for "A Mendelian randomization-based drug repurposing pipeline with integrated AI-facilitated prioritization: application to lipid traits and coronary artery disease"

### ABO_P16442_OID30675_v1_Inflammation_II.png

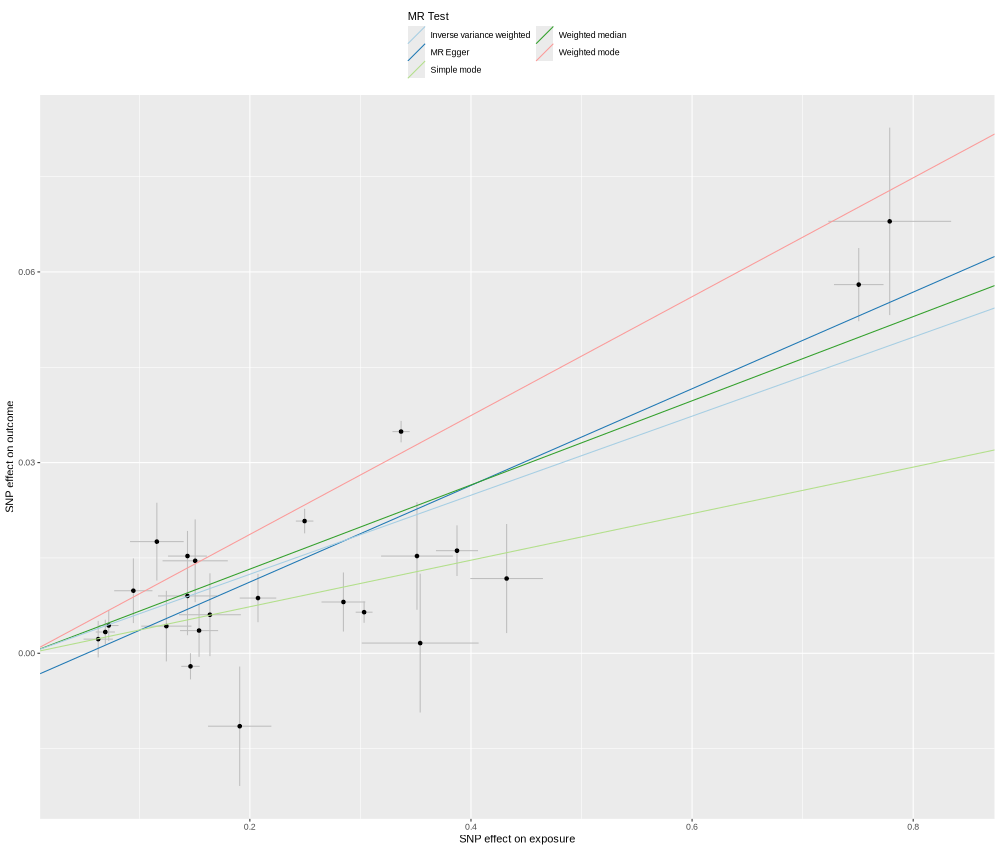

### ABO_P16442_OID30675_v1_Inflammation_II_singleSNP.png

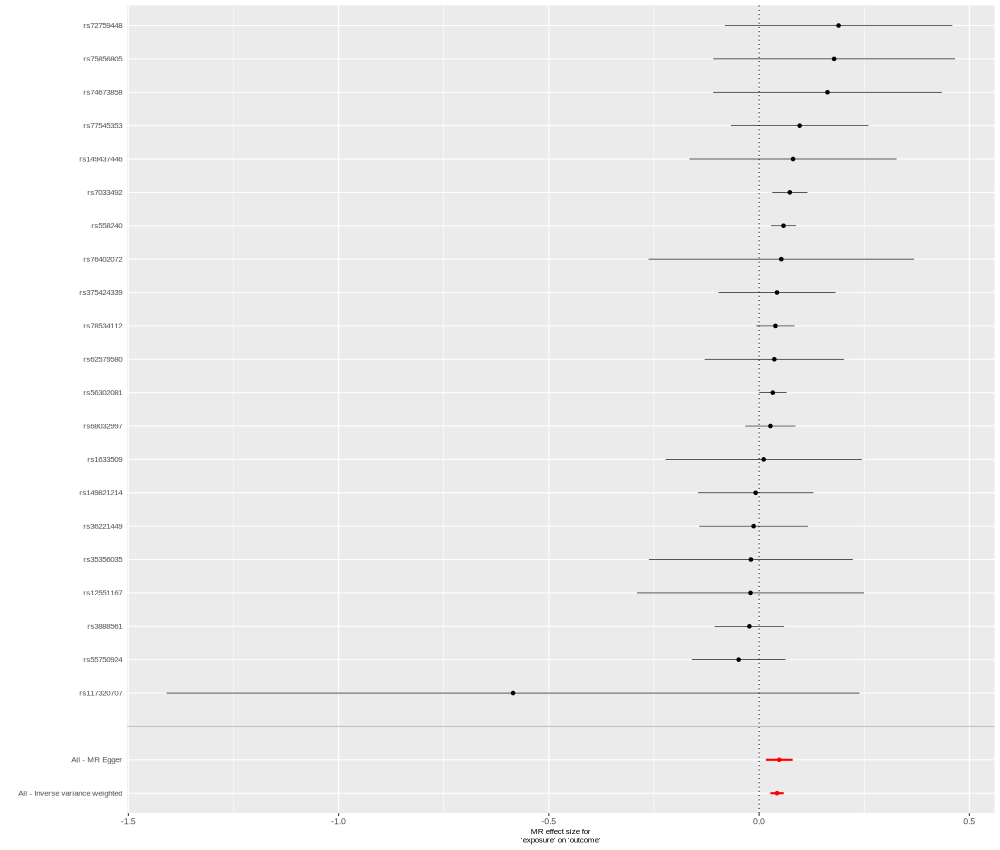

### ACE_P12821_OID30696_v1_Inflammation_II_singleSNP.png

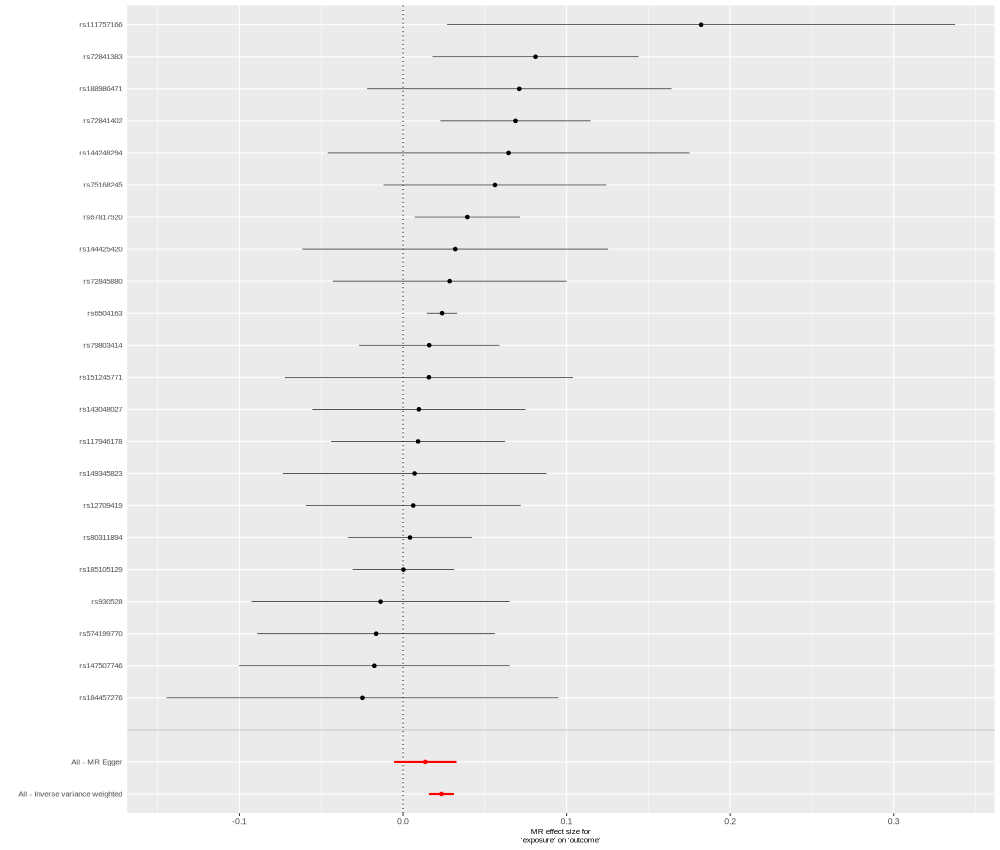

### ADM_P35318_OID21467_v1_Oncology.png

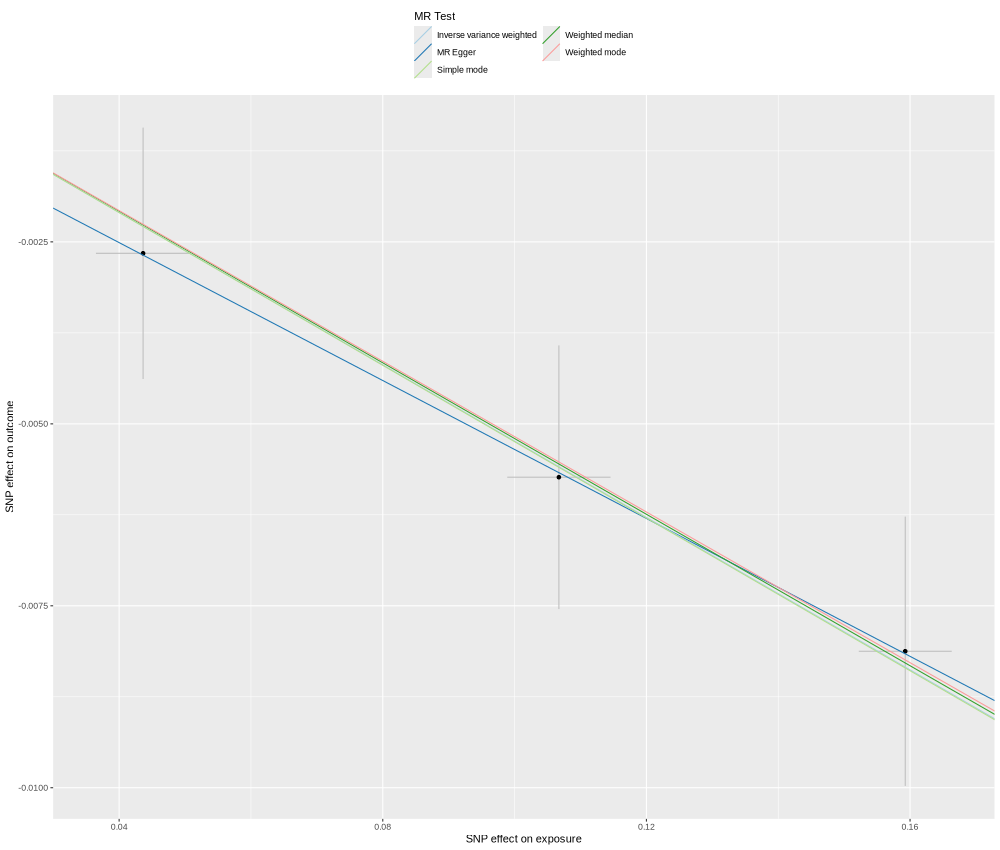

### AMN_Q9BXJ7_OID20492_v1_Inflammation_singleSNP.png

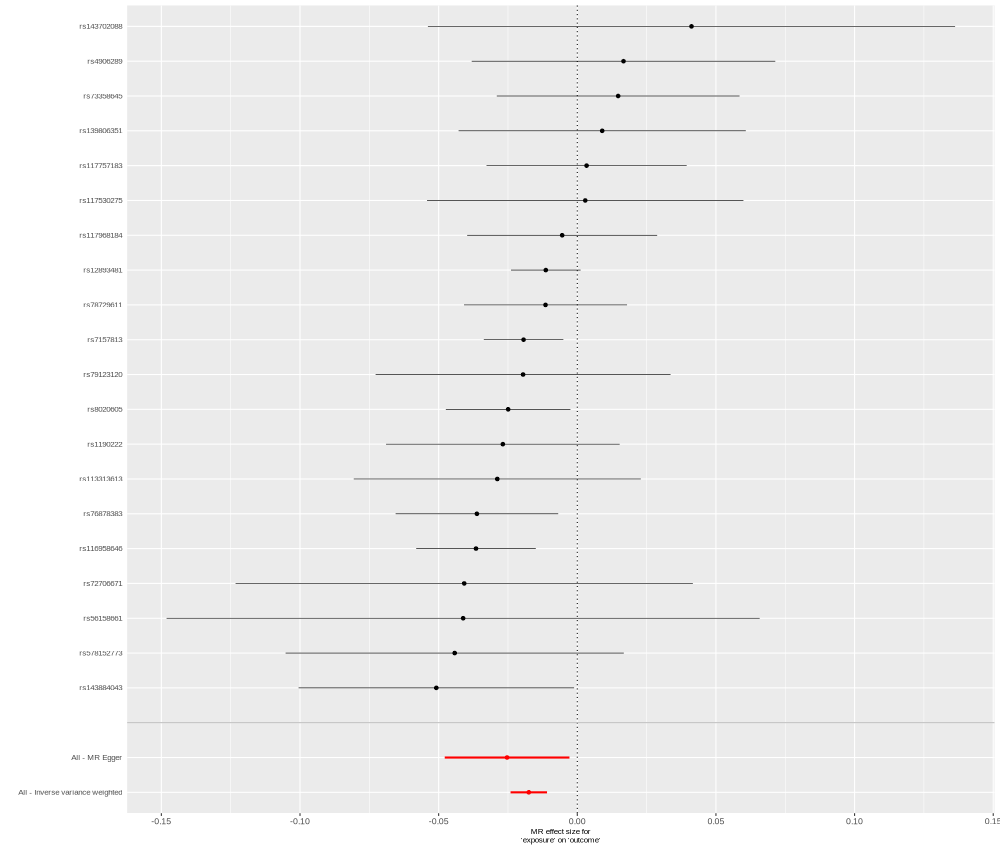

### ANGPTL3_Q9Y5C1_OID20407_v1_Cardiometabolic.png

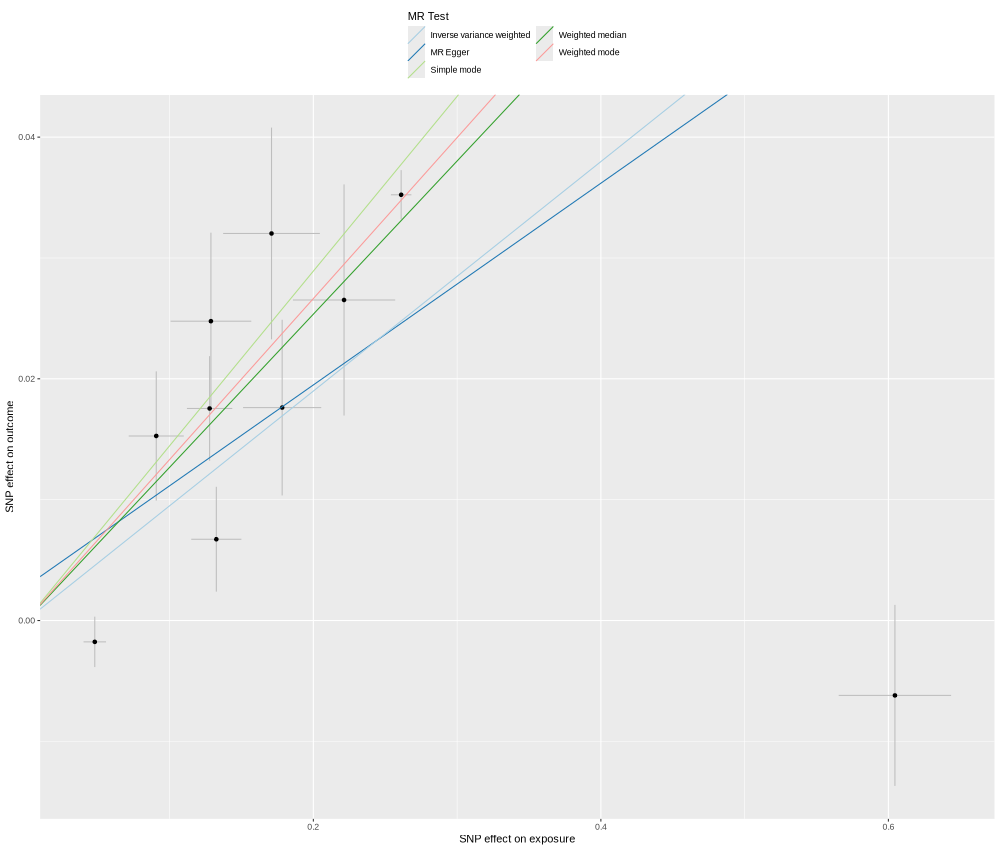

### ANGPTL3_Q9Y5C1_OID20407_v1_Cardiometabolic_singleSNP.png

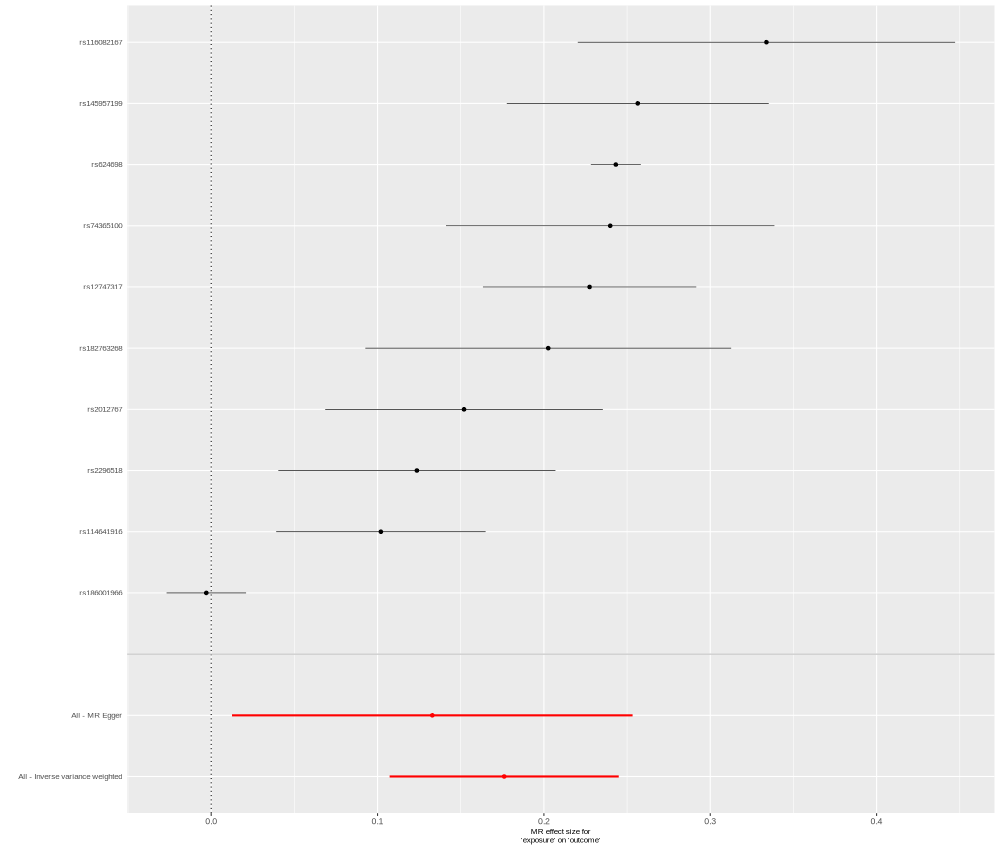

### APOC1_P02654_OID30749_v1_Inflammation_II_singleSNP.png

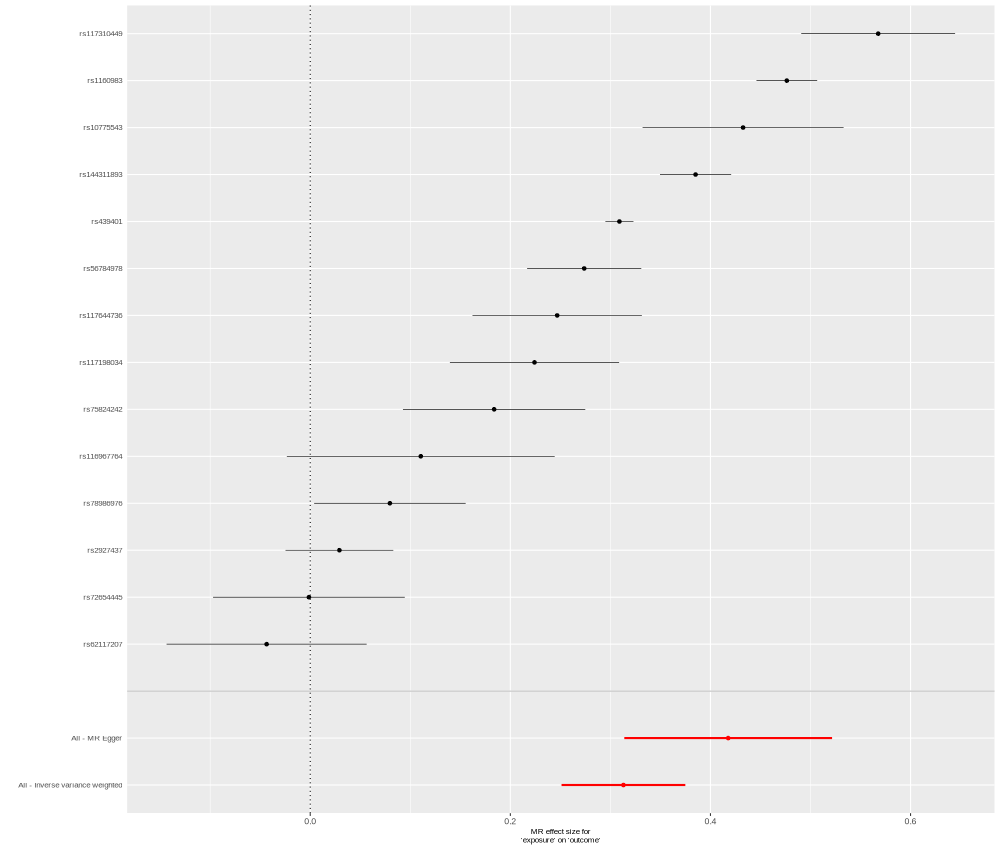

### APOE_P02649_OID30727_v1_Inflammation_II.png

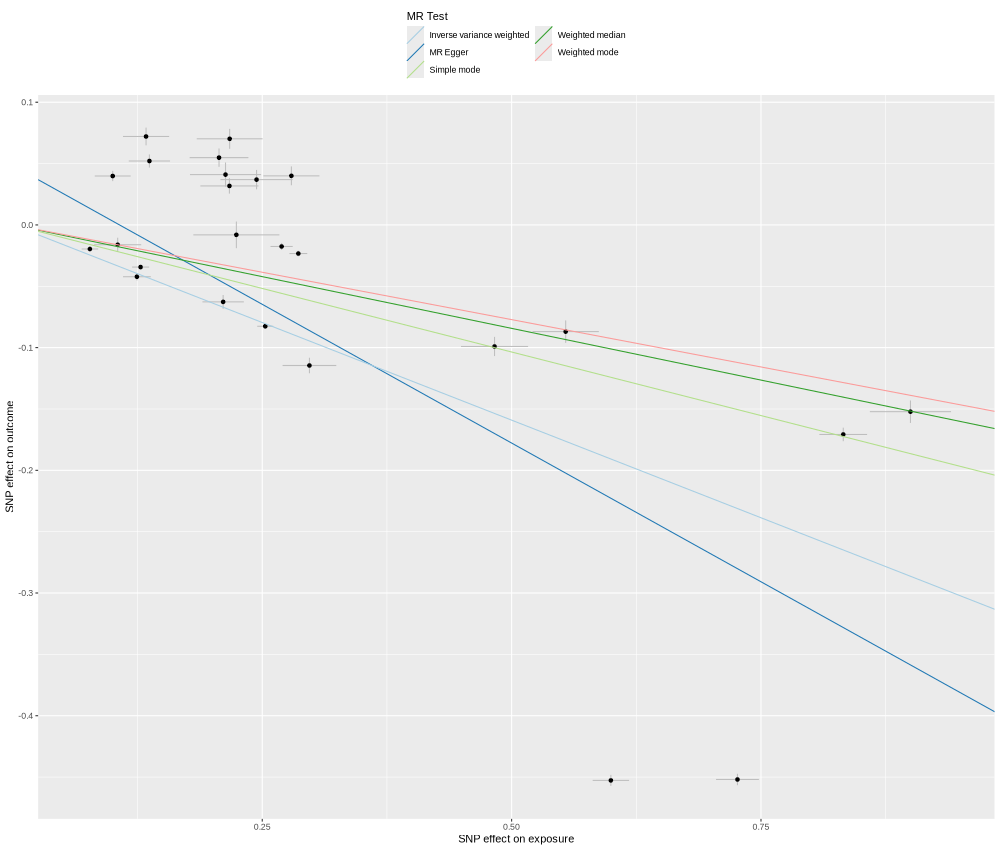

### APOE_P02649_OID30727_v1_Inflammation_II_singleSNP.png

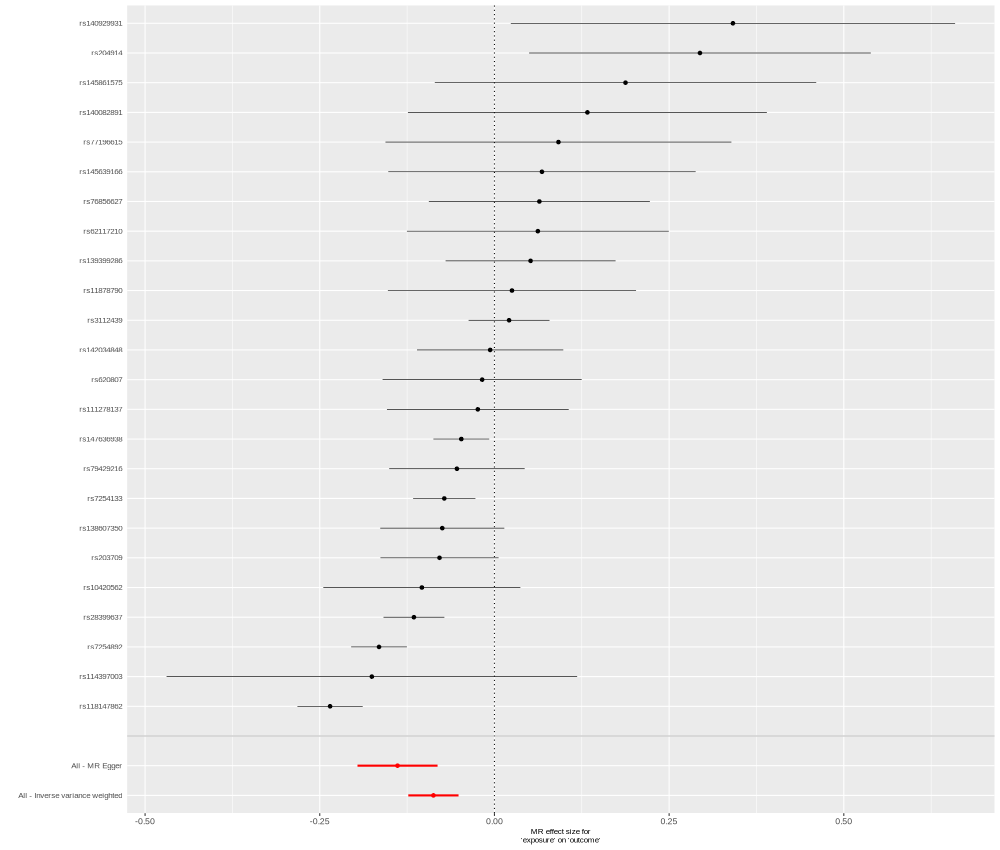

### APOH_P02749_OID21072_v1_Neurology_singleSNP.png

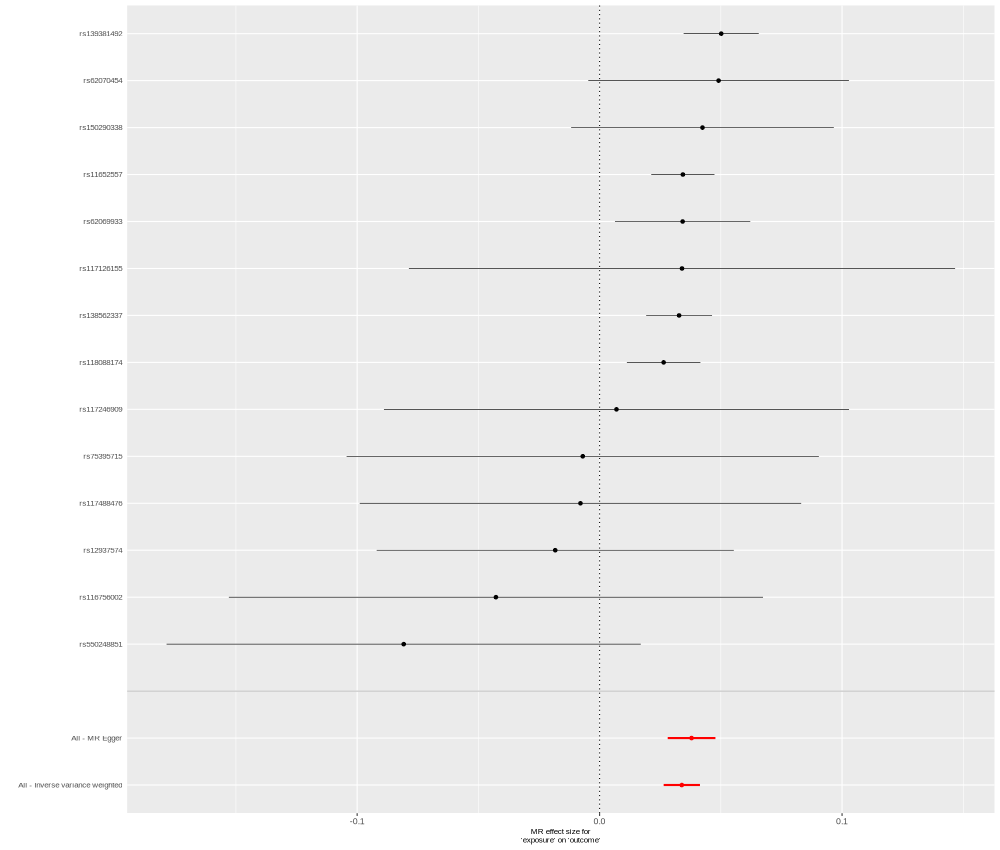

### ASGR1_P07306_OID20990_v1_Neurology.png

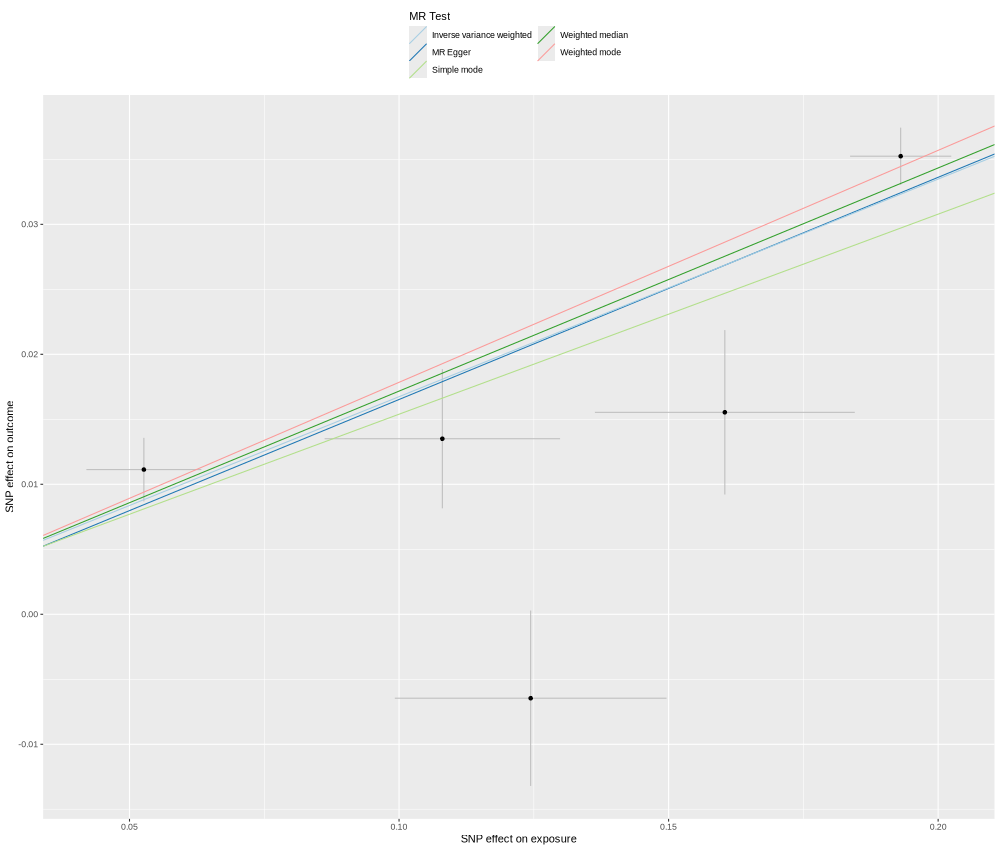

### ASGR1_P07306_OID20990_v1_Neurology_singleSNP.png

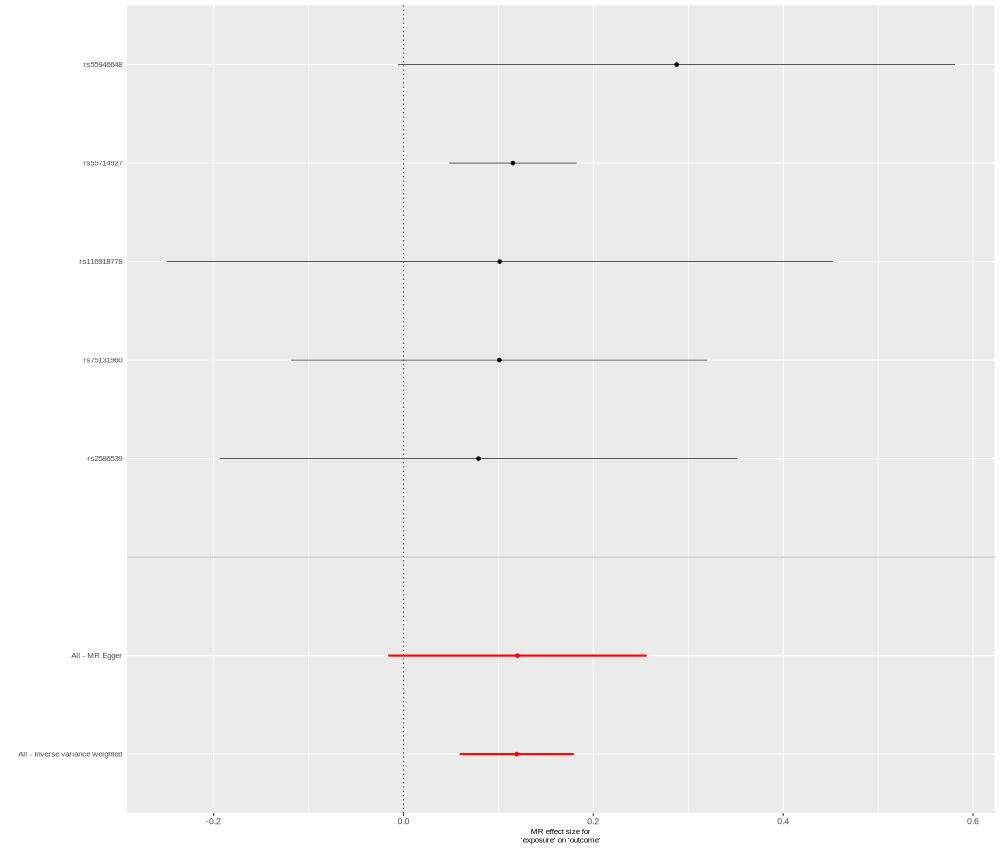

### ASGR1_P07306_OID20990_v1_Neurology_singleSNP.png

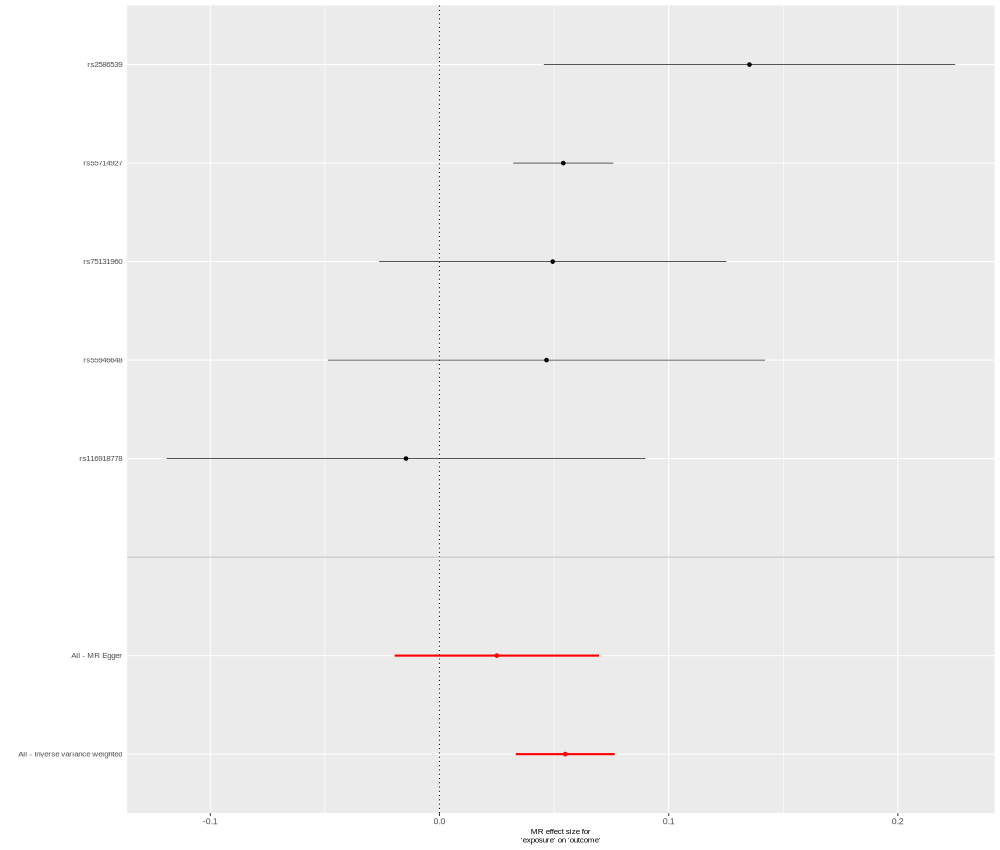

### AXL_P30530_OID20363_v1_Cardiometabolic_singleSNP.png

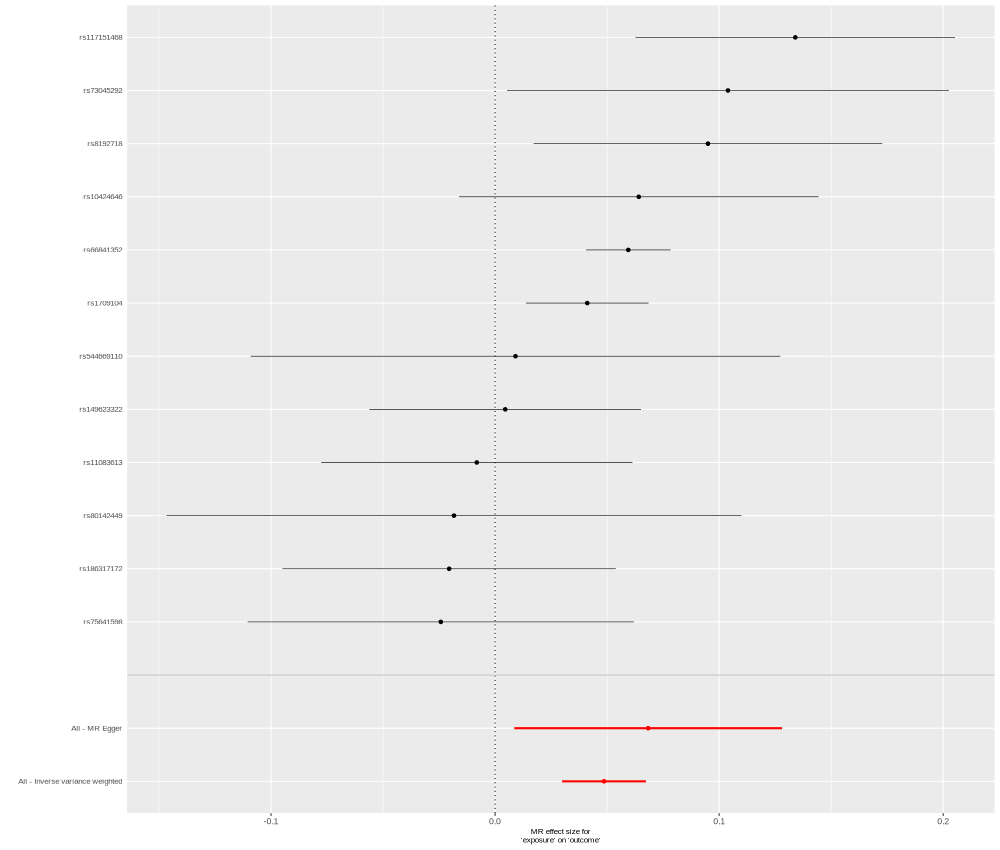

### BLMH_Q13867_OID20336_v1_Cardiometabolic.png

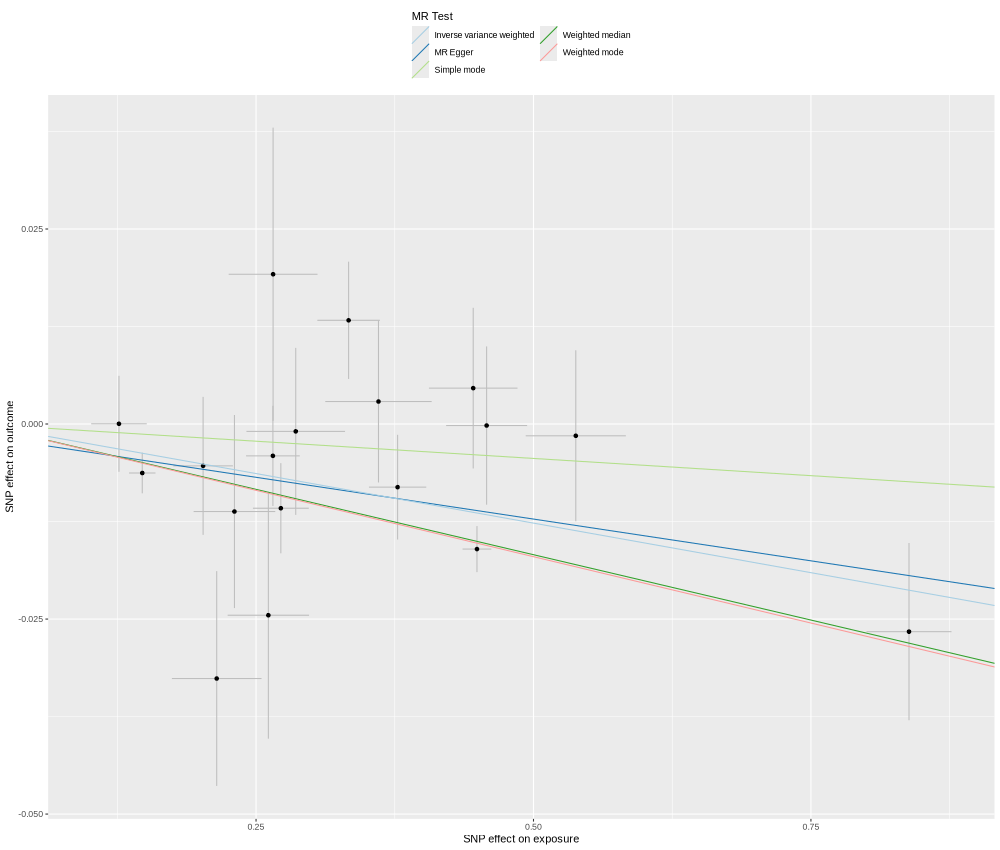

### CASP10_Q92851_OID20893_v1_Neurology_singleSNP.png

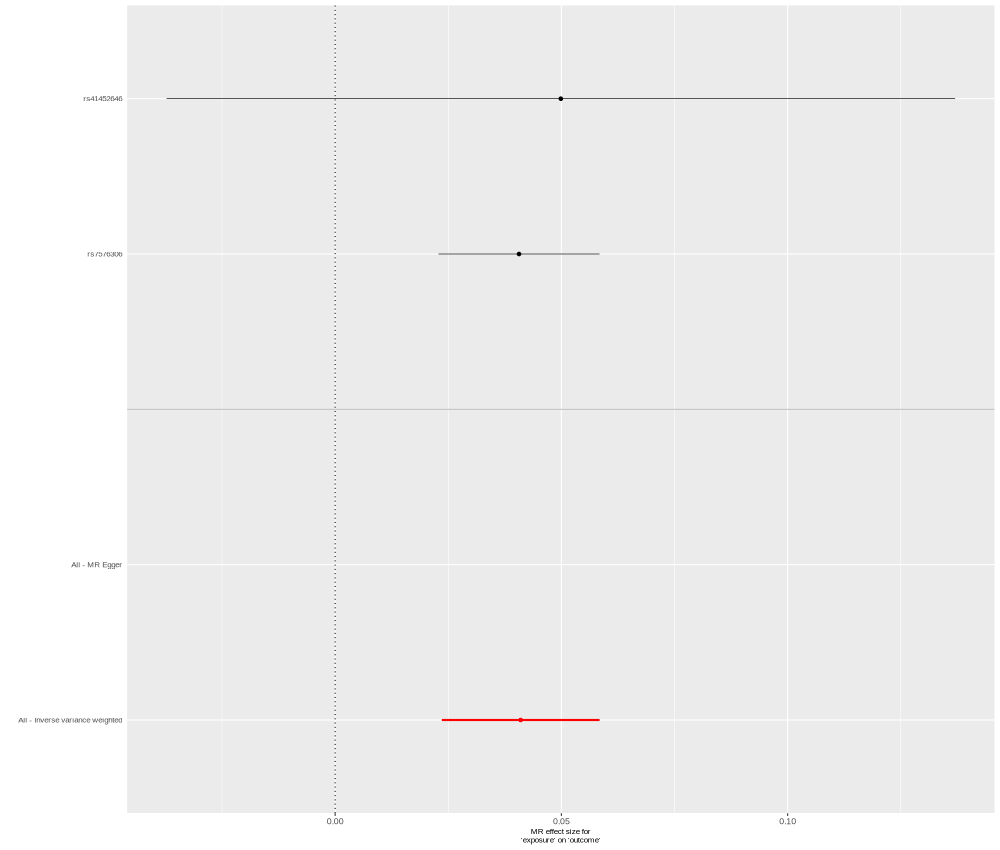

### CD300LG_Q6UXG3_OID21130_v1_Neurology.png

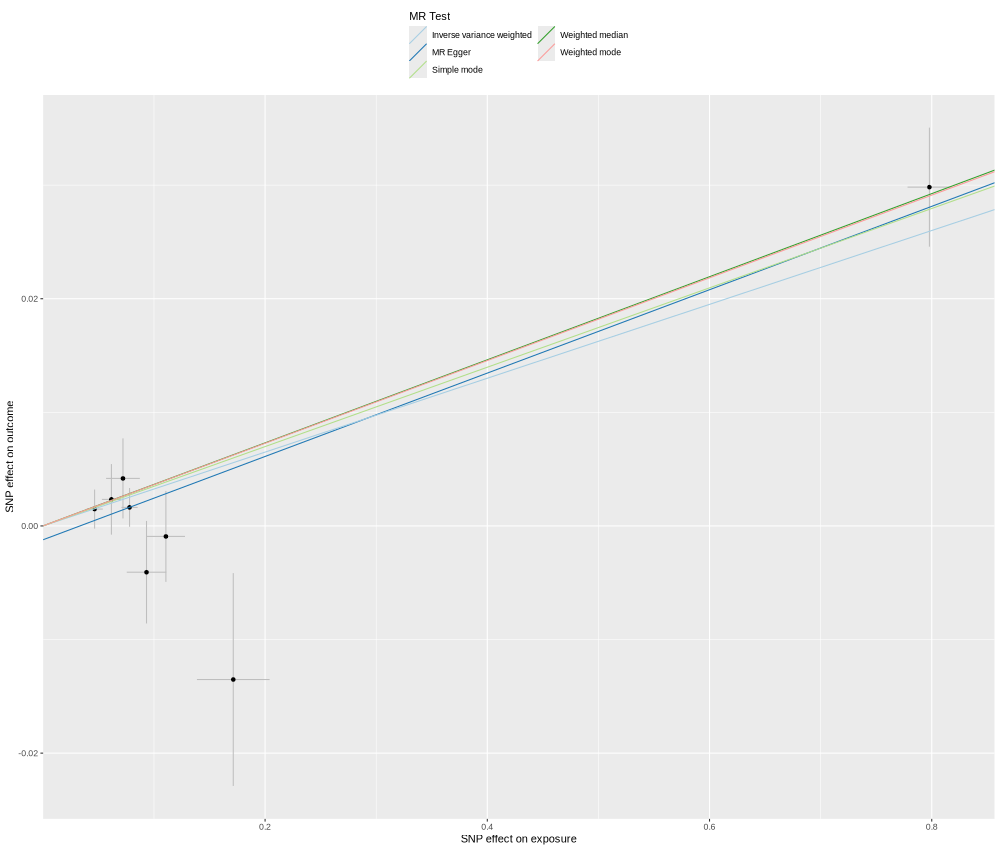

### CD300LG_Q6UXG3_OID21130_v1_Neurology_singleSNP.png

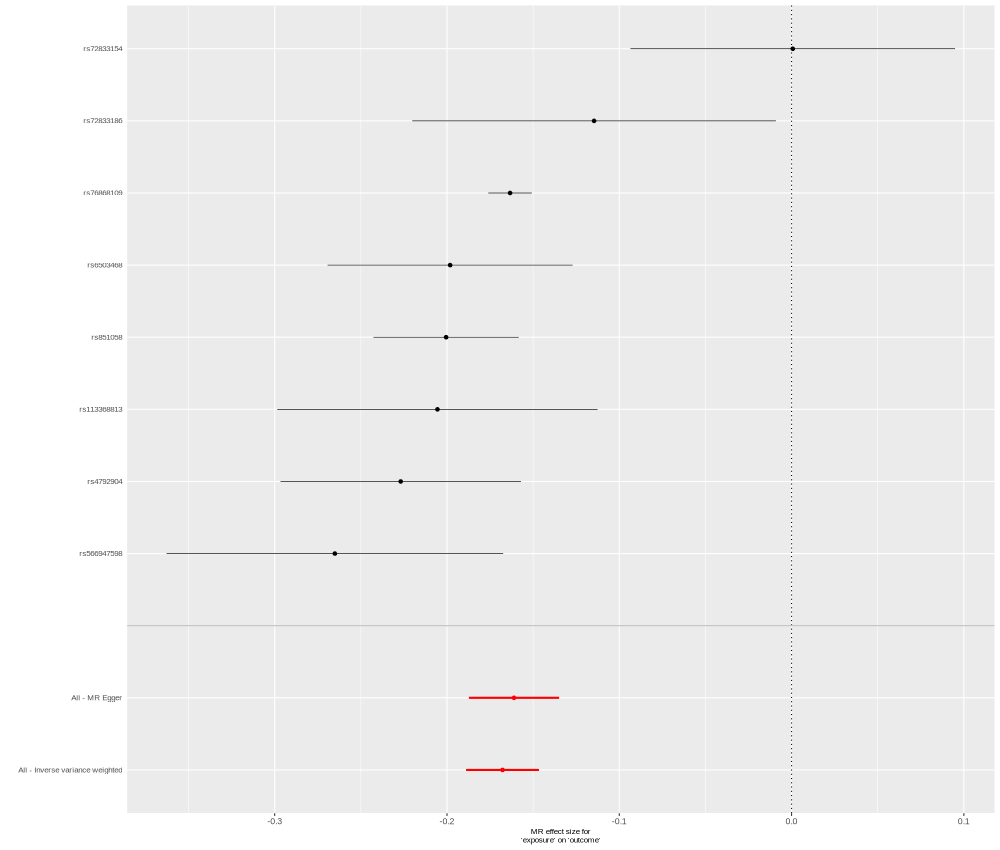

### CELSR2_Q9HCU4_OID30593_v1_Inflammation_II.png

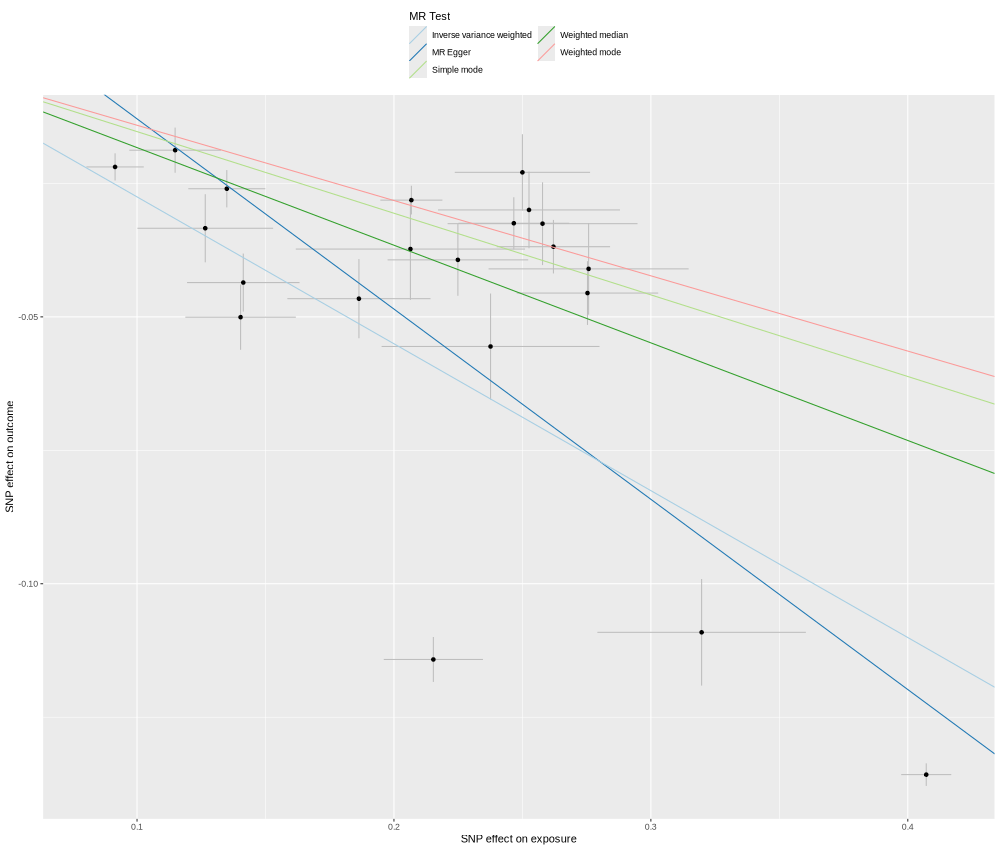

### CELSR2_Q9HCU4_OID30593_v1_Inflammation_II_singleSNP.png

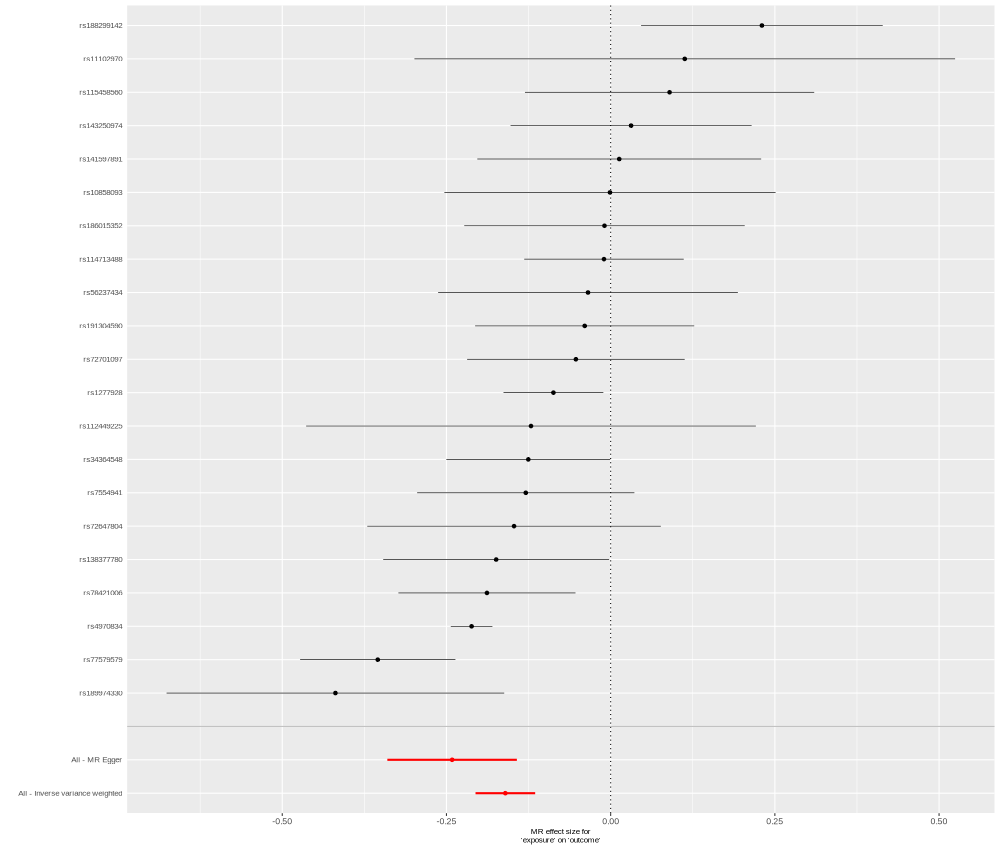

### CELSR2_Q9HCU4_OID30593_v1_Inflammation_II_singleSNP.png

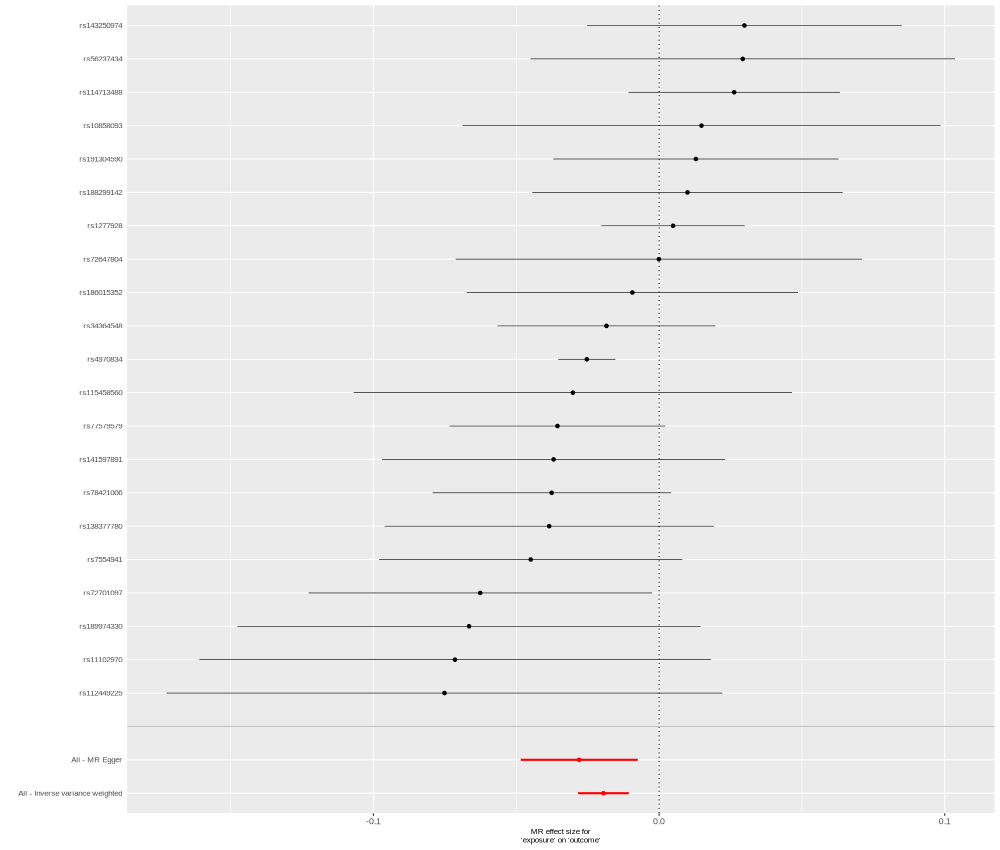

### CFI_P05156_OID30772_v1_Inflammation_II_singleSNP.png

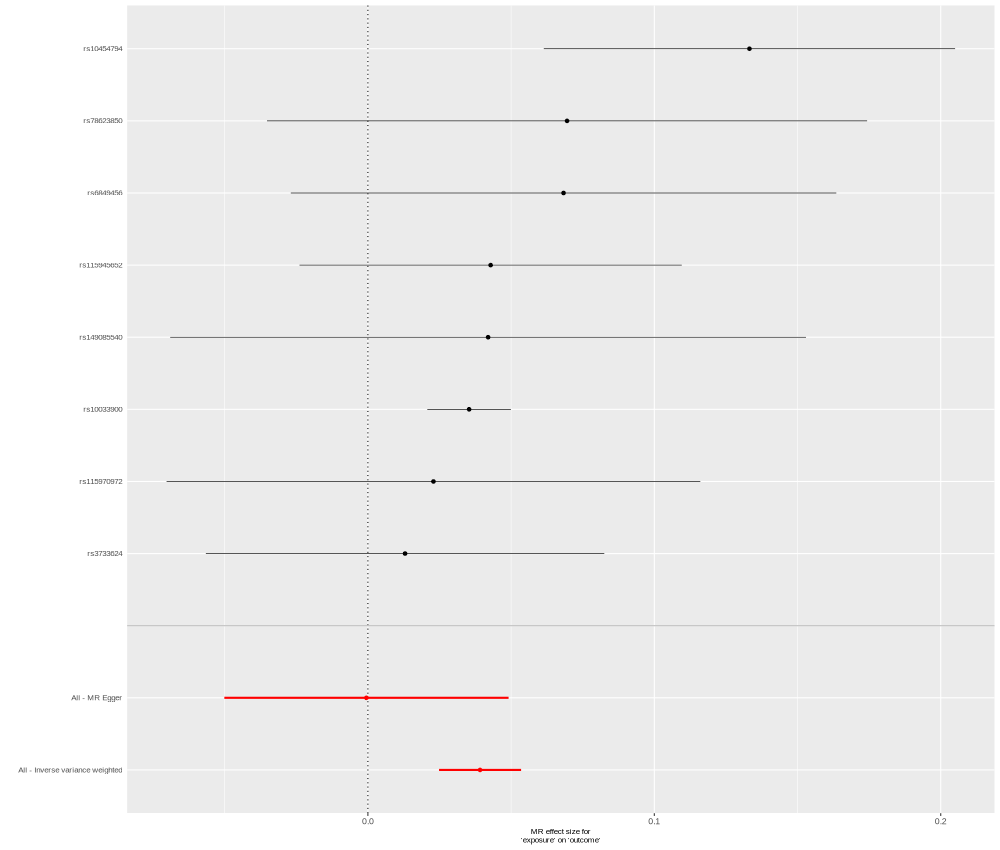

### CLPS_P04118_OID21156_v1_Neurology.png

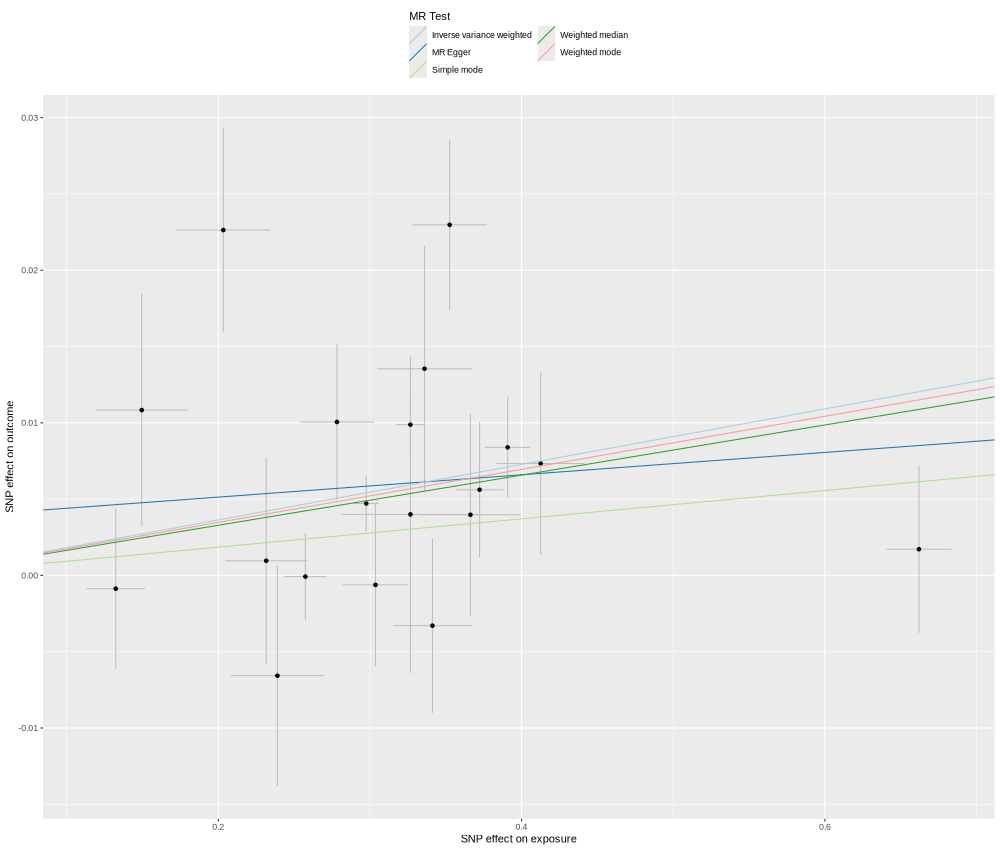

### CRELD2_Q6UXH1_OID20751_v1_Inflammation.png

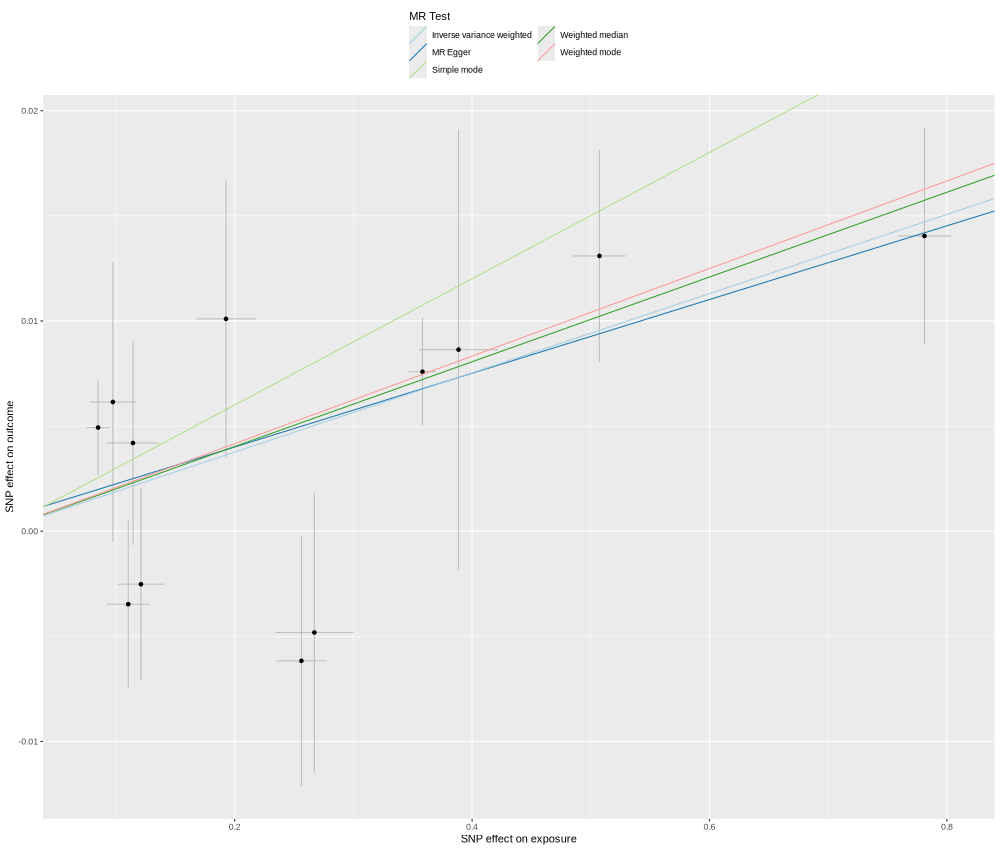

### CRELD2_Q6UXH1_OID20751_v1_Inflammation_singleSNP.png

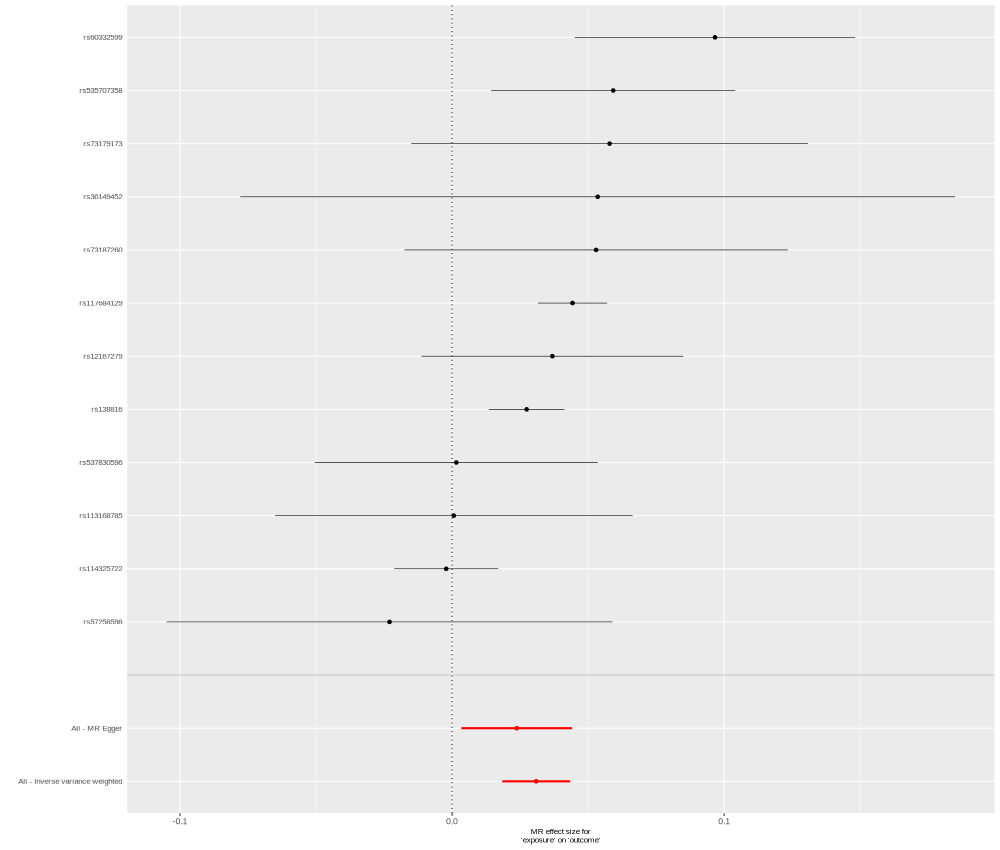

### CXCL16_Q9H2A7_OID20282_v1_Cardiometabolic_singleSNP.png

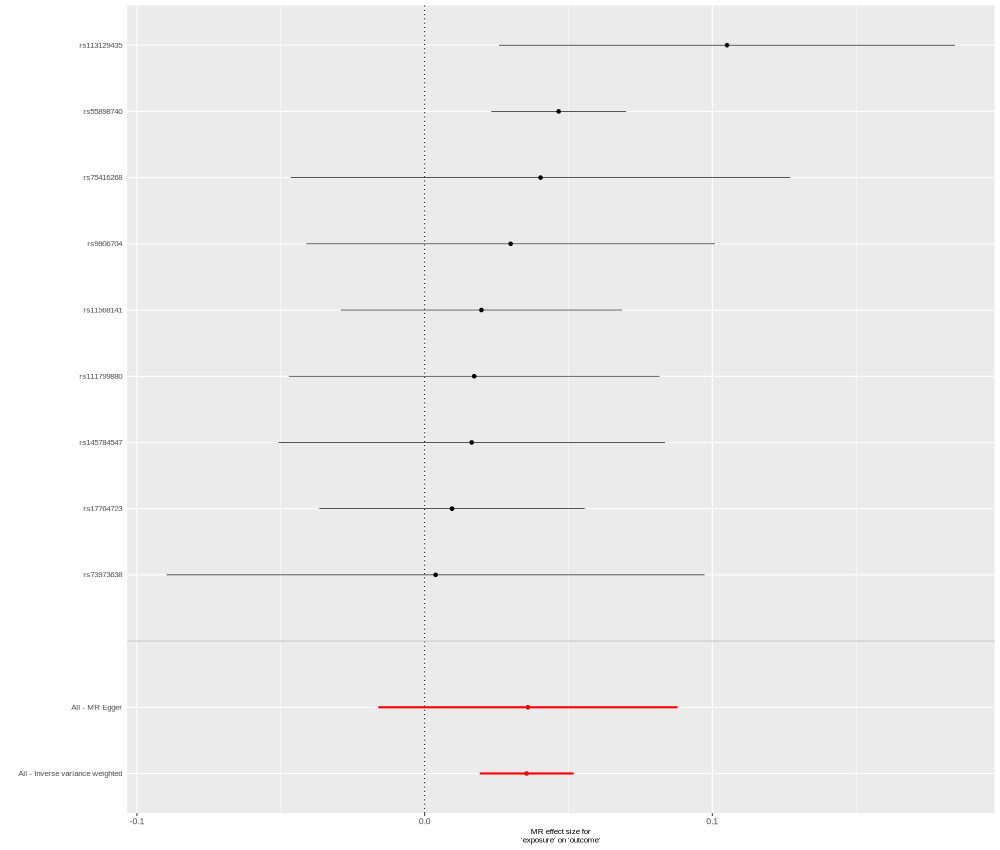

### DDX58_O95786_OID21226_v1_Oncology_singleSNP.png

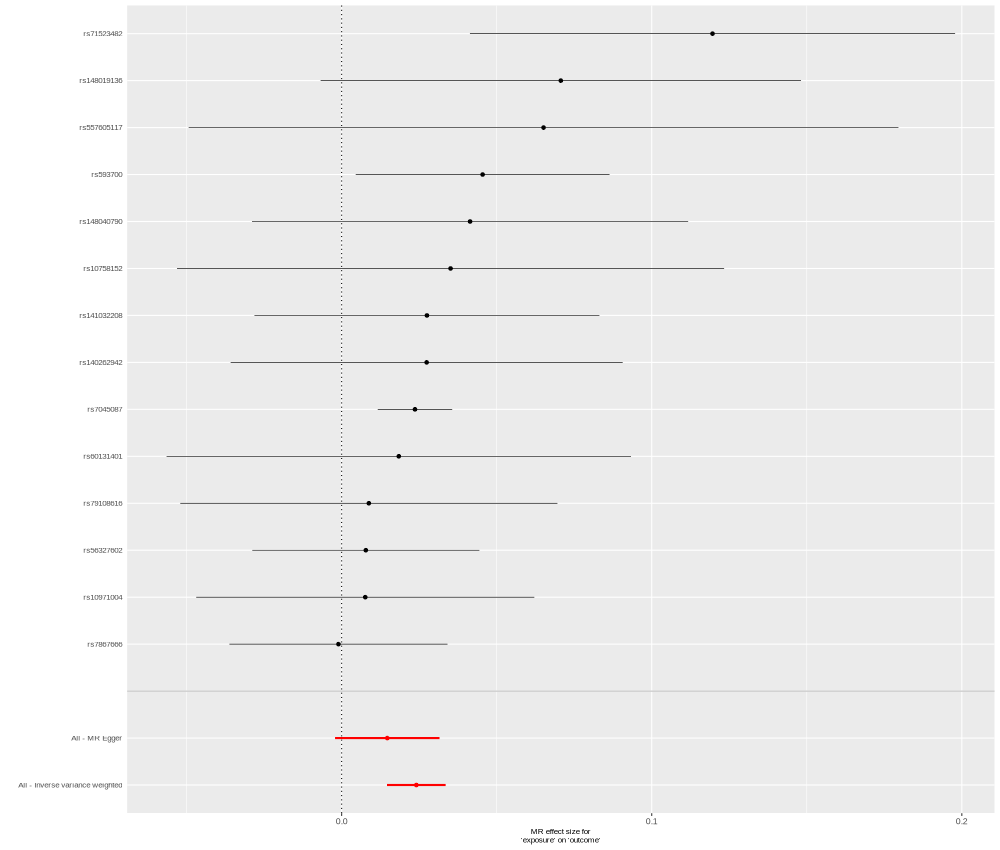

### DUSP13_Q6B8I1_OID31394_v1_Oncology_II_singleSNP.png

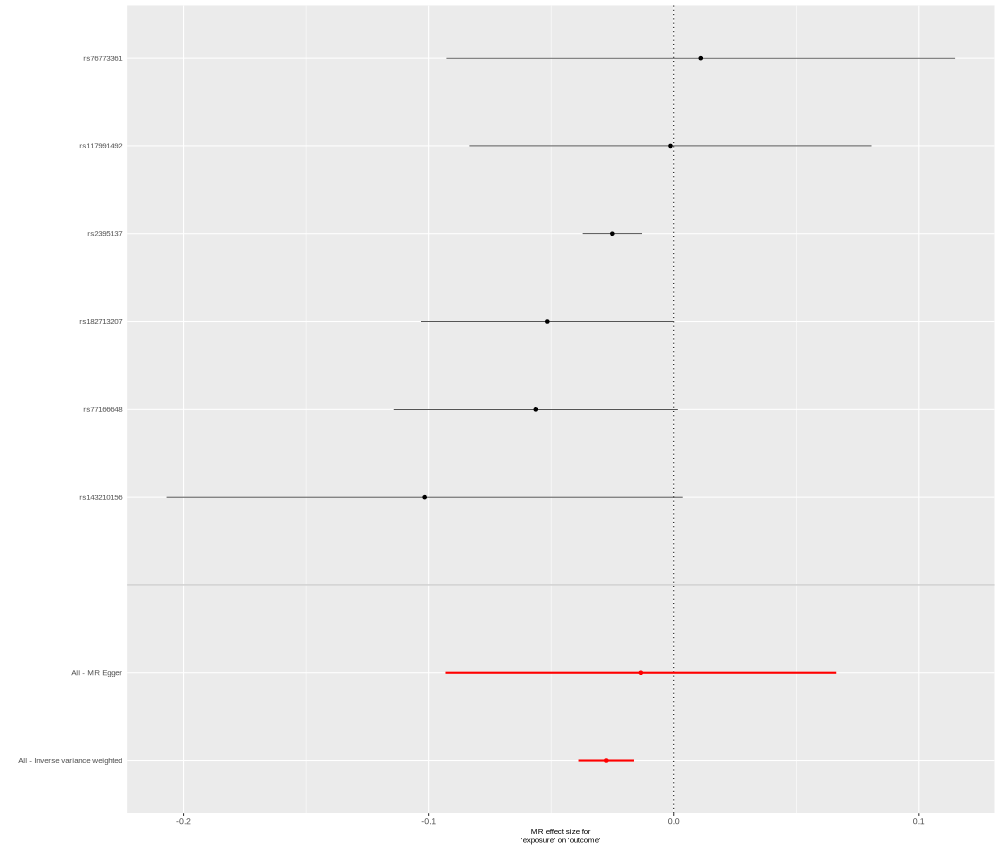

### EFNA1_P20827_OID21125_v1_Neurology.png

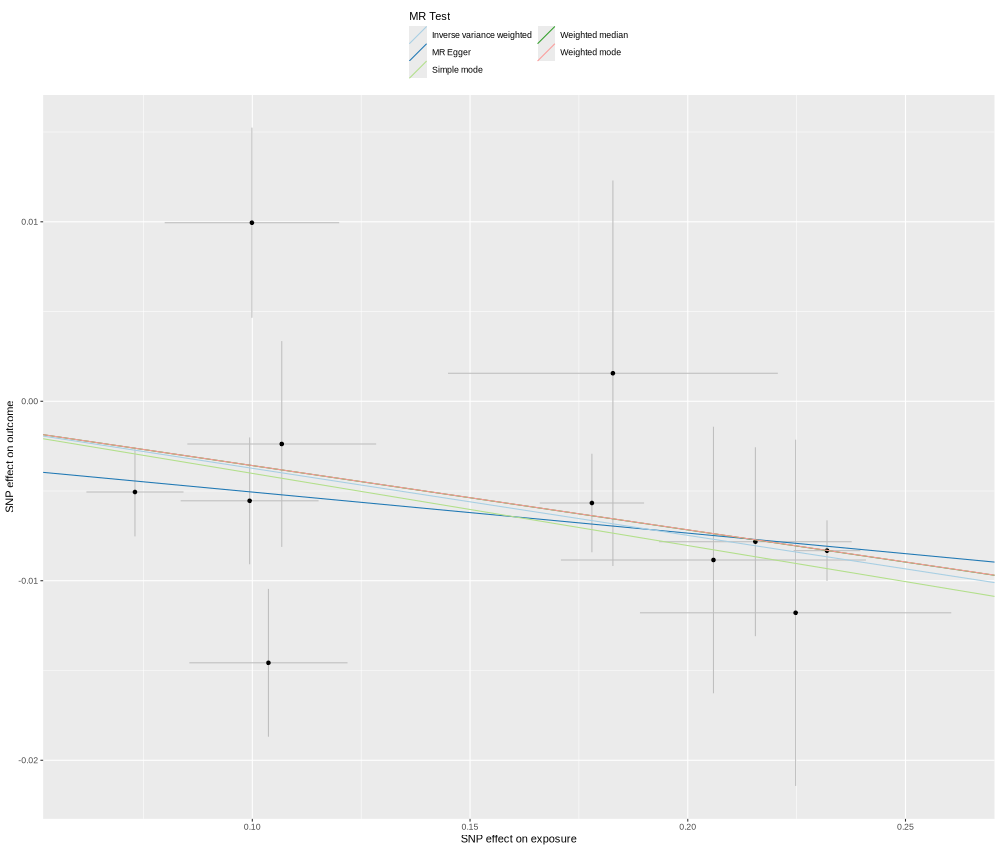

### EGF_P01133_OID20698_v1_Inflammation.png

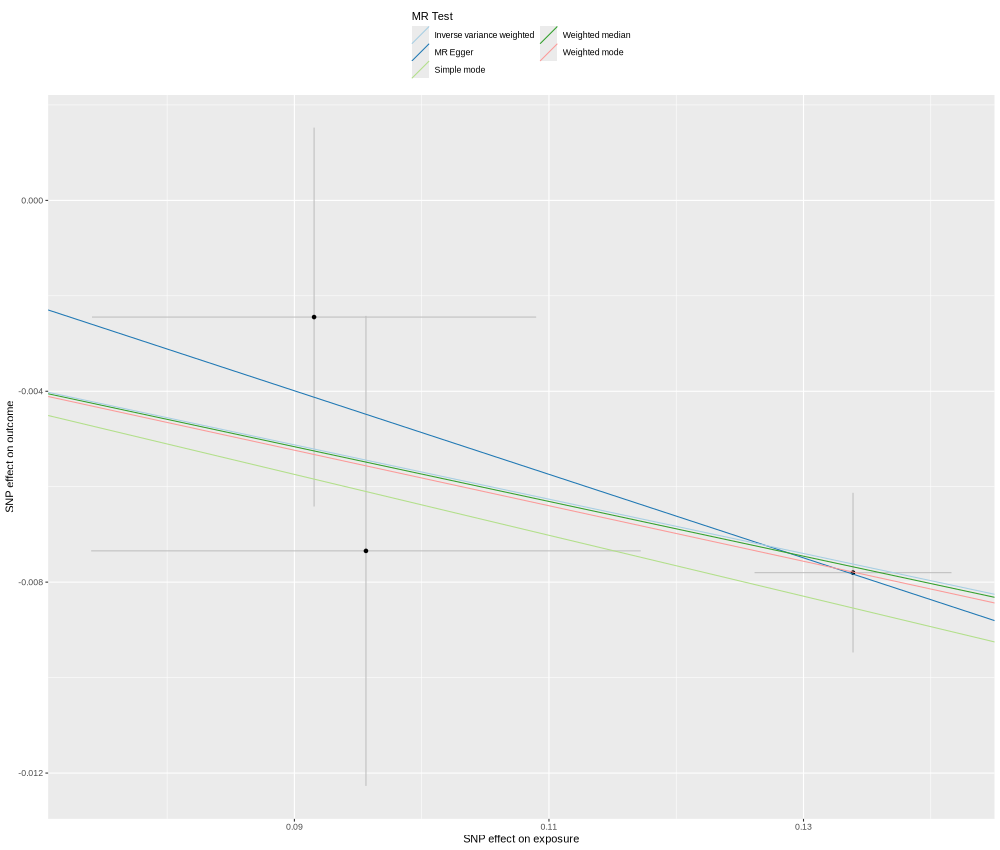

### EXTL1_Q92935_OID30114_v1_Cardiometabolic_II.png

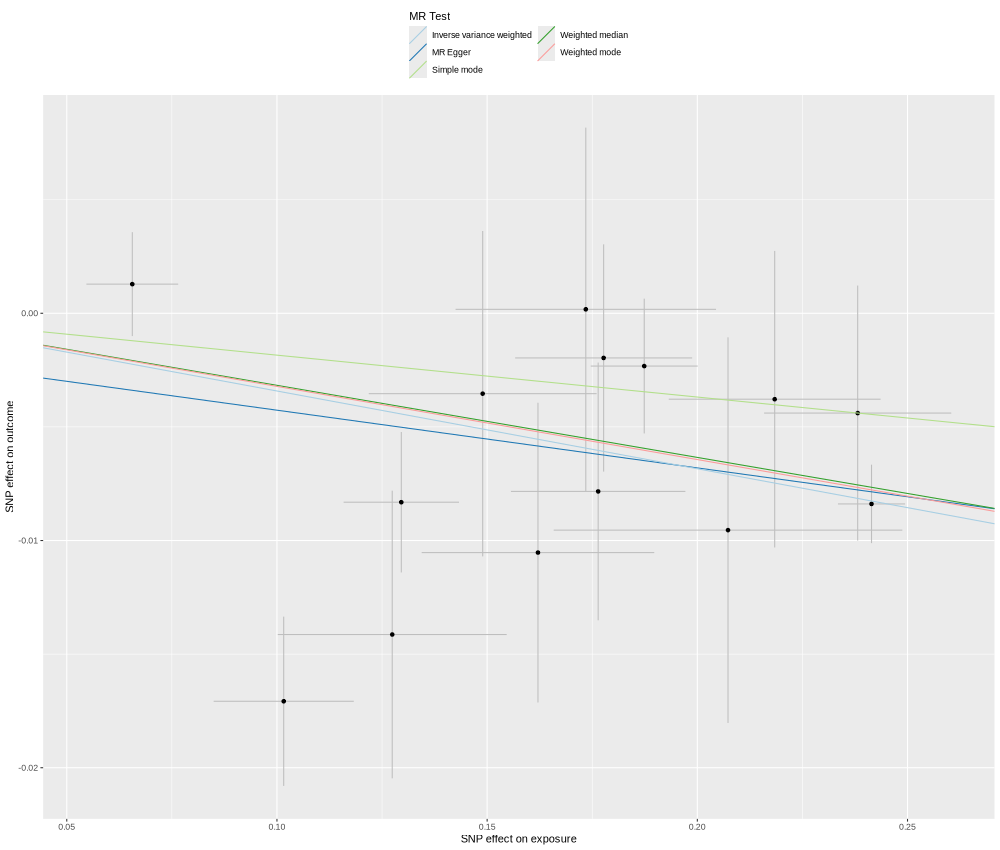

### EXTL1_Q92935_OID30114_v1_Cardiometabolic_II_singleSNP.png

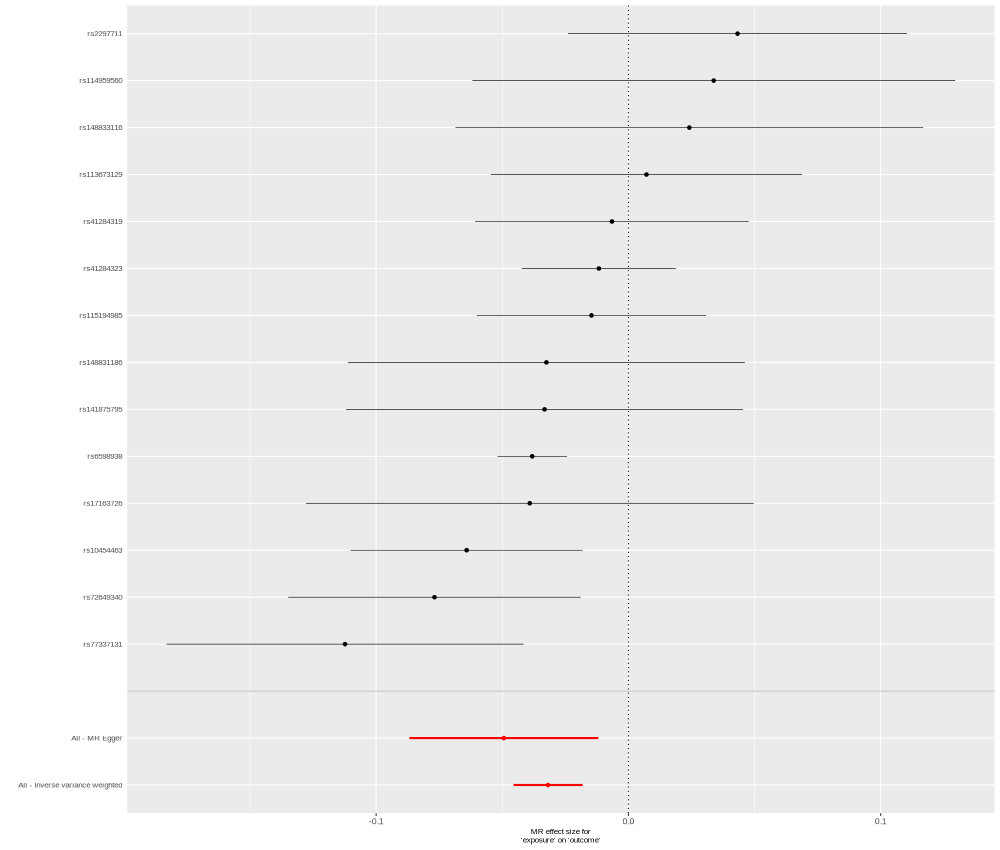

### FN1_P02751_OID30787_v1_Inflammation_II.png

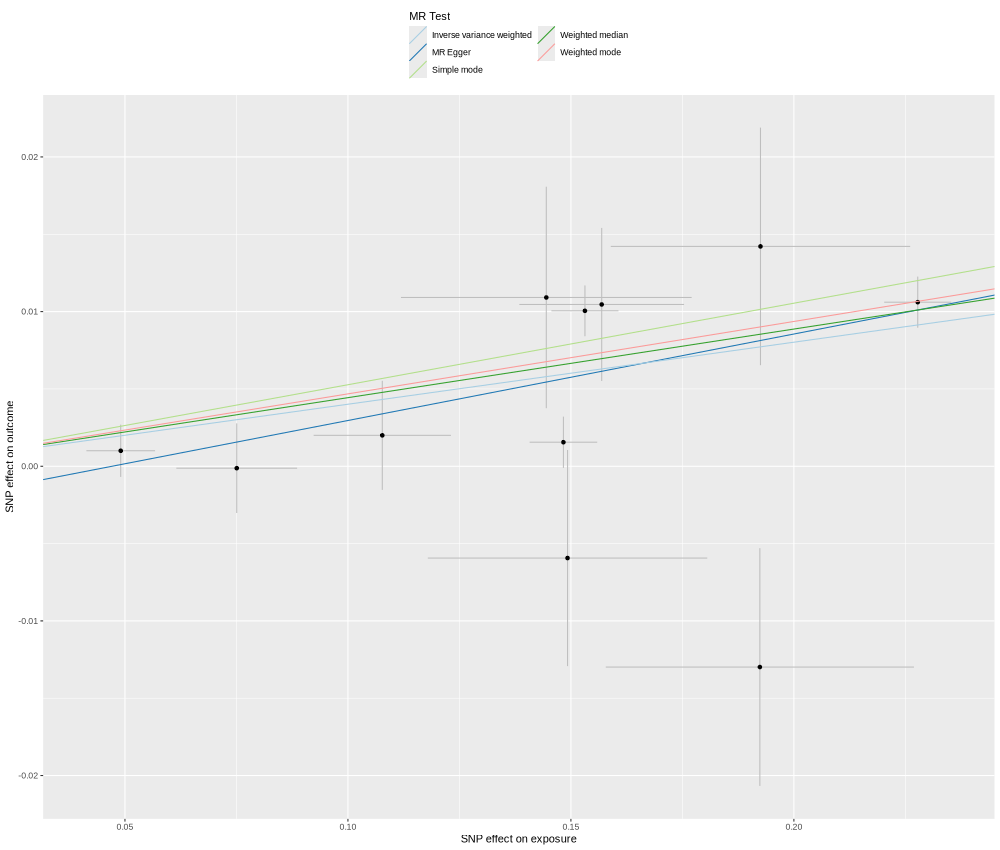

### FN1_P02751_OID30787_v1_Inflammation_II_singleSNP.png

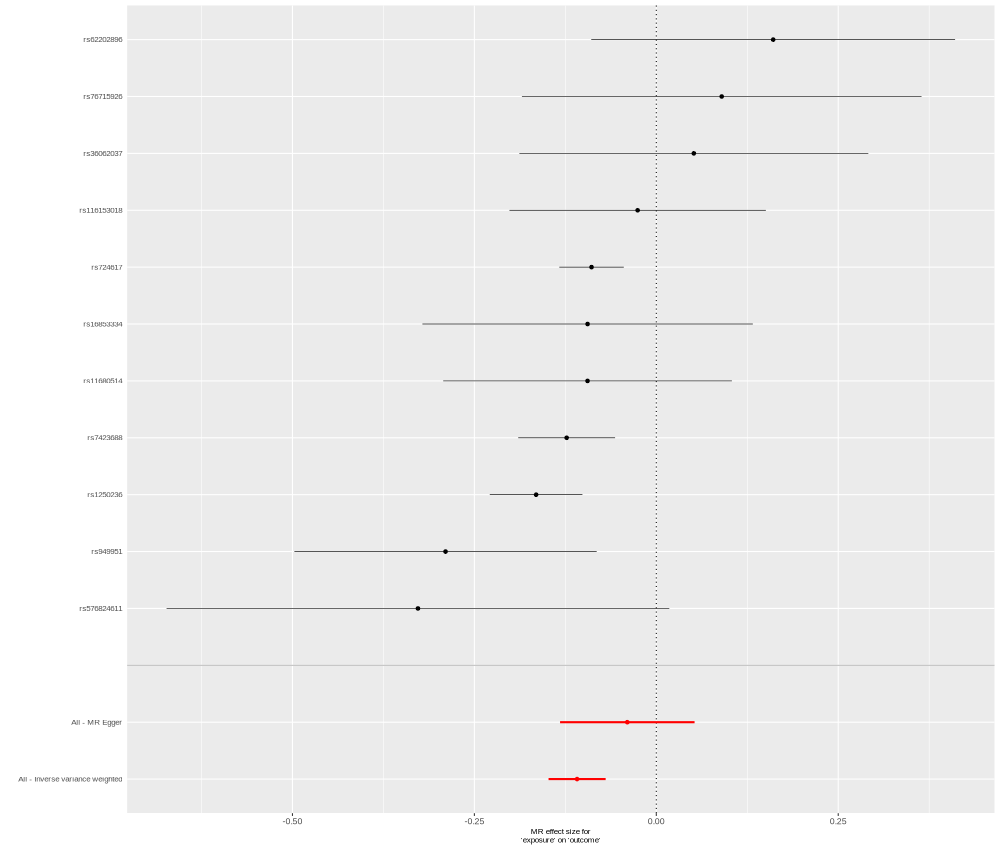

### FSHB_P01225_OID30233_v1_Cardiometabolic_II_singleSNP.png

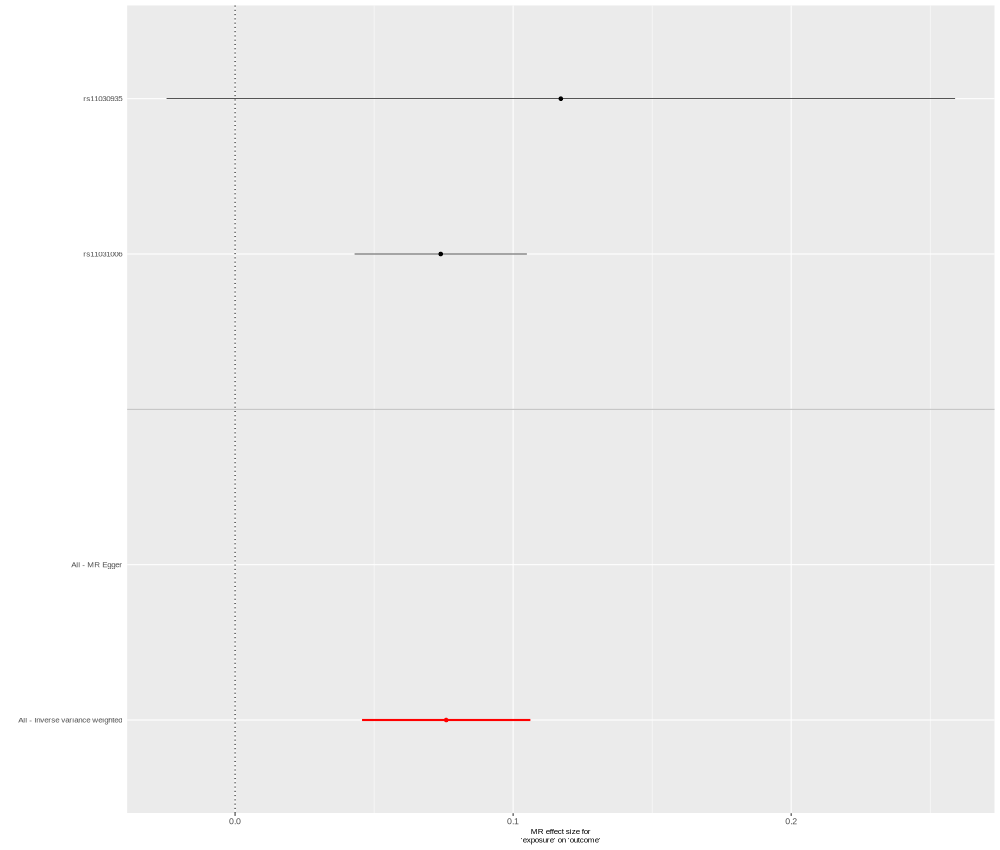

### FSTL3_O95633_OID20782_v1_Inflammation_singleSNP.png

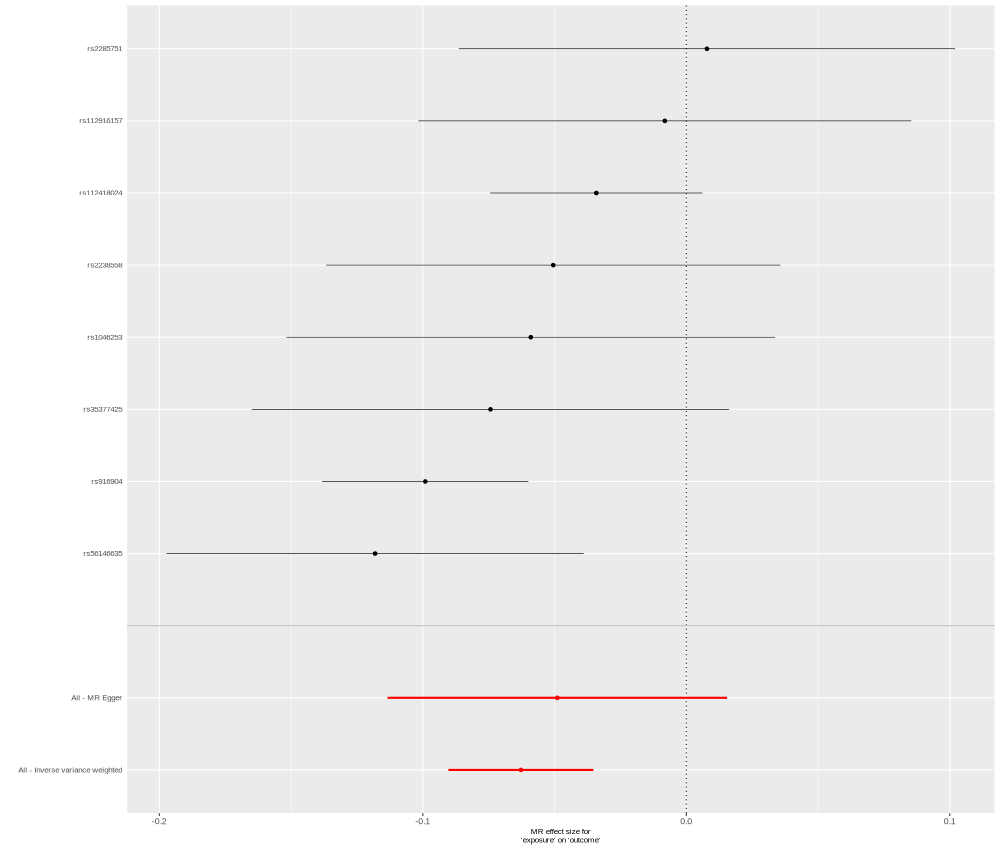

### FURIN_P09958_OID21514_v1_Oncology.png

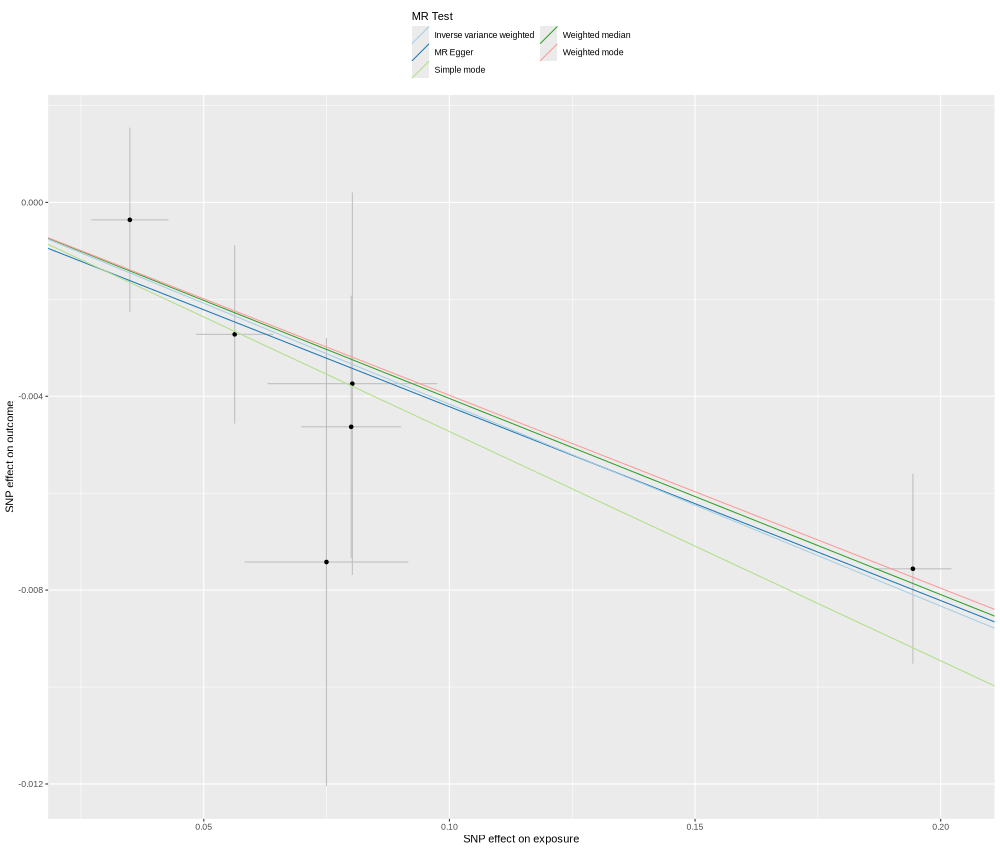

### FURIN_P09958_OID21514_v1_Oncology_singleSNP.png

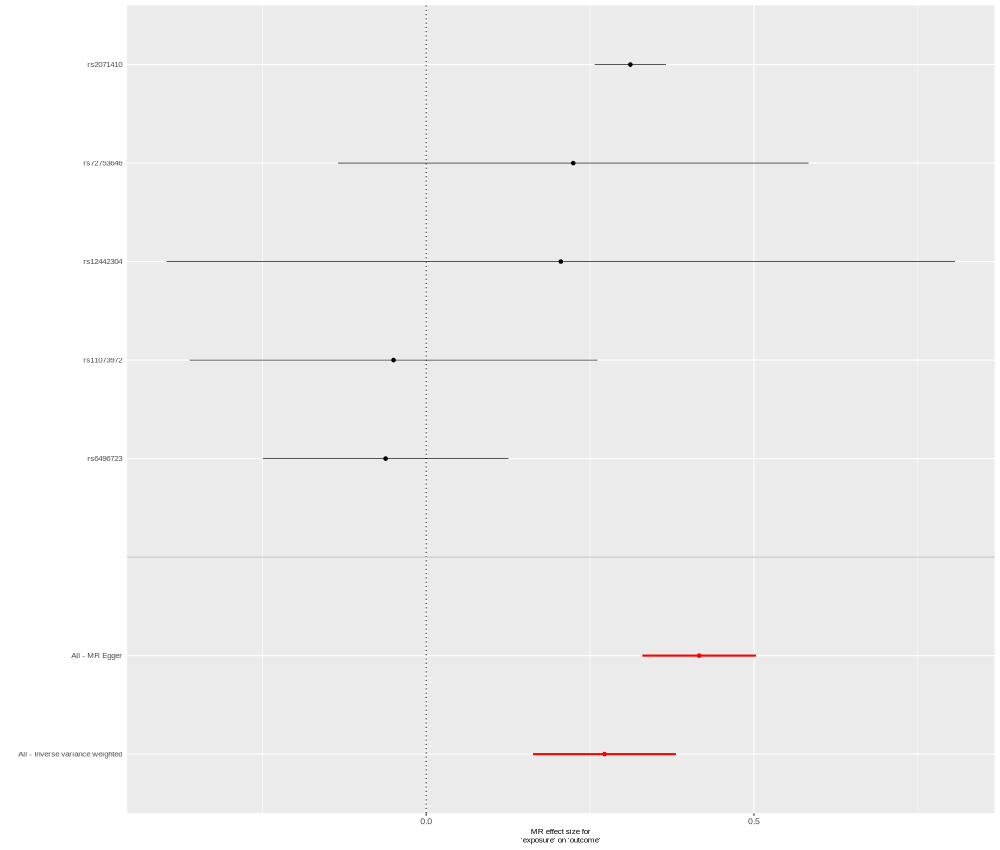

### GALNT2_Q10471_OID21423_v1_Oncology_singleSNP.png

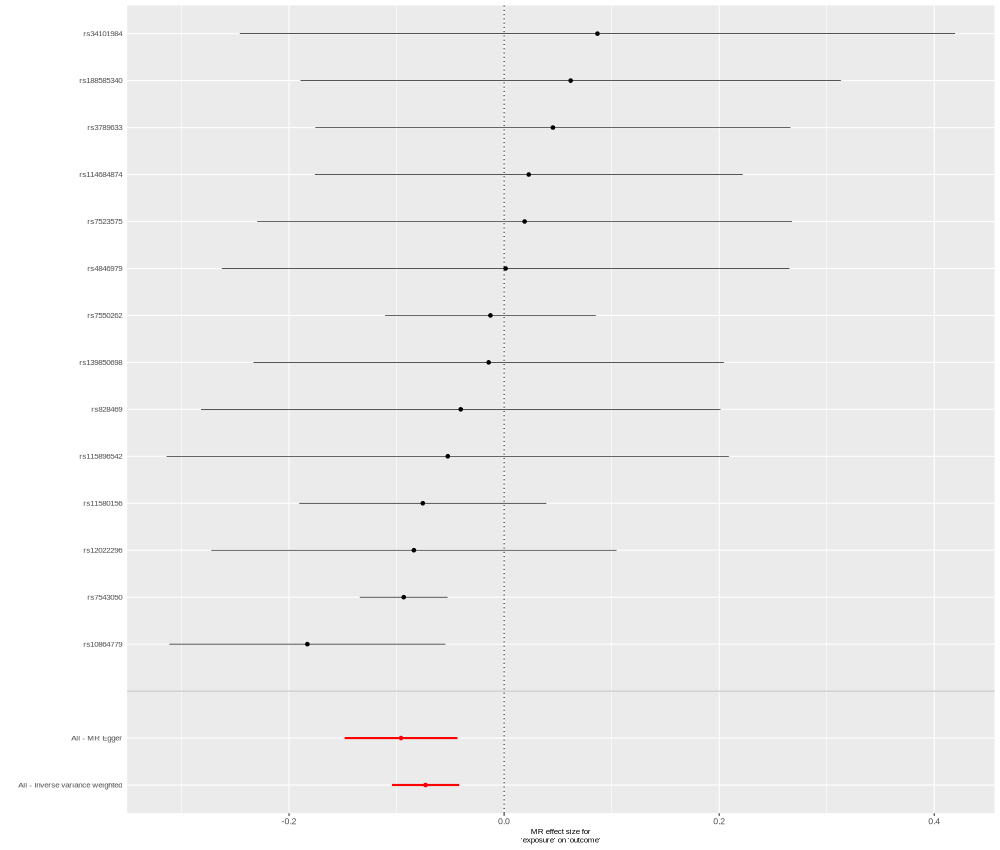

### GALNT2_Q10471_OID21423_v1_Oncology_singleSNP.png

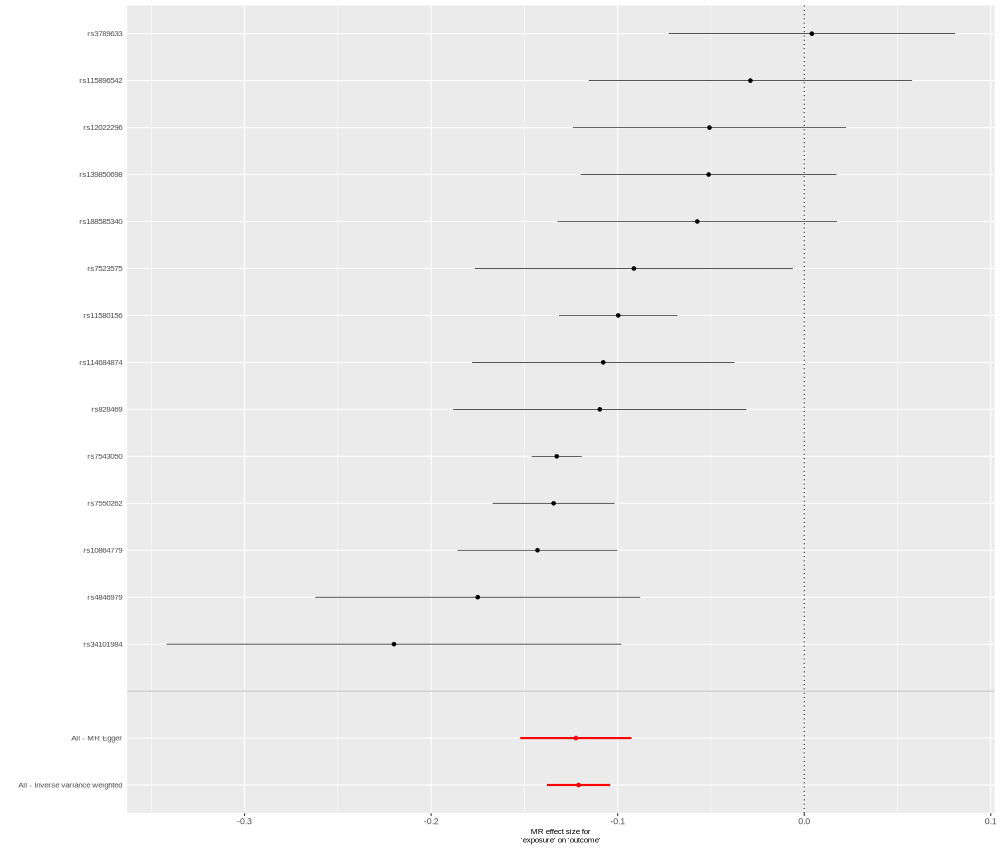

### GIPC2_Q8TF65_OID31153_v1_Neurology_II.png

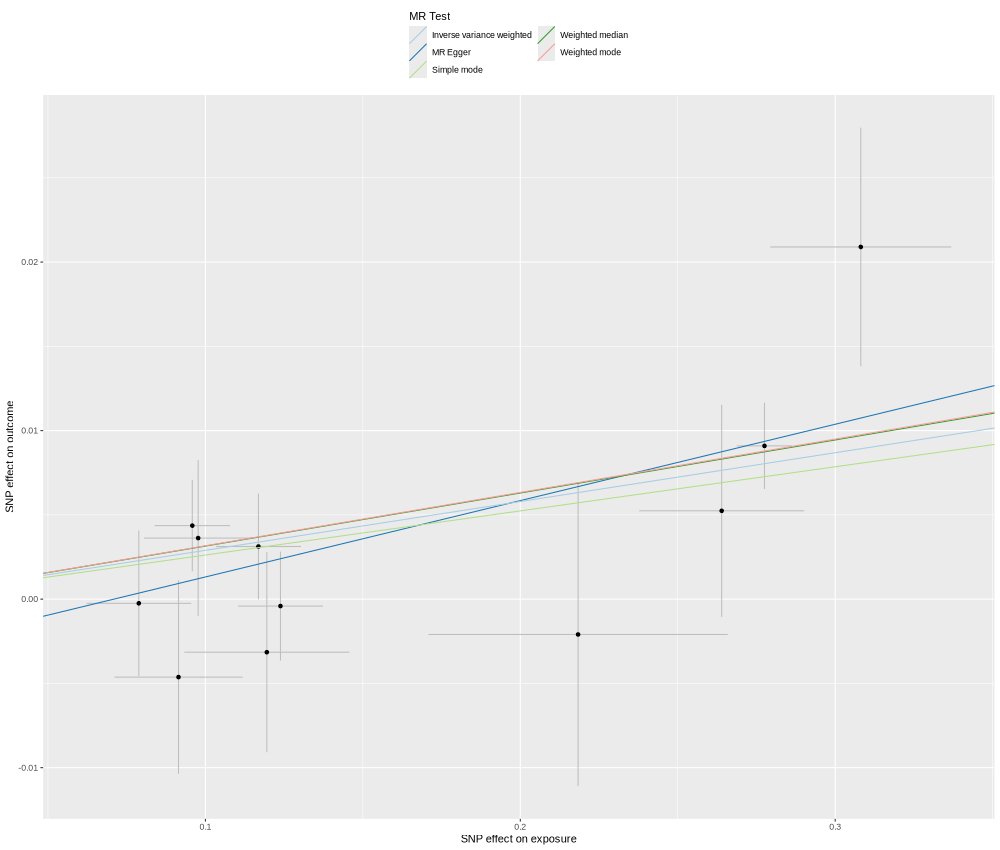

### GSTA1_P08263_OID20166_v1_Cardiometabolic_singleSNP.png

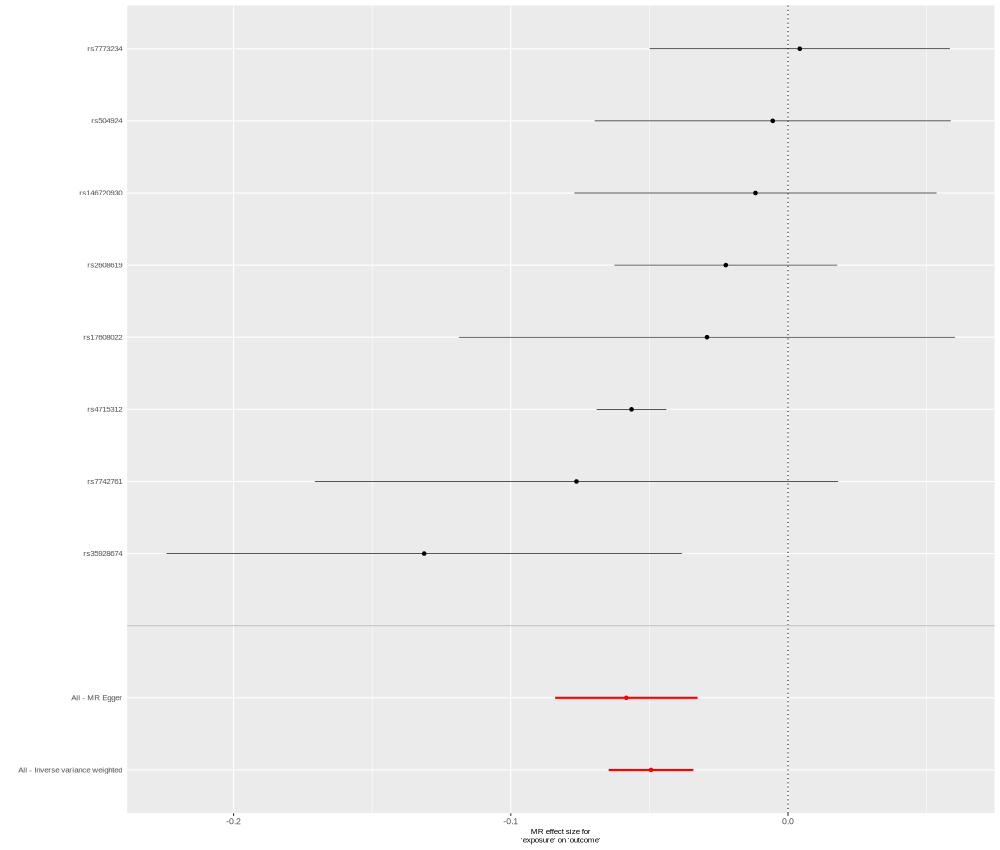

### GSTA3_Q16772_OID21242_v1_Oncology_singleSNP.png

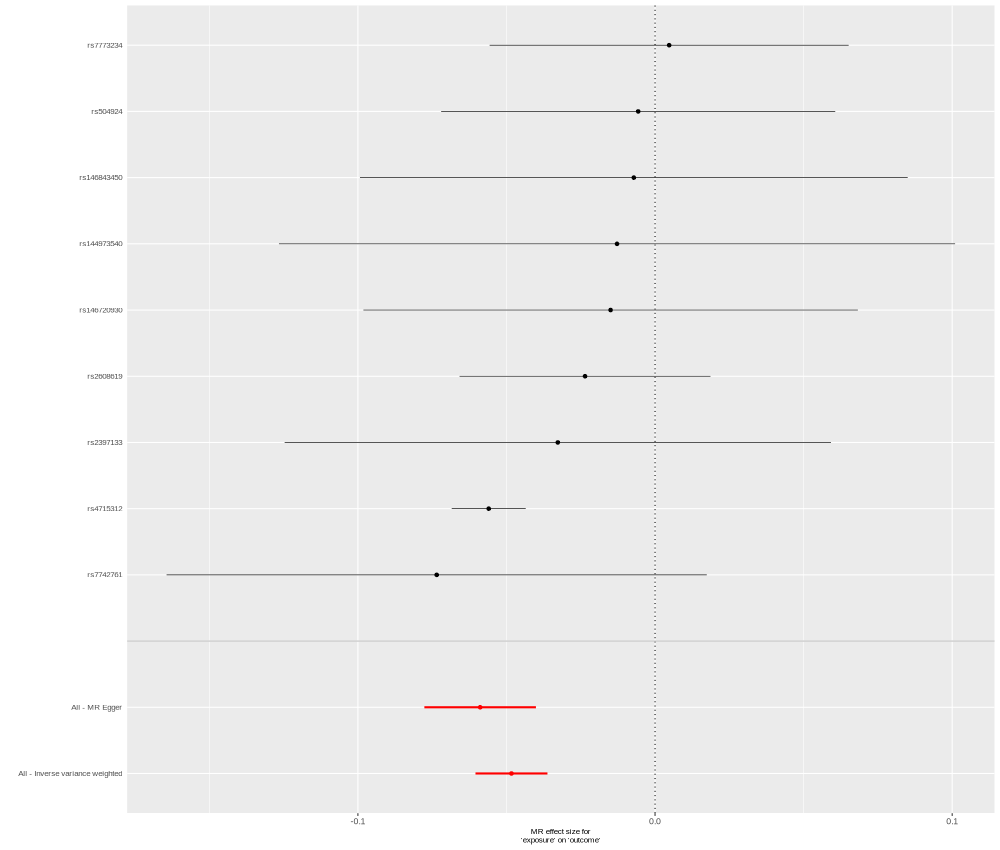

### GSTT2B_P0CG30_OID31097_v1_Neurology_II_singleSNP.png

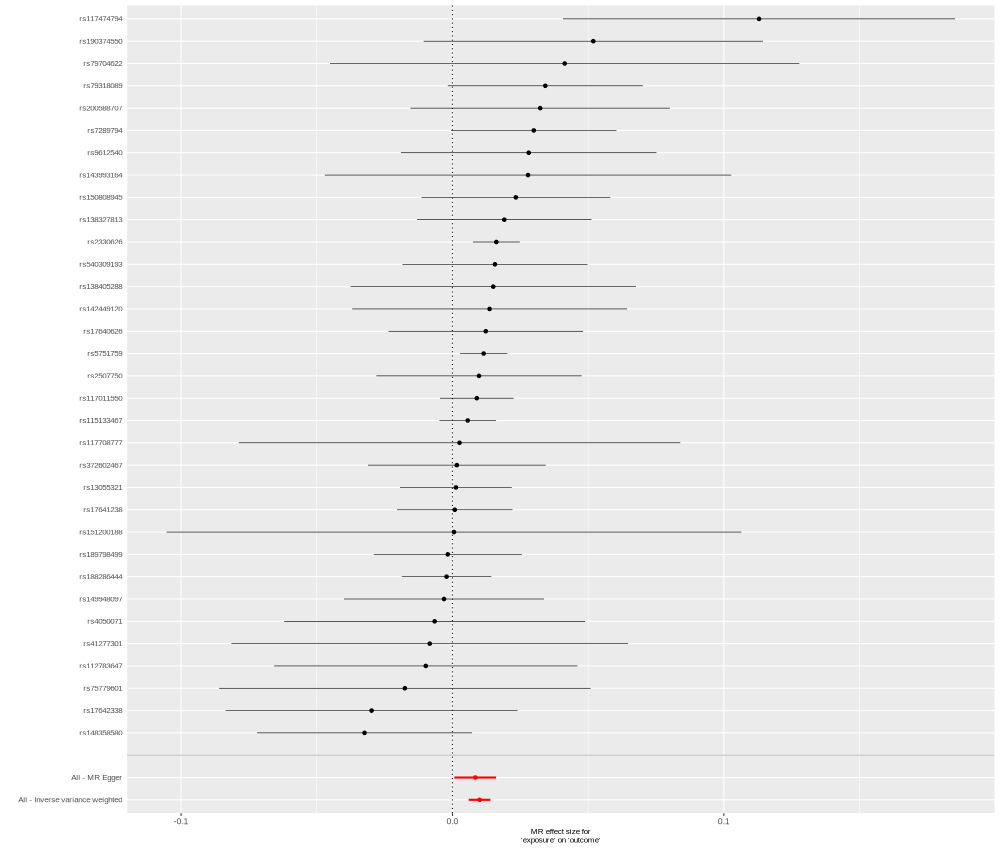

### HEBP1_Q9NRV9_OID20063_v1_Cardiometabolic_singleSNP.png

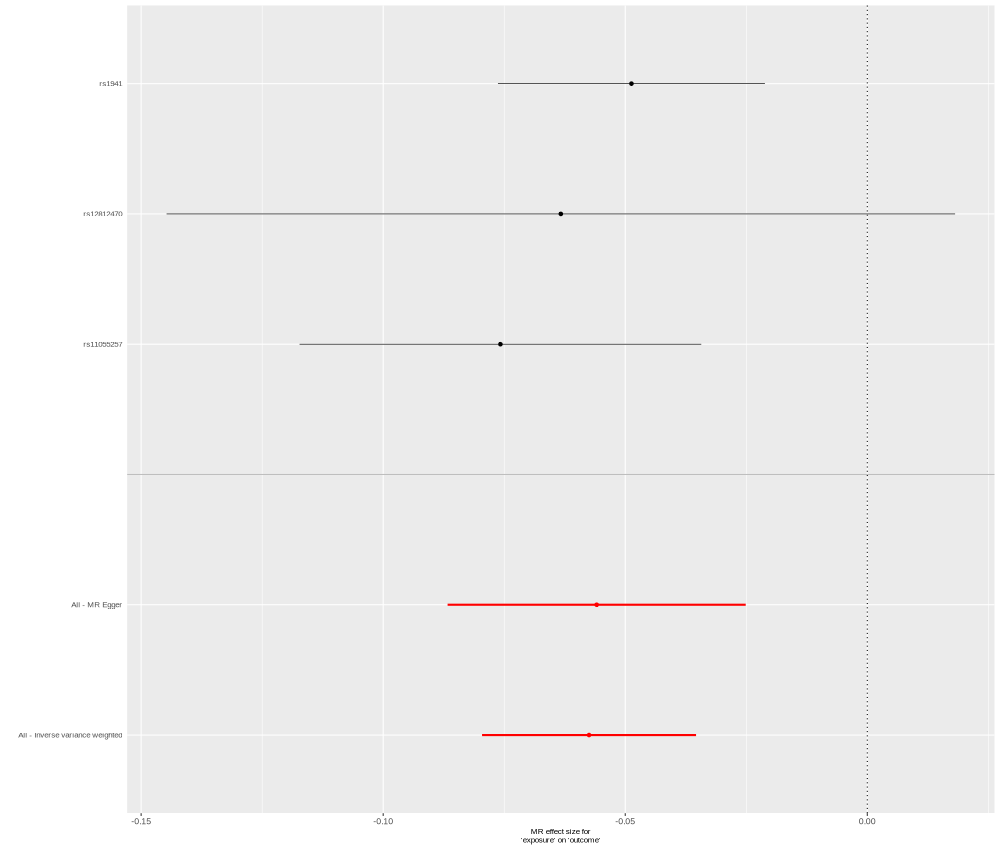

### HYOU1_Q9Y4L1_OID20300_v1_Cardiometabolic_singleSNP.png

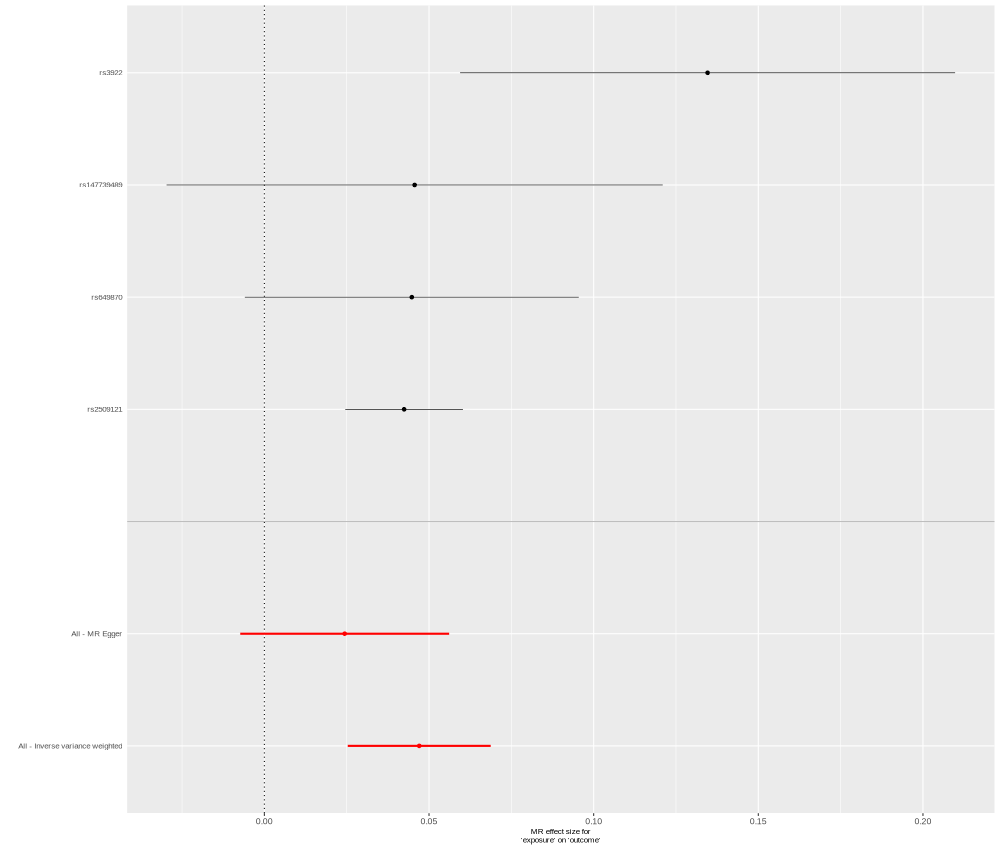

### IGFBP7_Q16270_OID20236_v1_Cardiometabolic.png

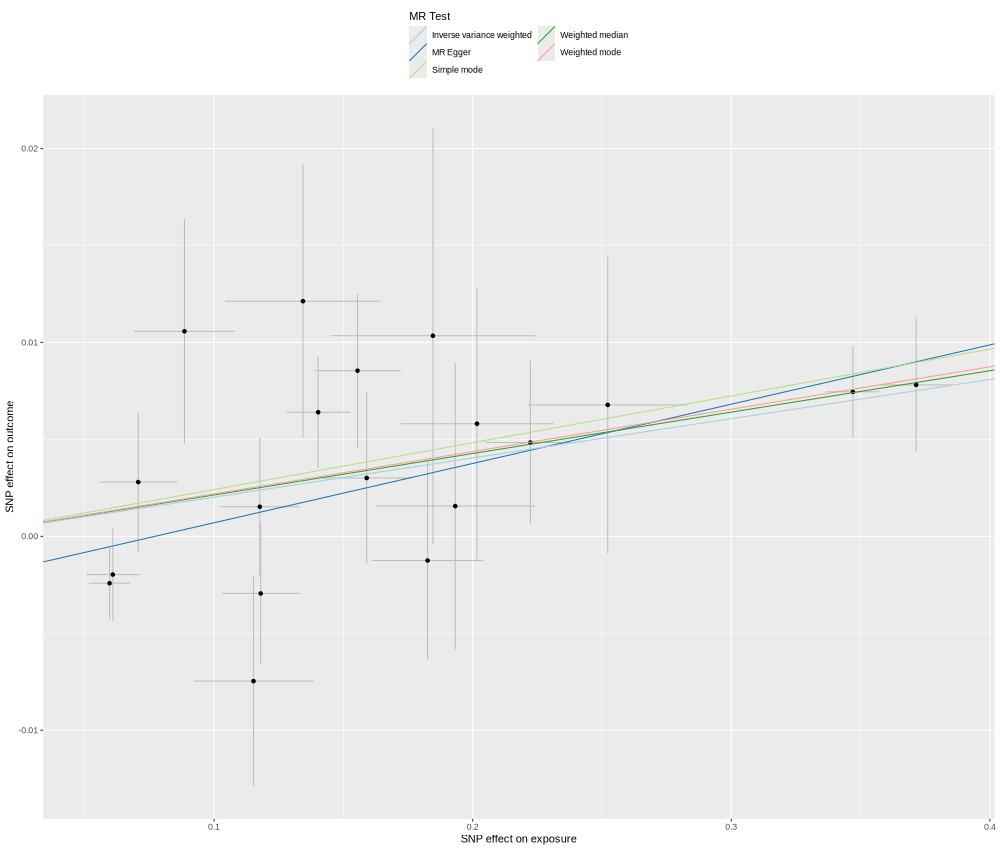

### IL1RN_P18510_OID20700_v1_Inflammation.png

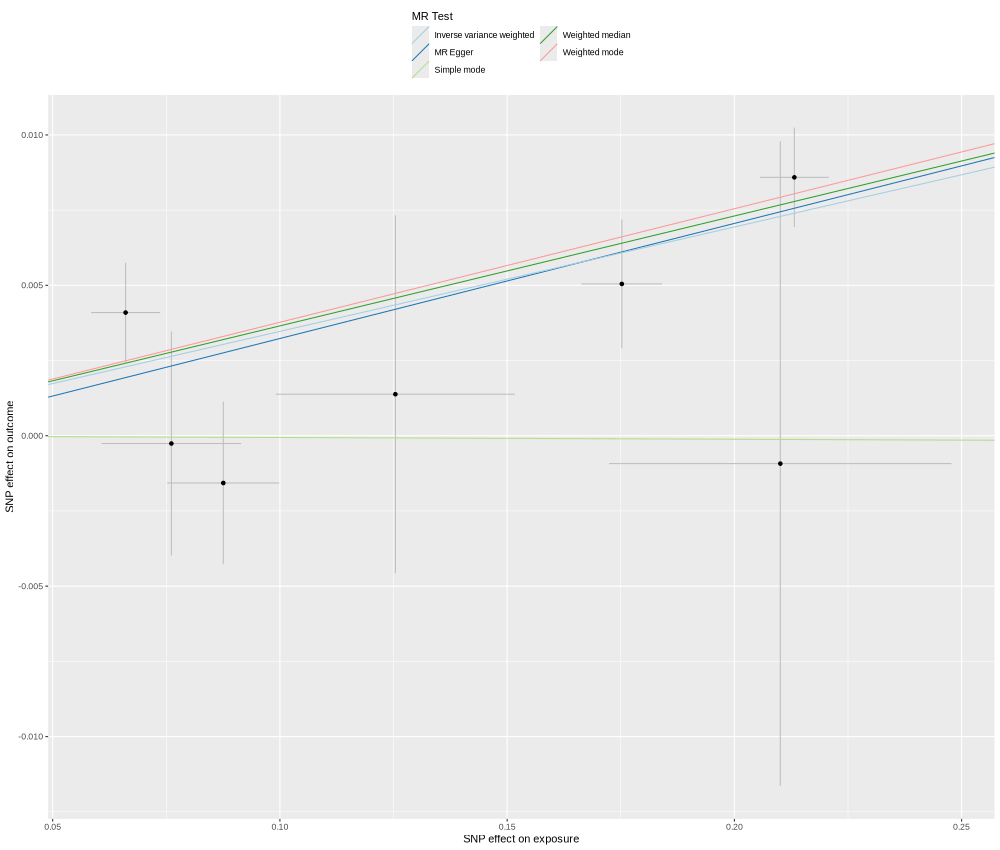

### IL1RN_P18510_OID20700_v1_Inflammation_singleSNP.png

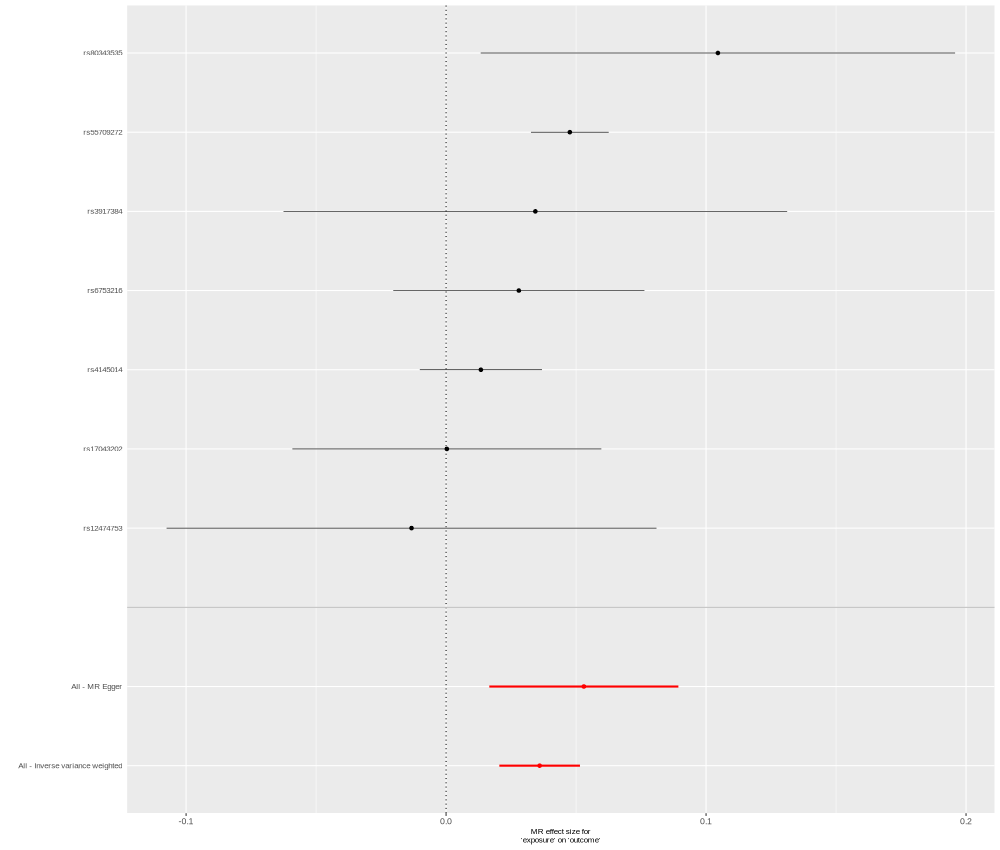

### INHBB_P09529_OID30625_v1_Inflammation_II.png

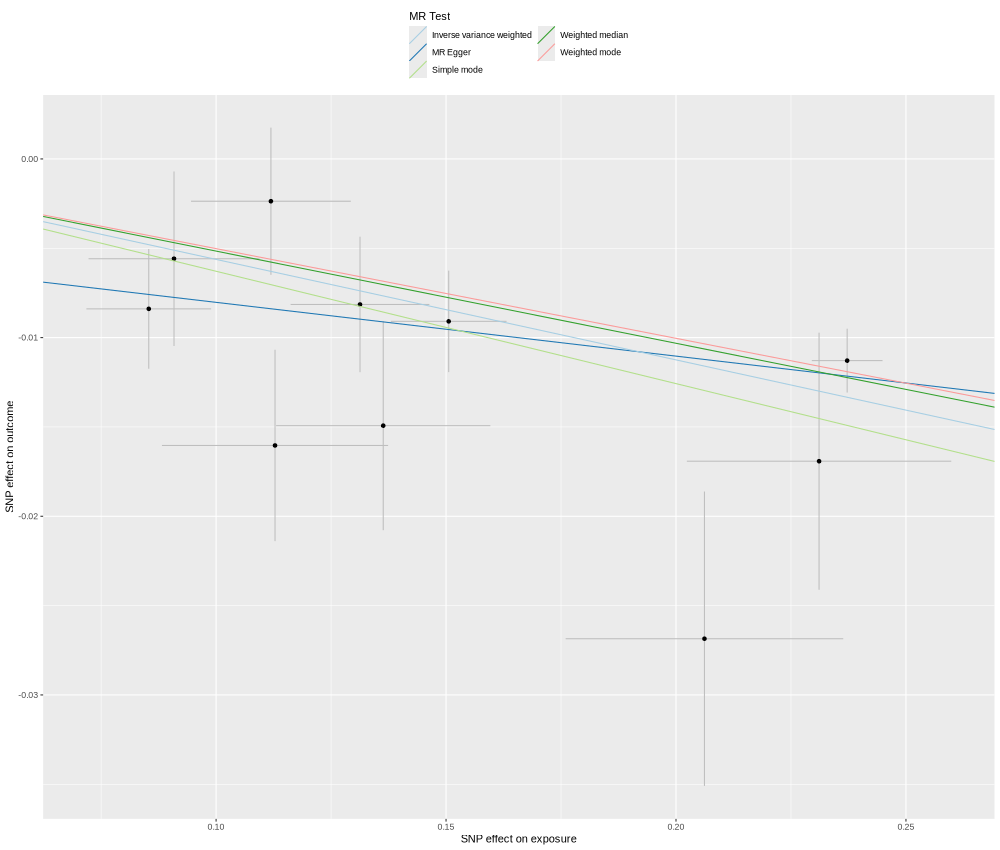

### INHBB_P09529_OID30625_v1_Inflammation_II_singleSNP.png

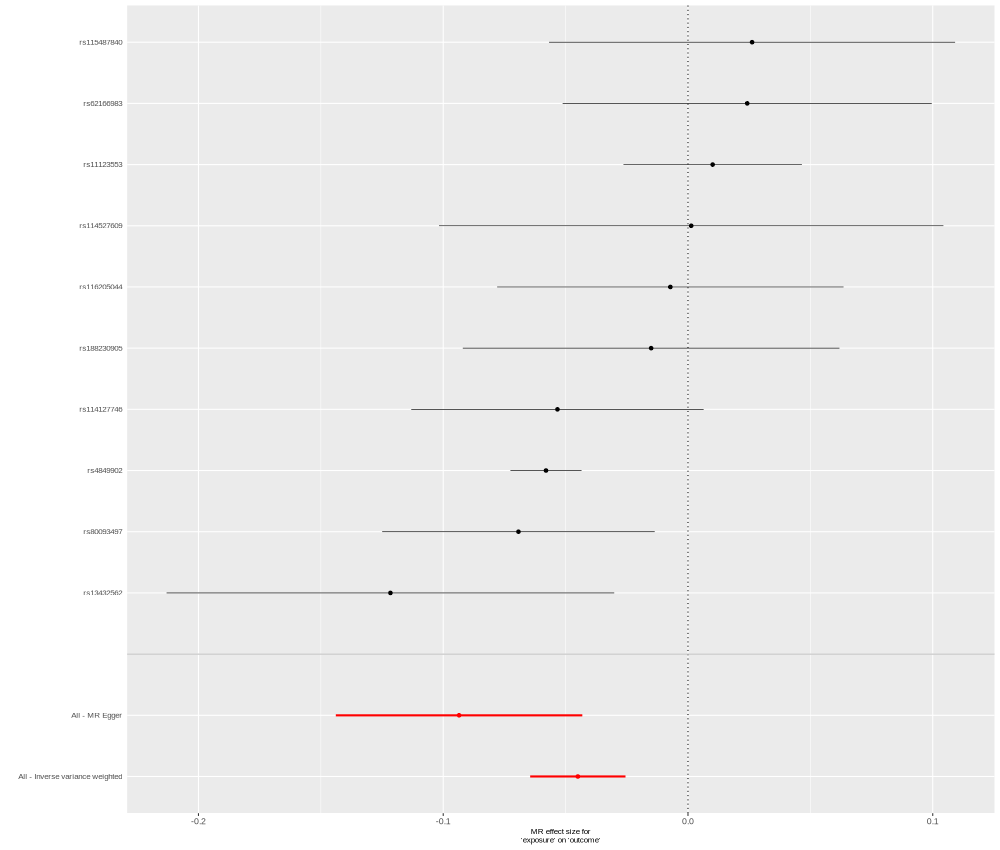

### INHBC_P55103_OID21093_v1_Neurology_singleSNP.png

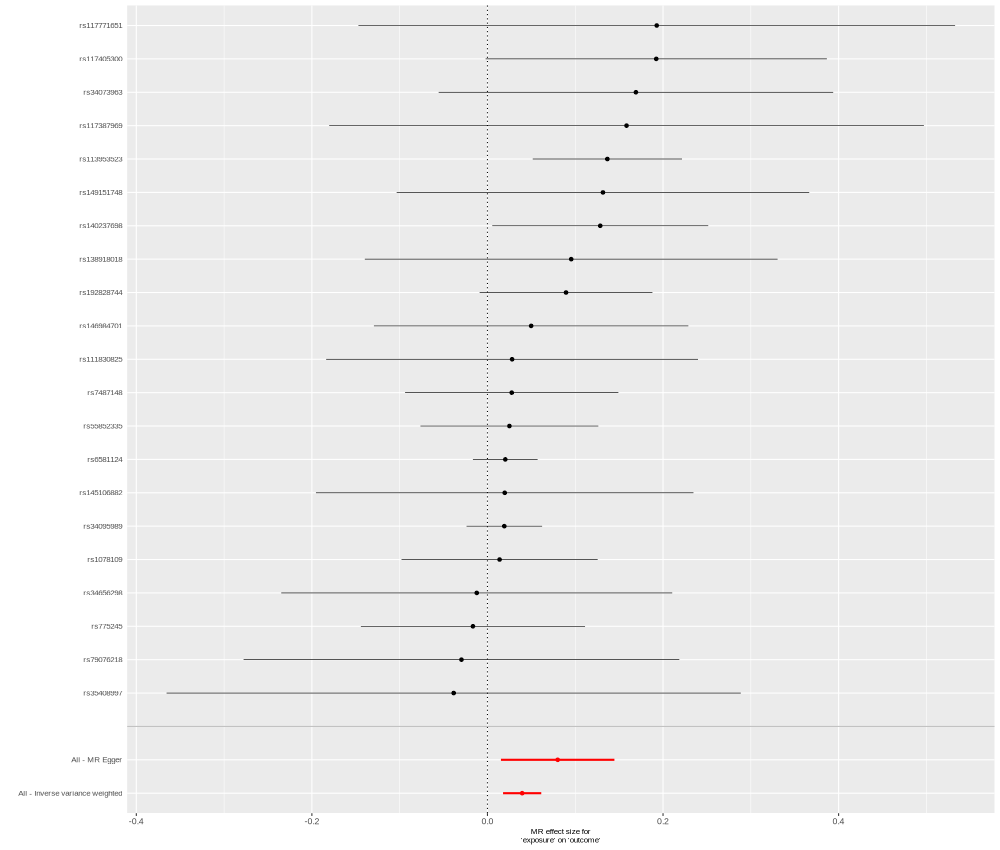

### INHBC_P55103_OID21093_v1_Neurology_singleSNP.png

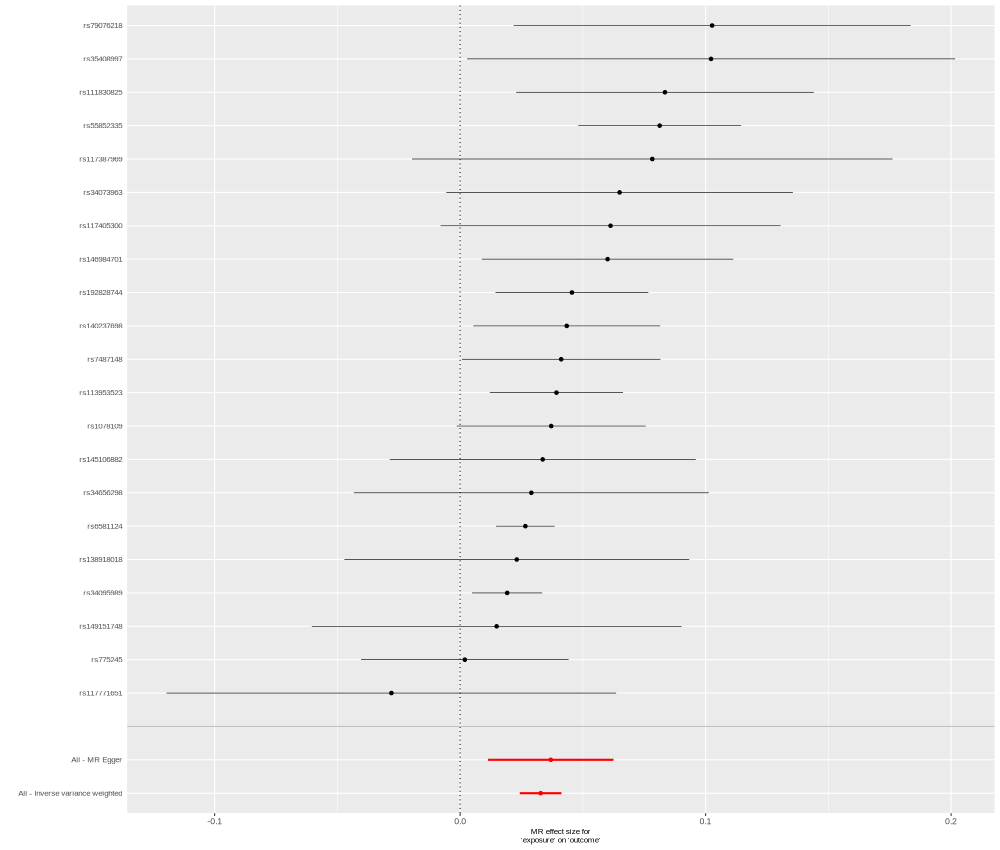

### INSR_P06213_OID30566_v1_Inflammation_II_singleSNP.png

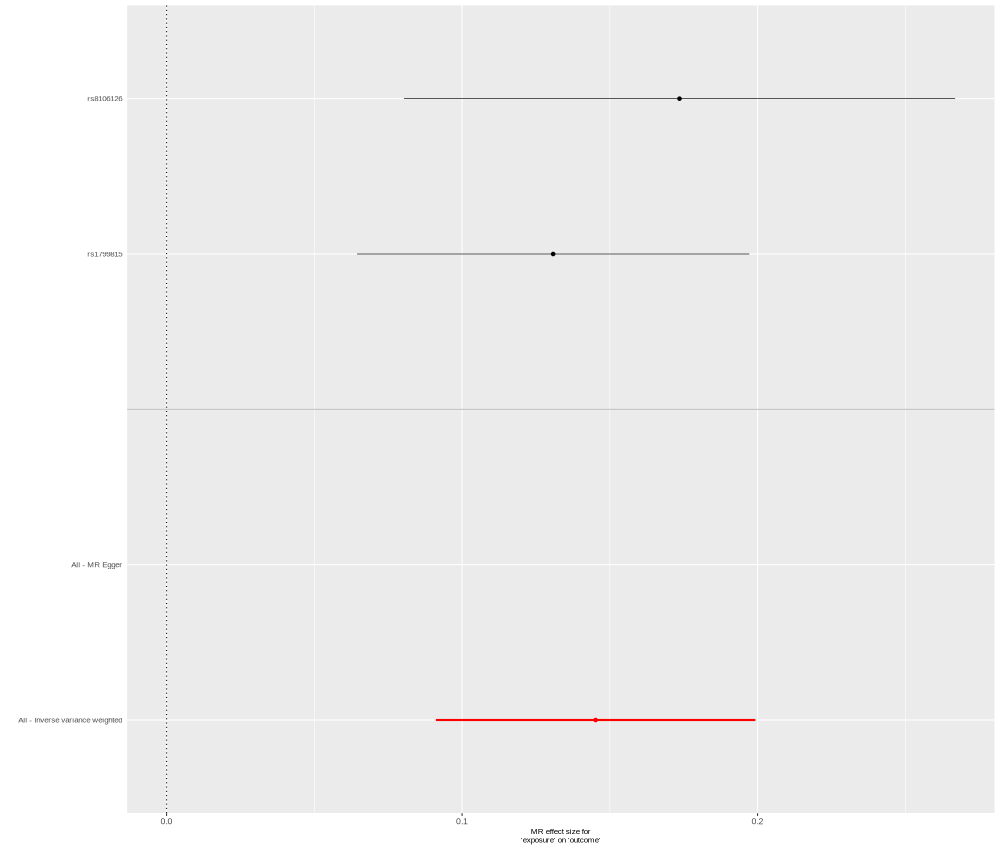

### ITIH3_Q06033_OID20352_v1_Cardiometabolic.png

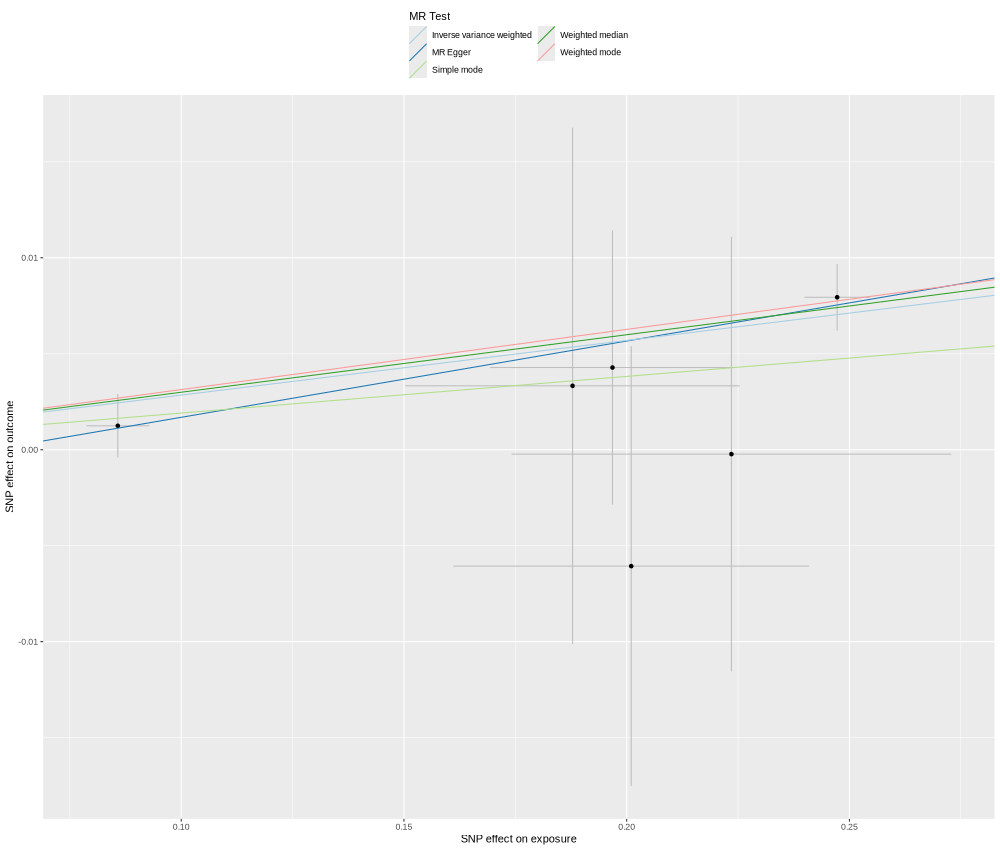

### KYAT1_Q16773_OID20232_v1_Cardiometabolic.png

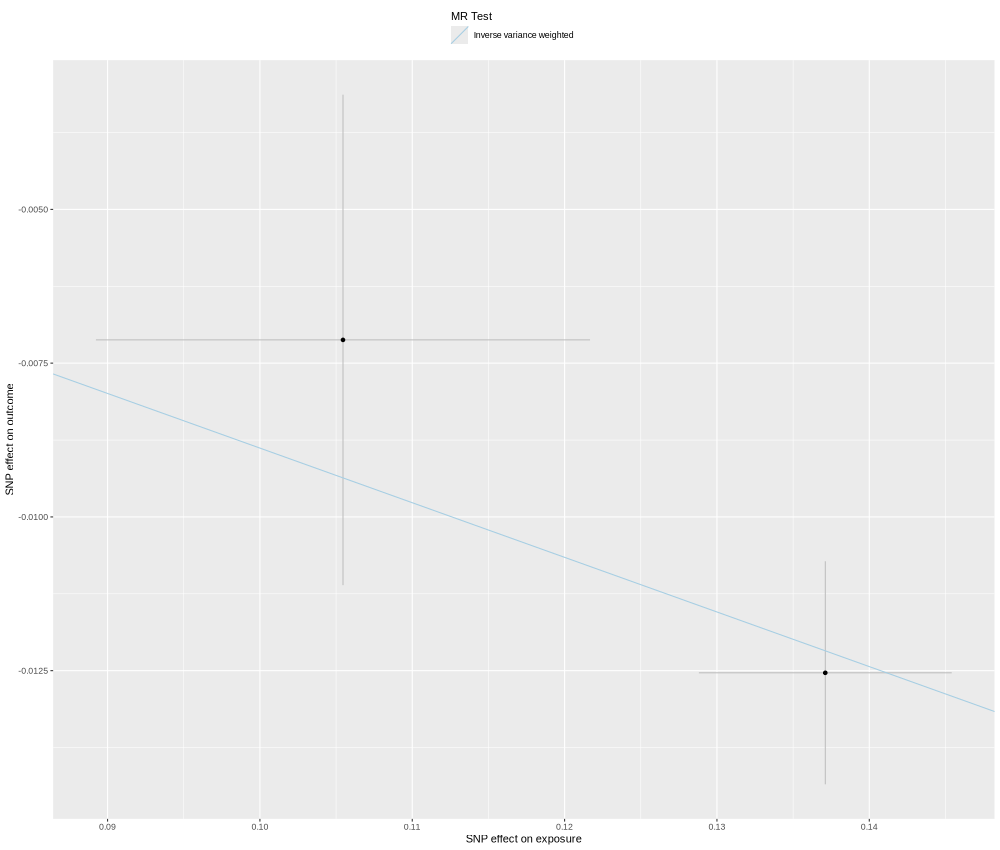

### LDLRAP1_Q5SW96_OID31068_v1_Neurology_II_singleSNP.png

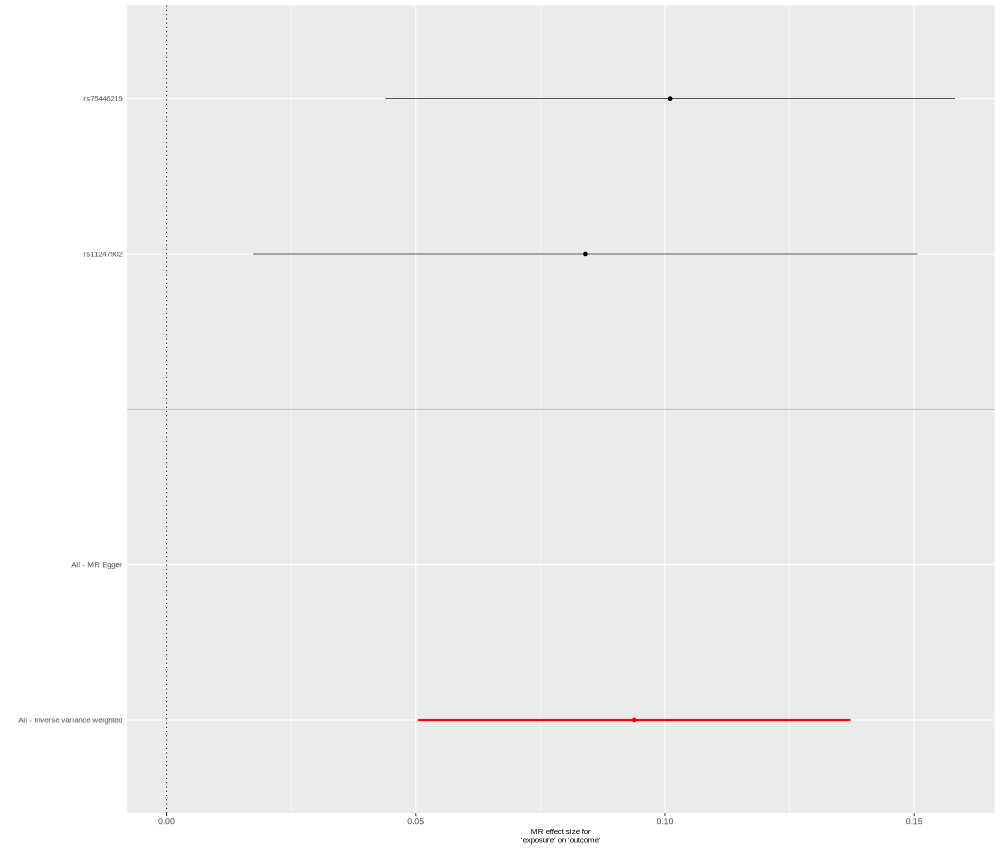

### LPA_P08519_OID30747_v1_Inflammation_II.png

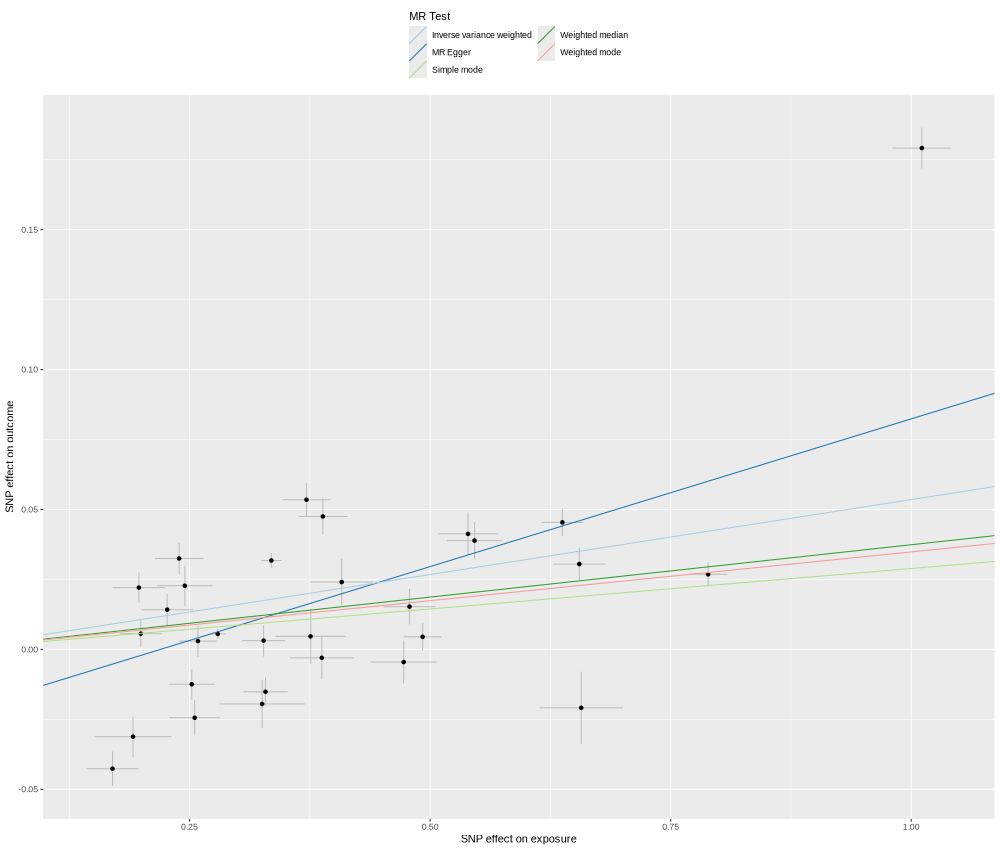

### LPA_P08519_OID30747_v1_Inflammation_II_singleSNP.png

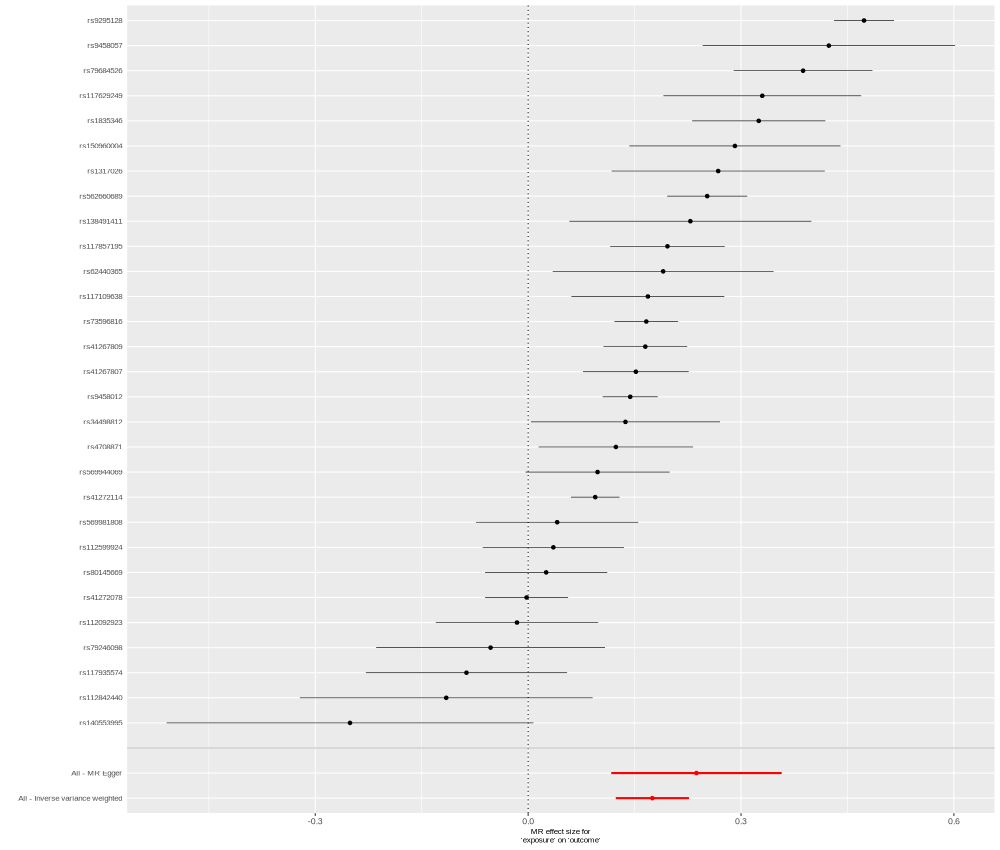

### LRRFIP1_Q32MZ4_OID31344_v1_Oncology_II.png

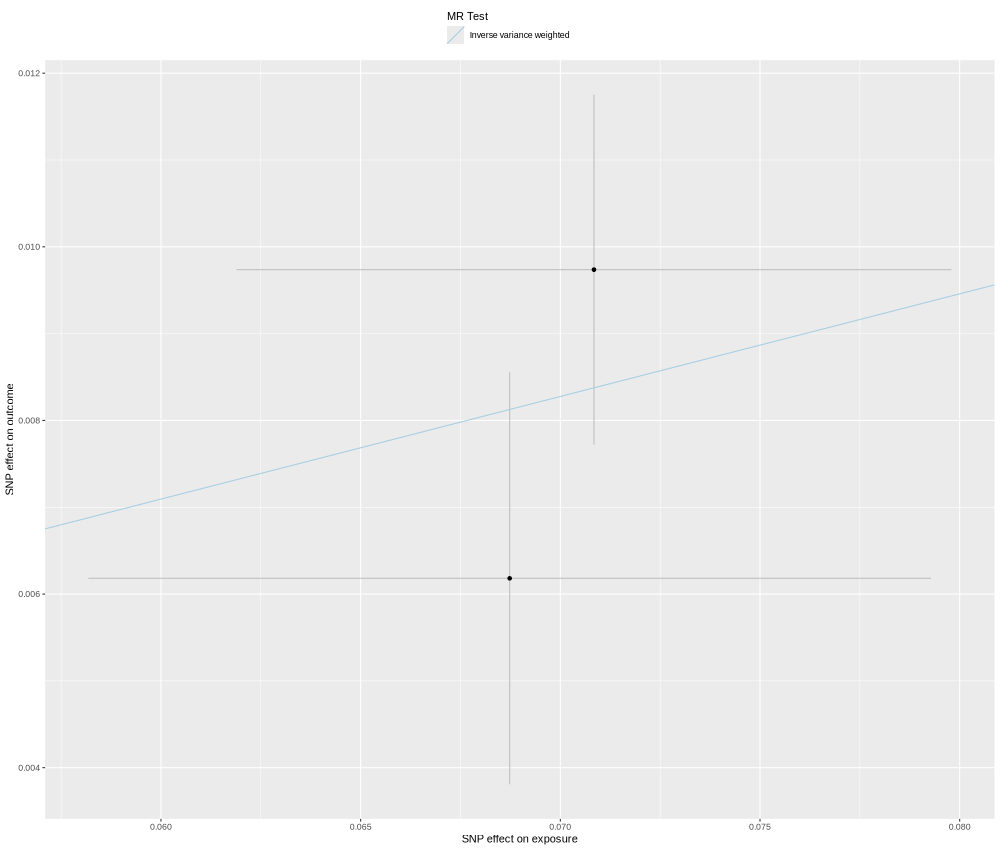

### MEGF9_Q9H1U4_OID20285_v1_Cardiometabolic_singleSNP.png

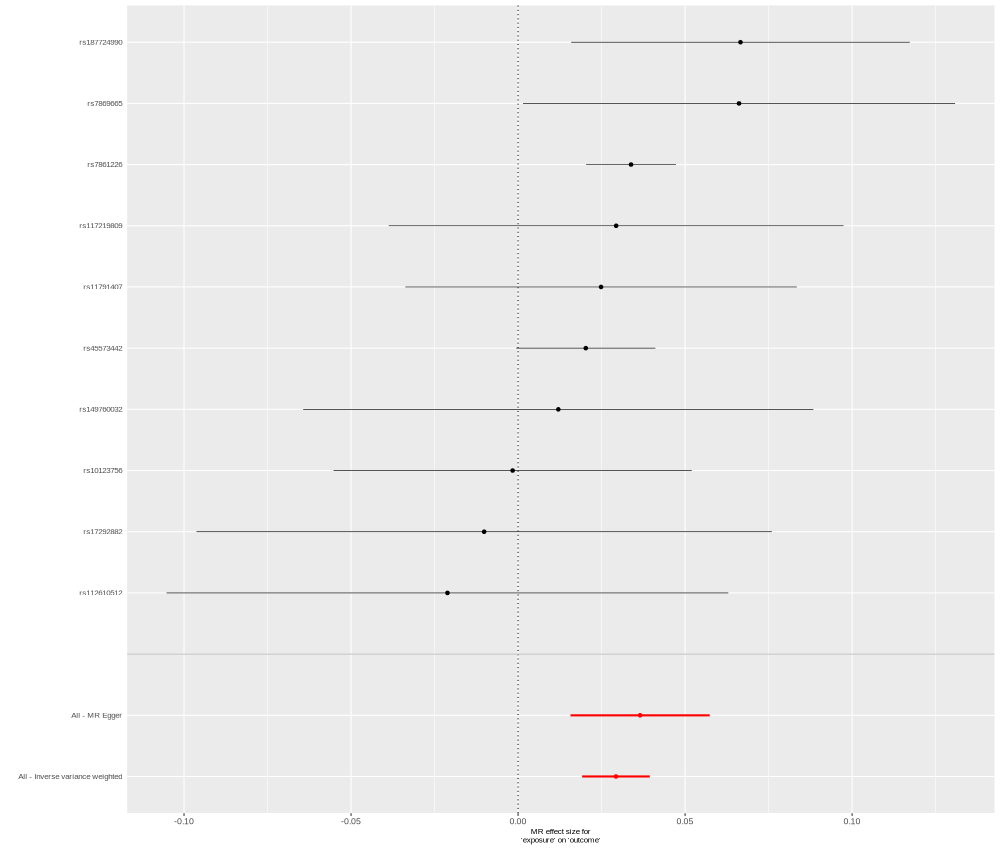

### MERTK_Q12866_OID20657_v1_Inflammation_singleSNP.png

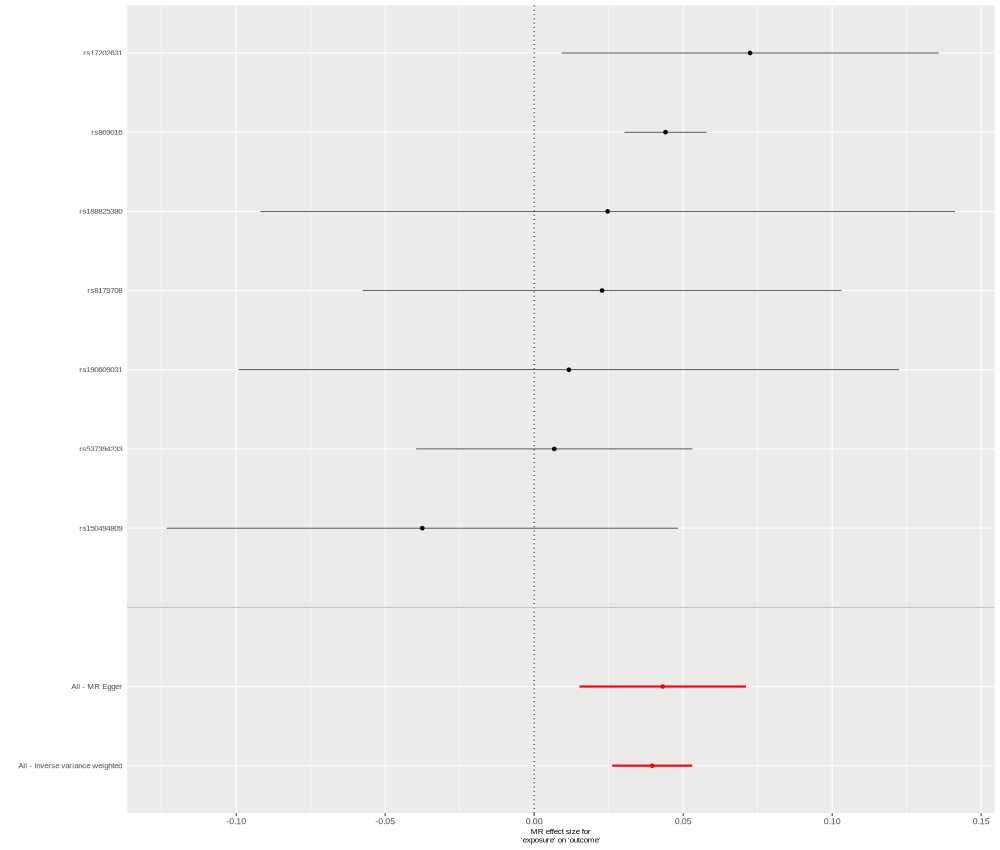

### MIA_Q16674_OID21531_v1_Oncology.png

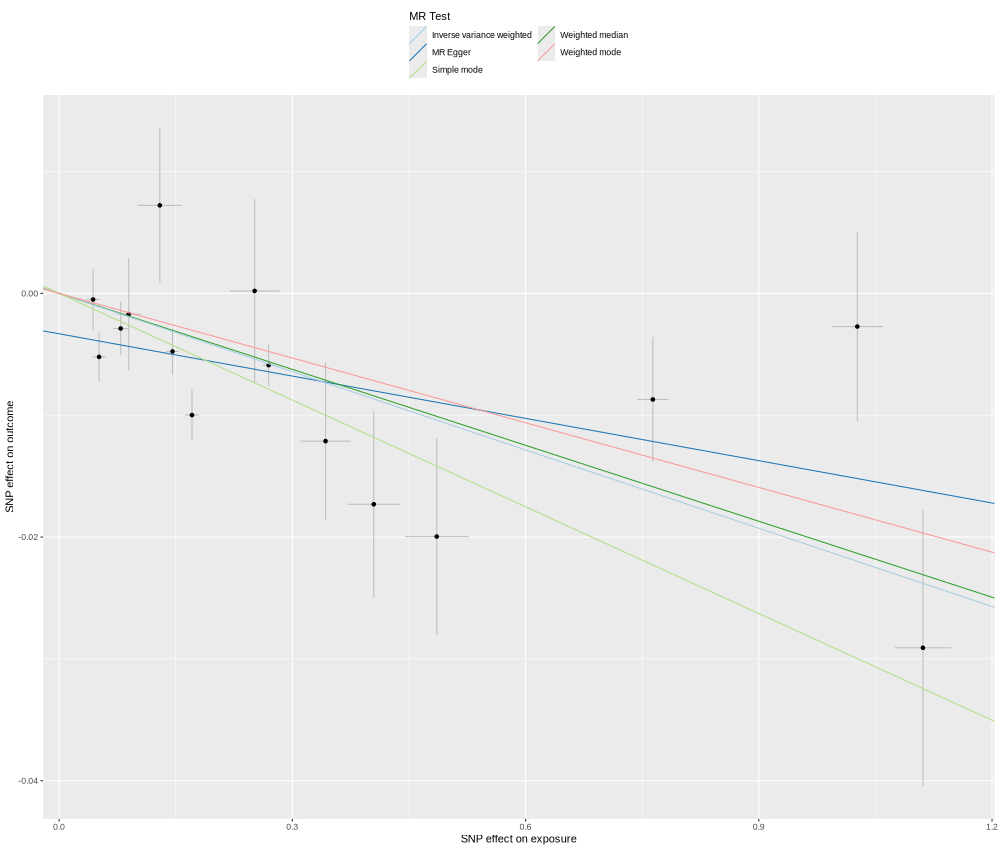

### MIA_Q16674_OID21531_v1_Oncology_singleSNP.png

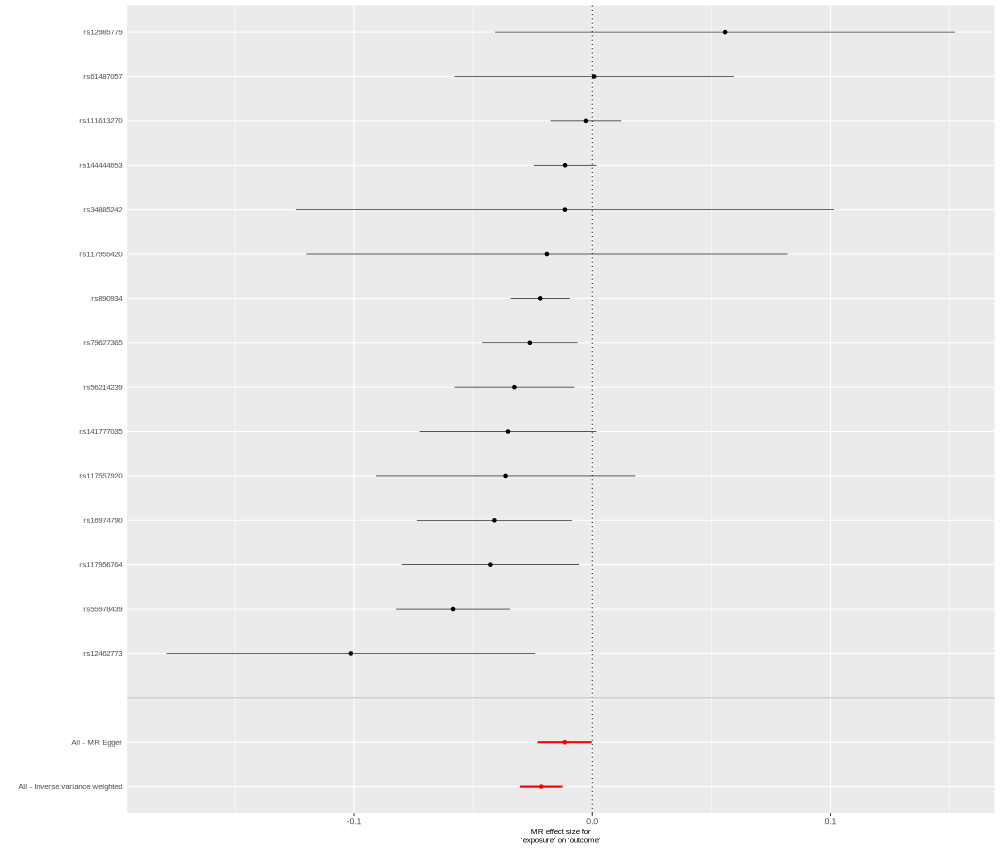

### MLN_P12872_OID20541_v1_Inflammation_singleSNP.png

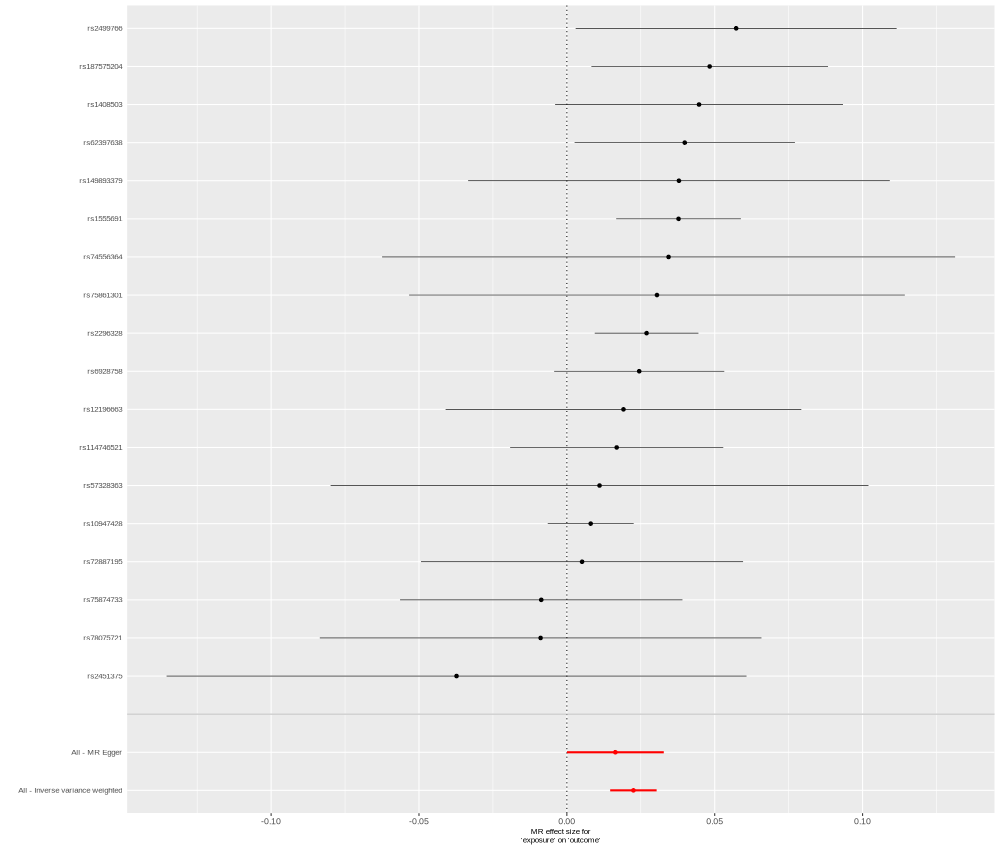

### MPI_P34949_OID21288_v1_Oncology.png

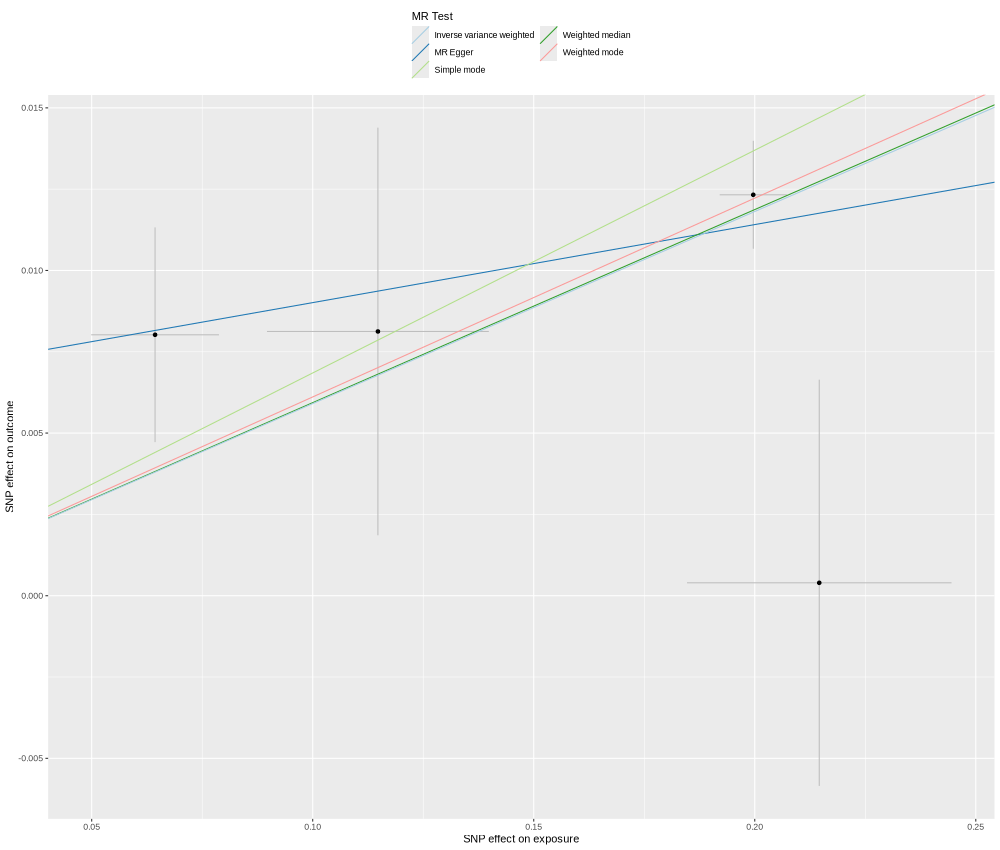

### MSR1_P21757_OID21063_v1_Neurology_singleSNP.png

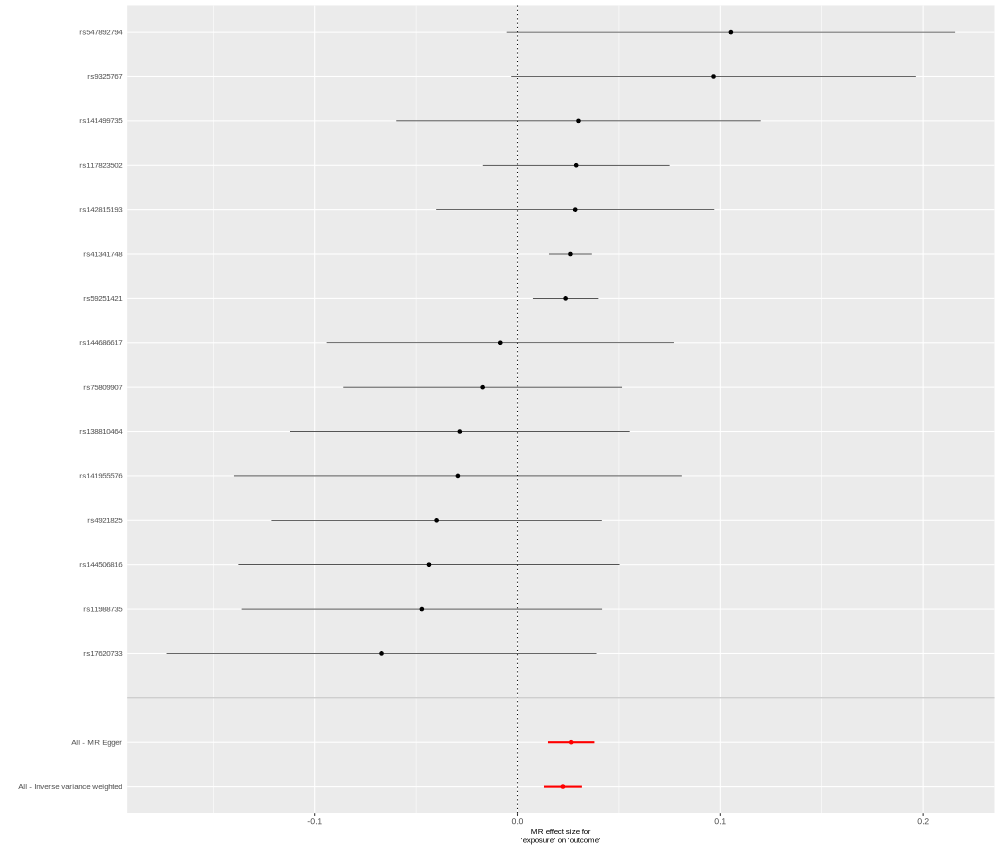

### OPLAH_O14841_OID30201_v1_Cardiometabolic_II.png

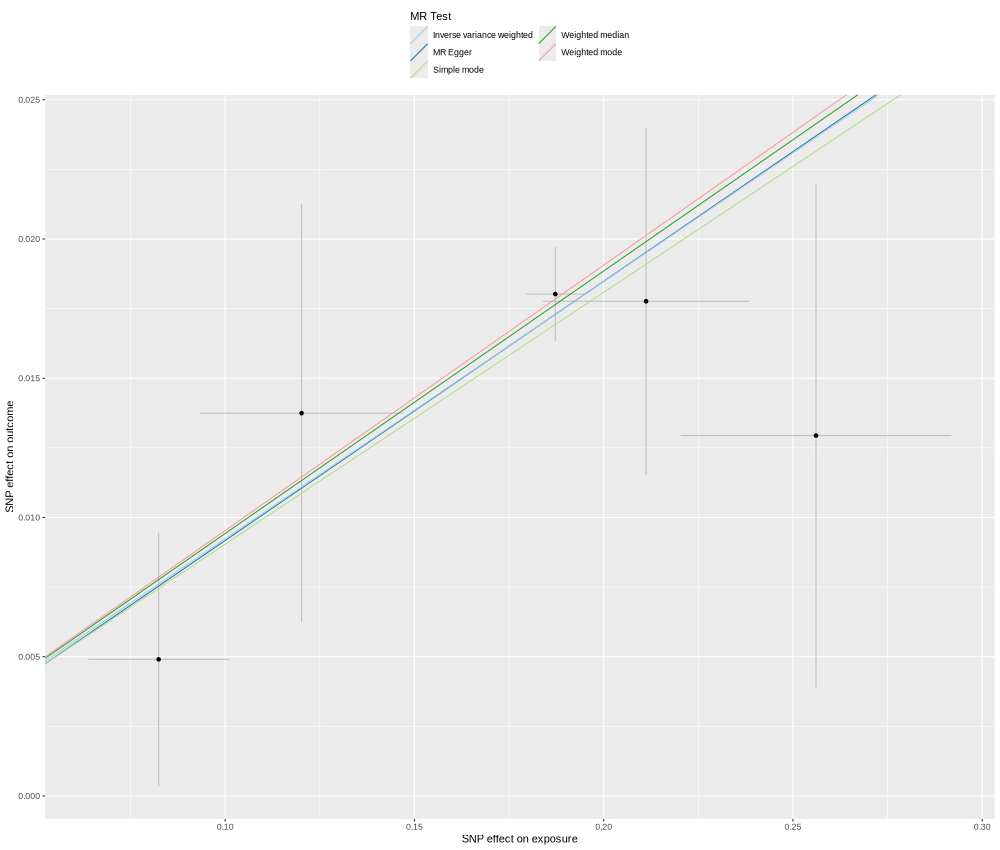

### PCSK9_Q8NBP7_OID20235_v1_Cardiometabolic.png

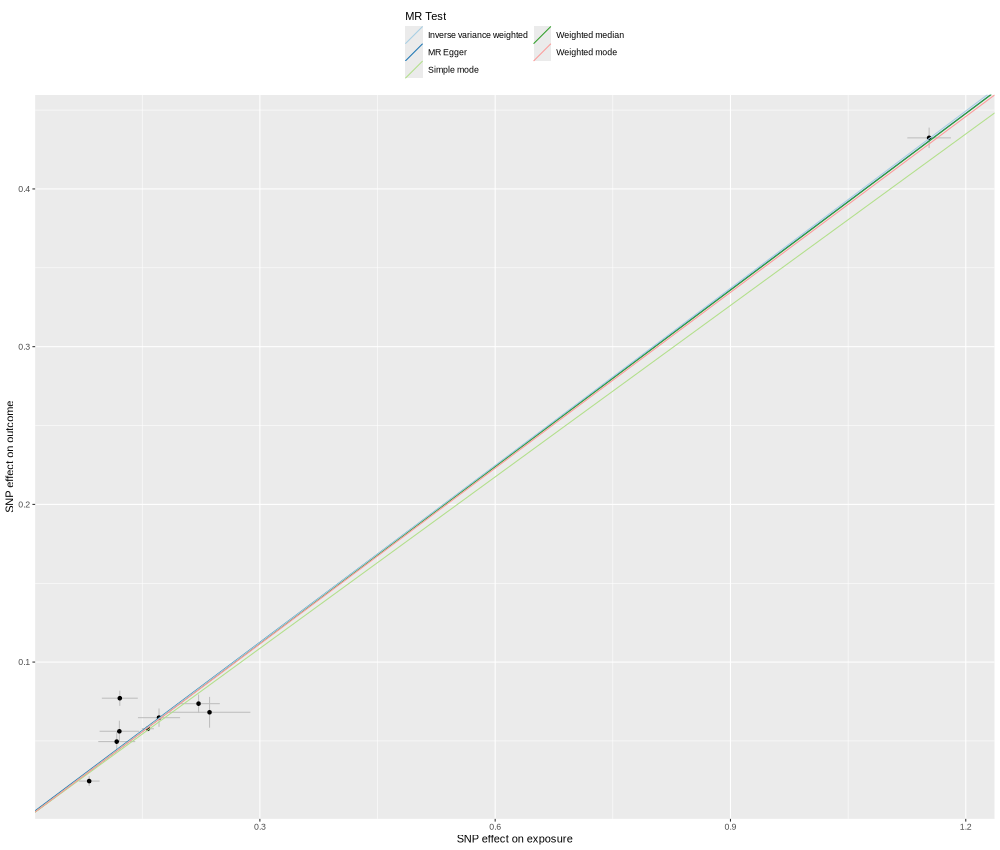

### PCSK9_Q8NBP7_OID20235_v1_Cardiometabolic_singleSNP.png

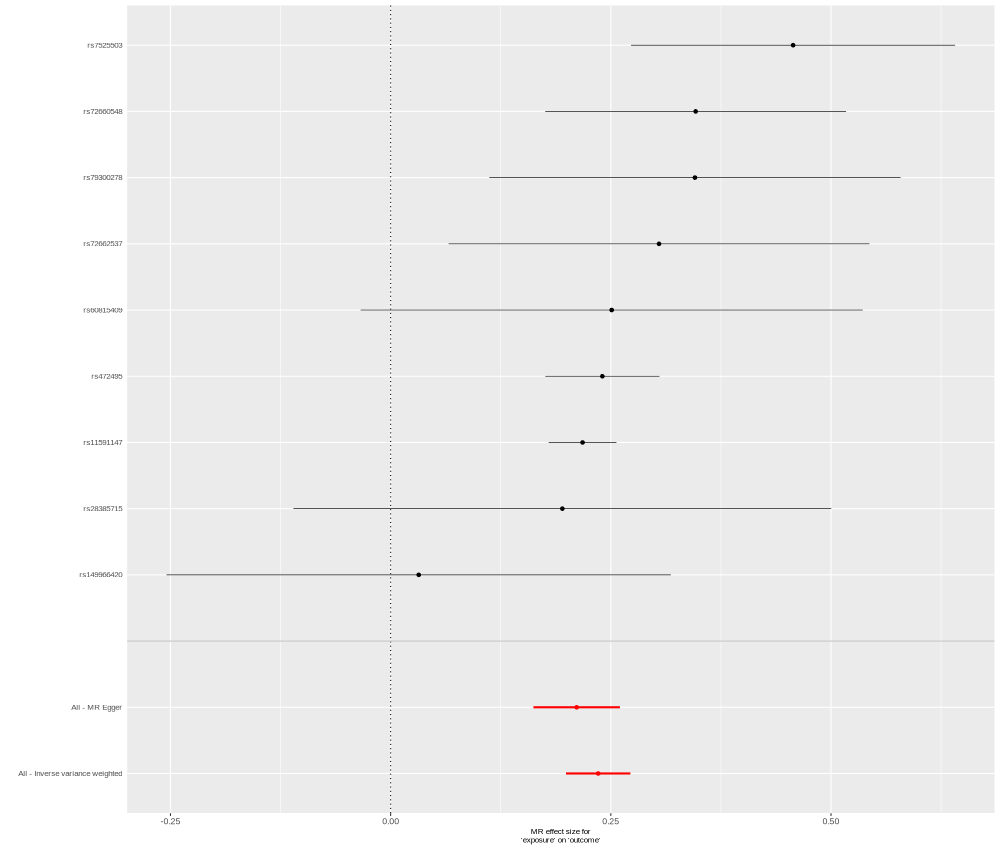

### PF4_P02776_OID30764_v1_Inflammation_II.png

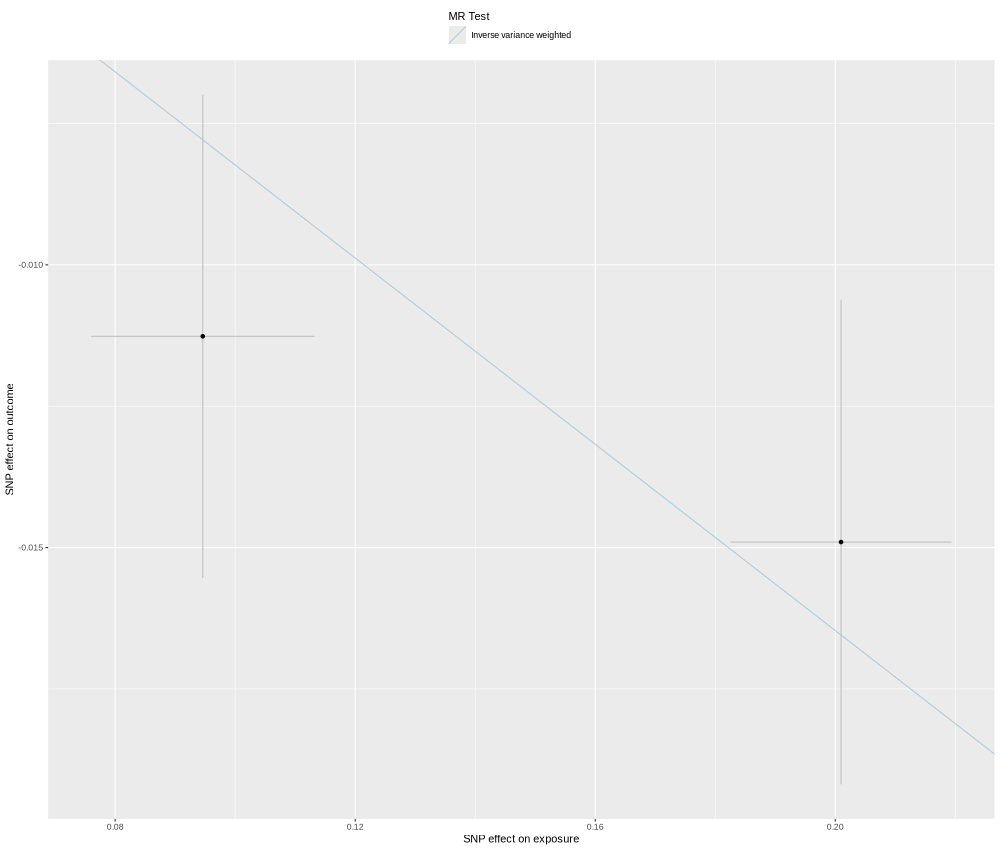

### PKN3_Q6P5Z2_OID31311_v1_Oncology_II.png

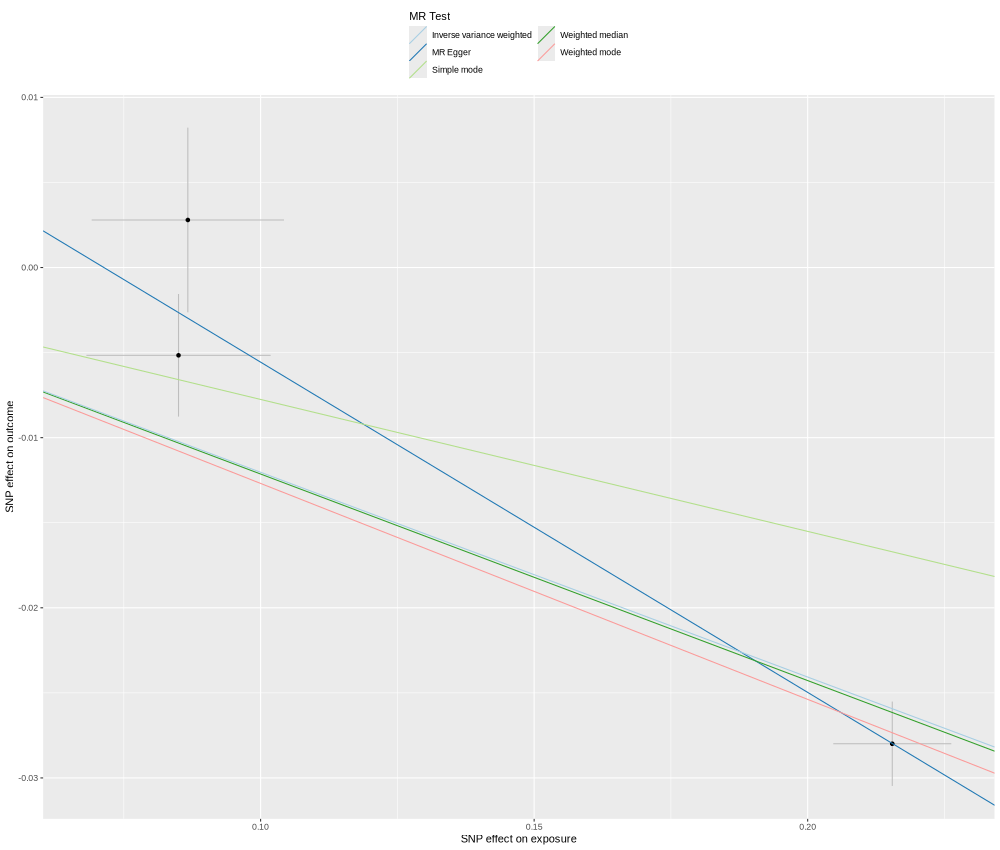

### PLTP_P55058_OID20238_v1_Cardiometabolic_singleSNP.png

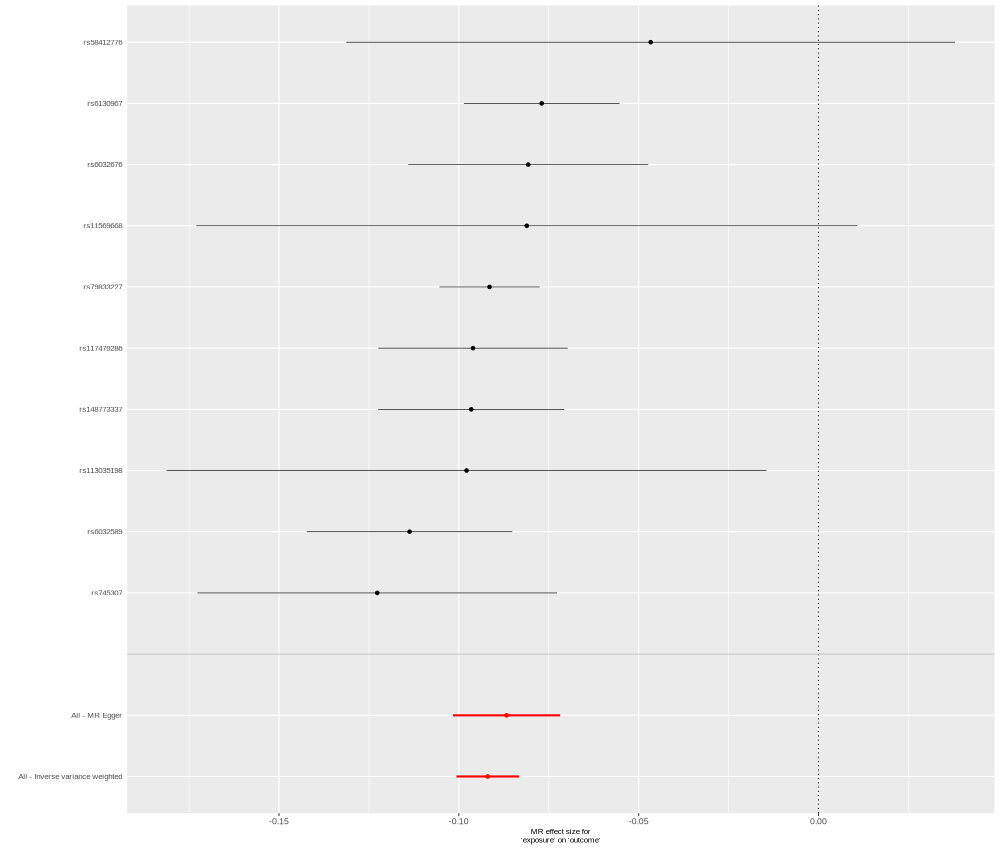

### PLXNB2_O15031_OID20317_v1_Cardiometabolic_singleSNP.png

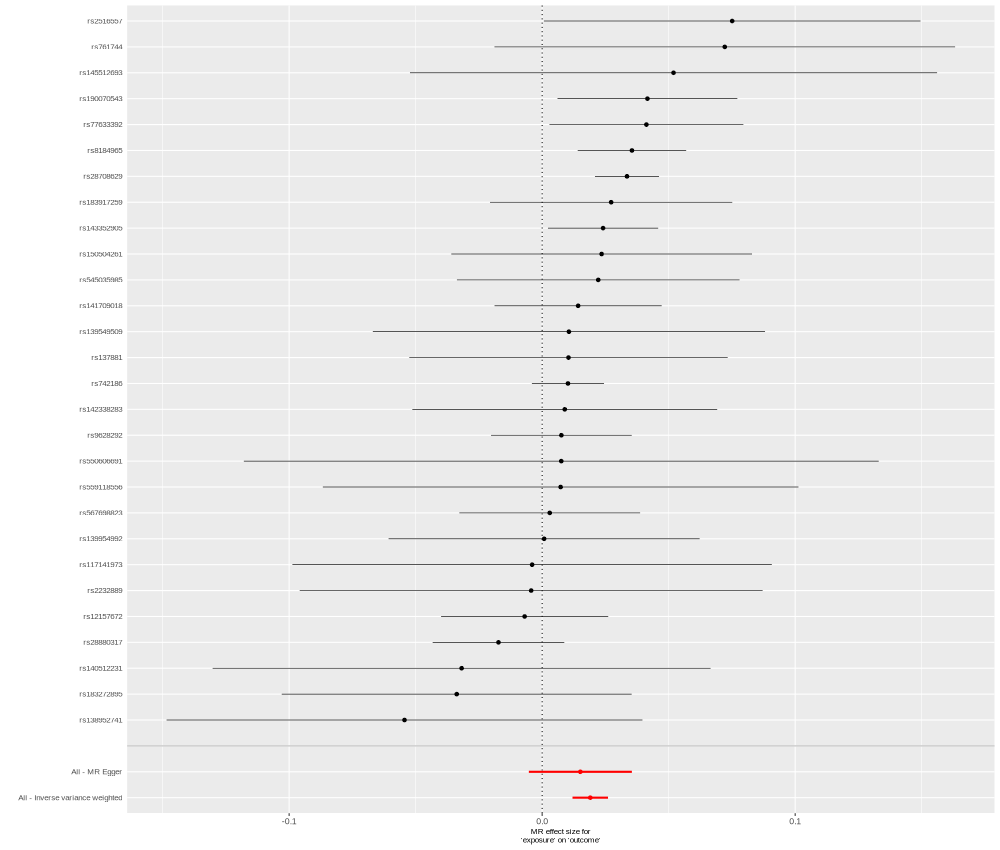

### PNLIPRP2_P54317_OID20775_v1_Inflammation.png

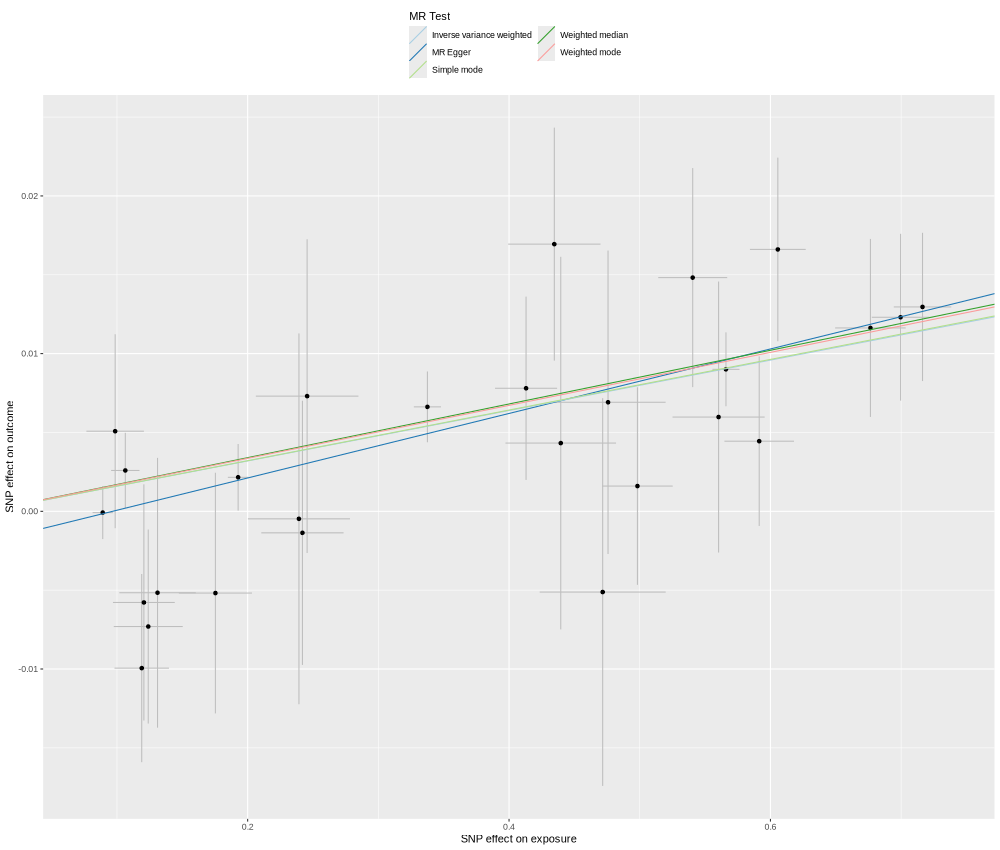

### PPCDC_Q96CD2_OID21027_v1_Neurology.png

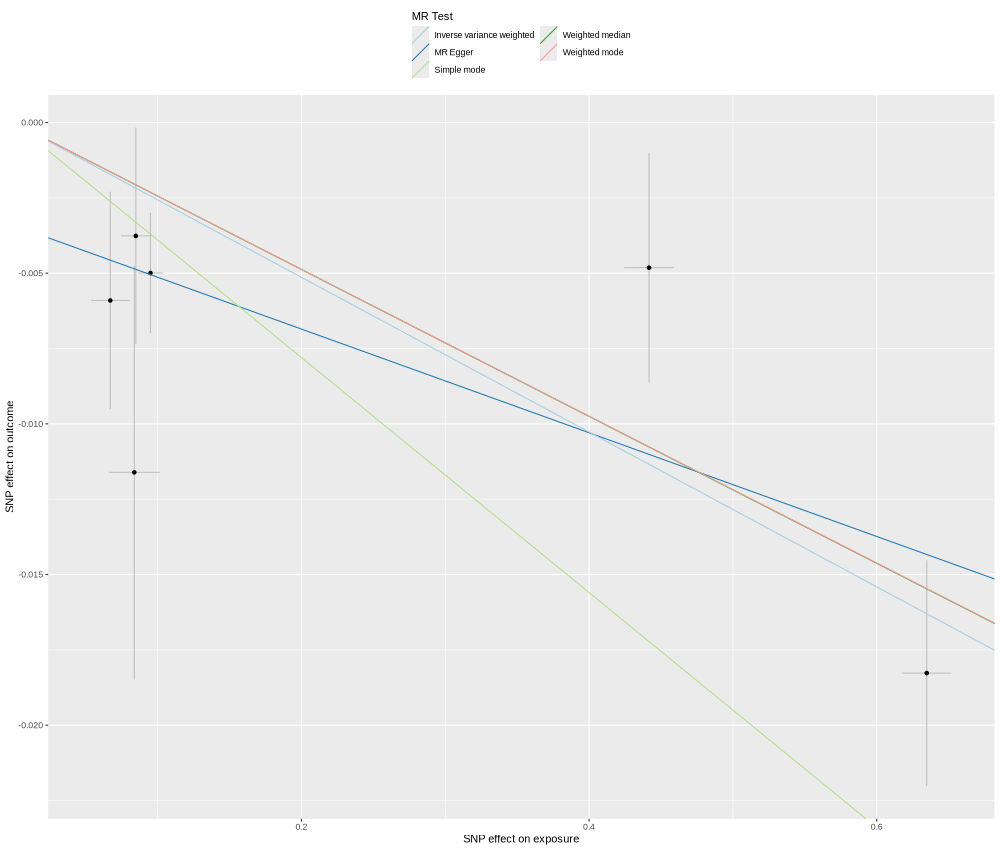

### RSPO3_Q9BXY4_OID21415_v1_Oncology.png

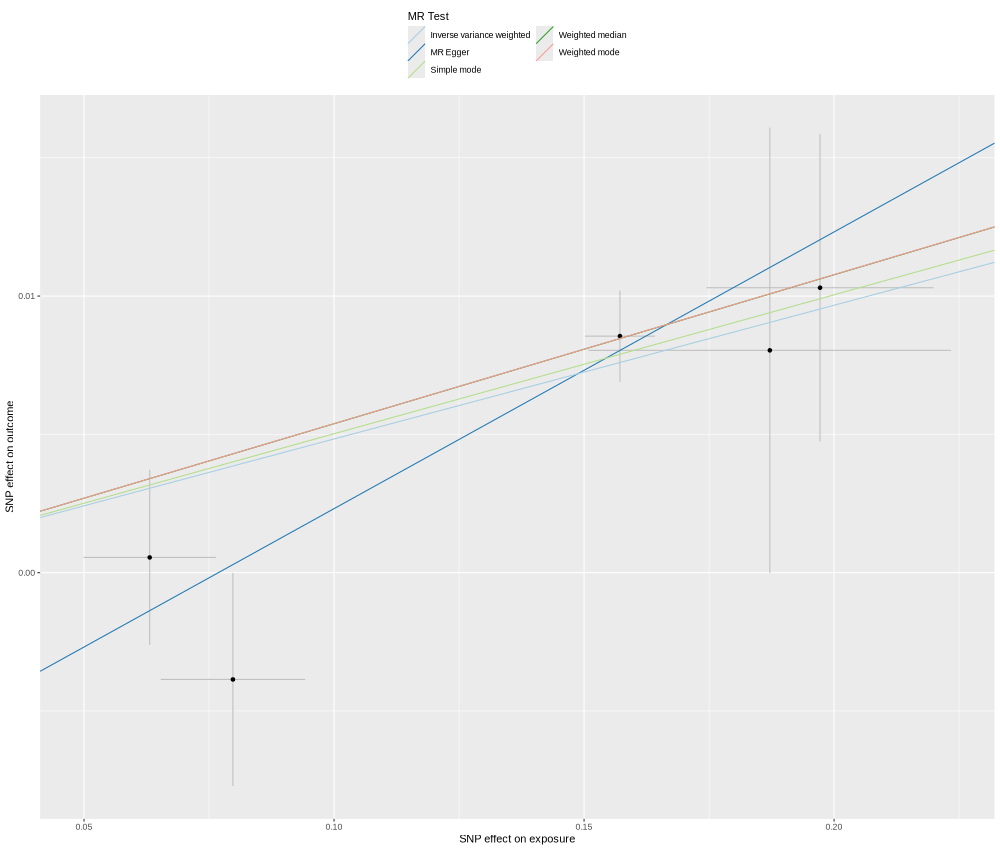

### RSPO3_Q9BXY4_OID21415_v1_Oncology_singleSNP.png

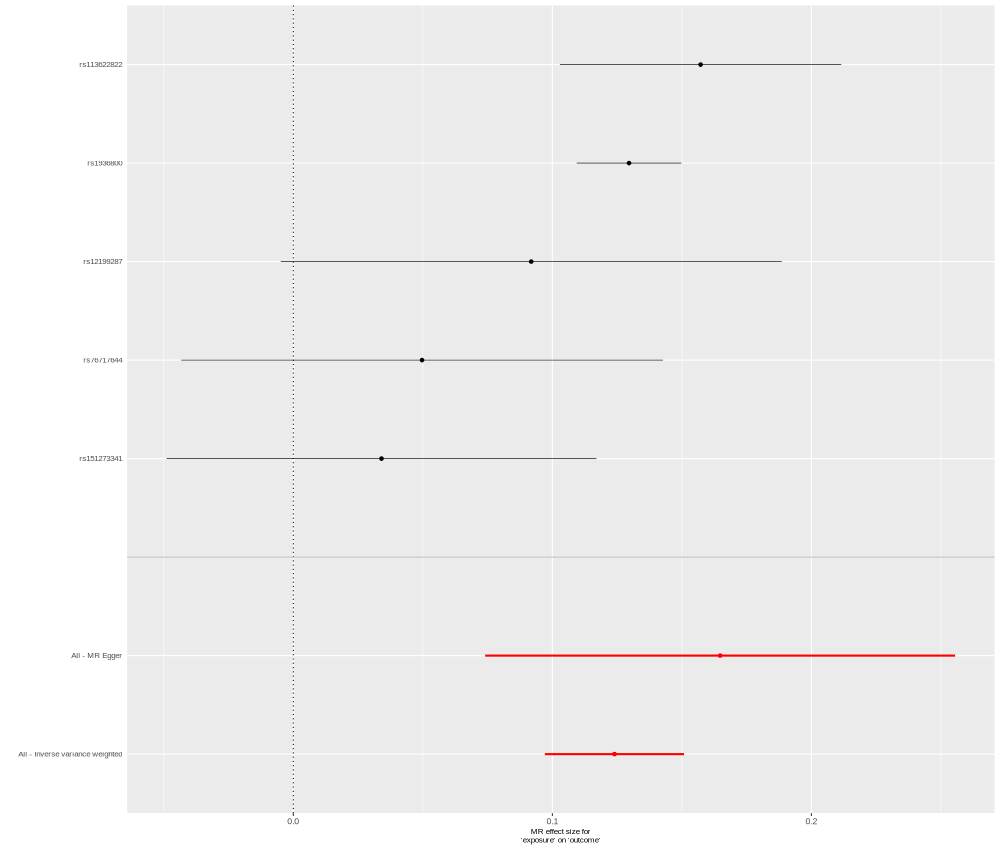

### SERPINA1_P01009_OID30750_v1_Inflammation_II.png

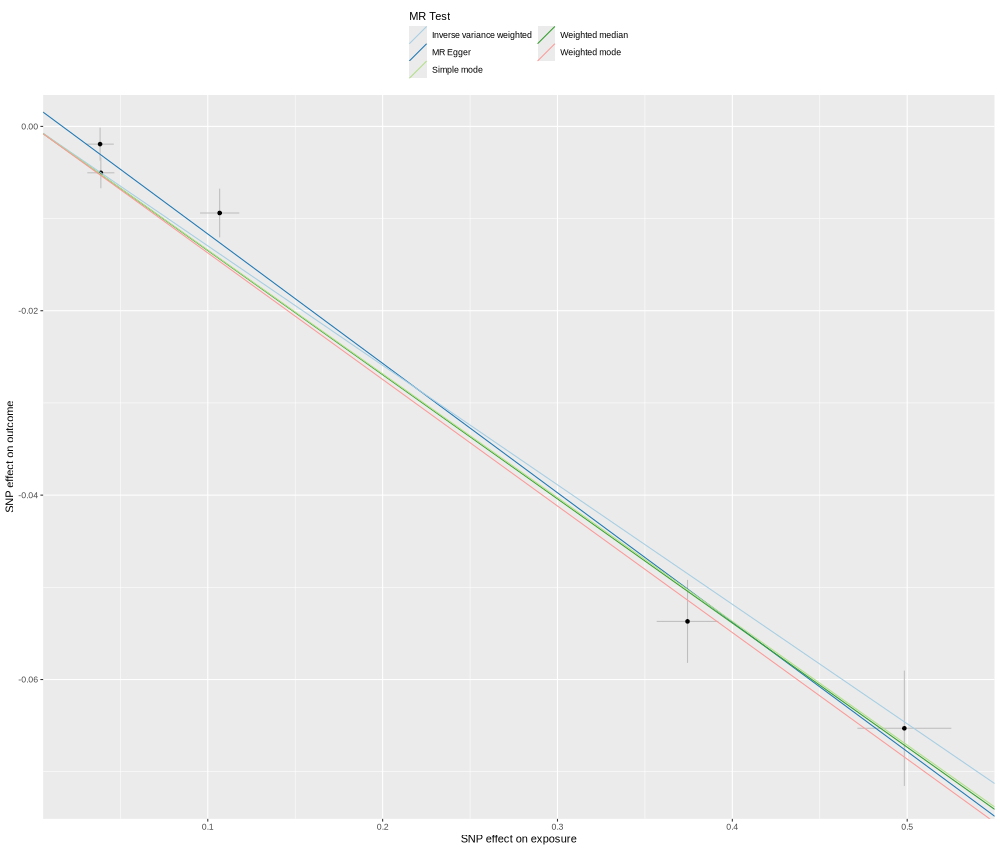

### SERPINF2_P08697_OID30779_v1_Inflammation_II_singleSNP.png

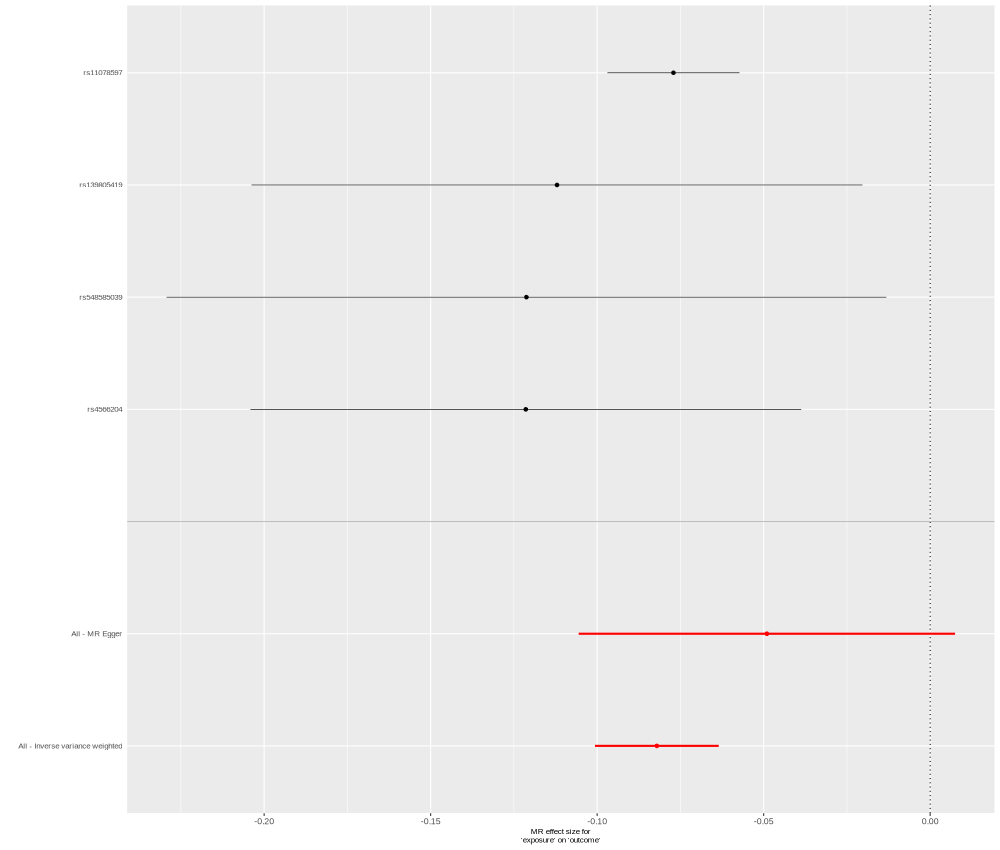

### SHBG_P04278_OID30685_v1_Inflammation_II_singleSNP.png

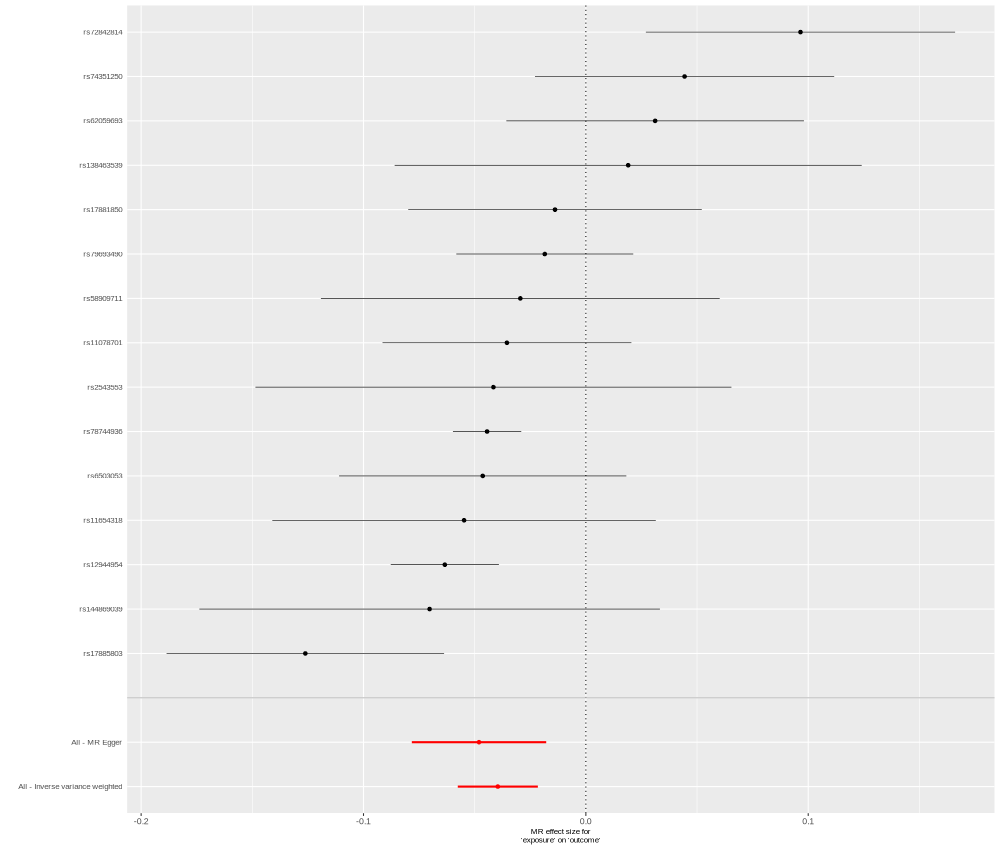

### SHMT1_P34896_OID20714_v1_Inflammation.png

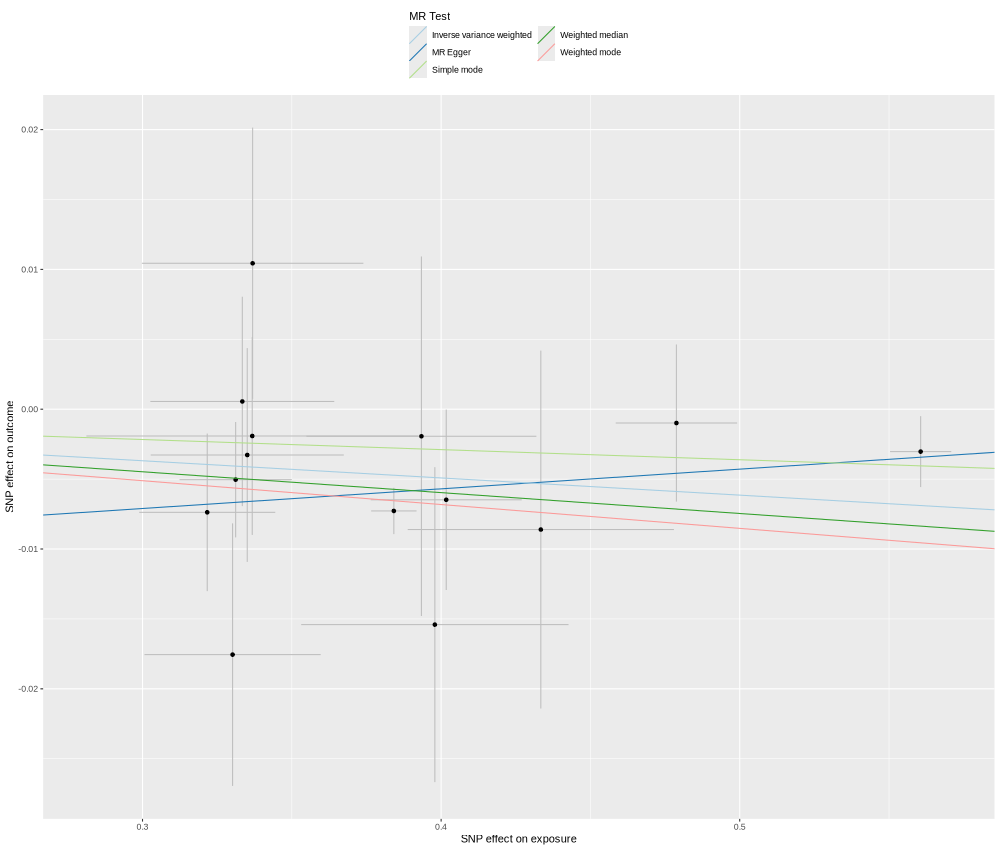

### SORCS2_Q96PQ0_OID21262_v1_Oncology.png

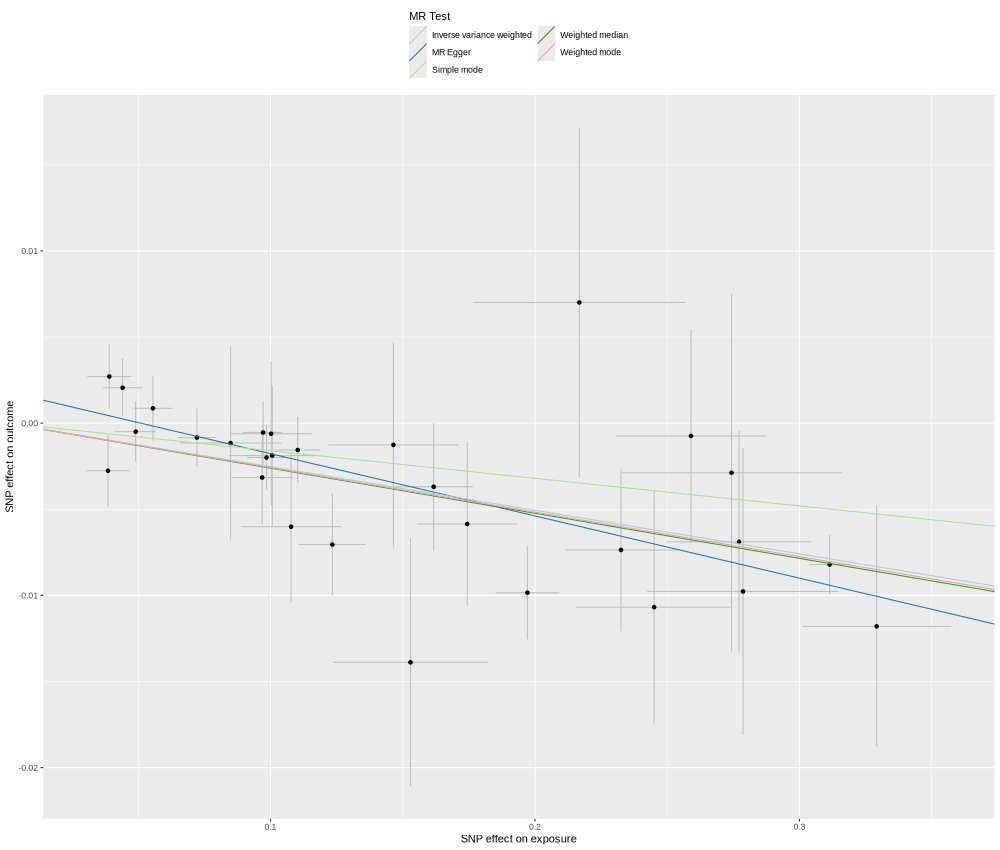

### SULT2A1_Q06520_OID20508_v1_Inflammation_singleSNP.png

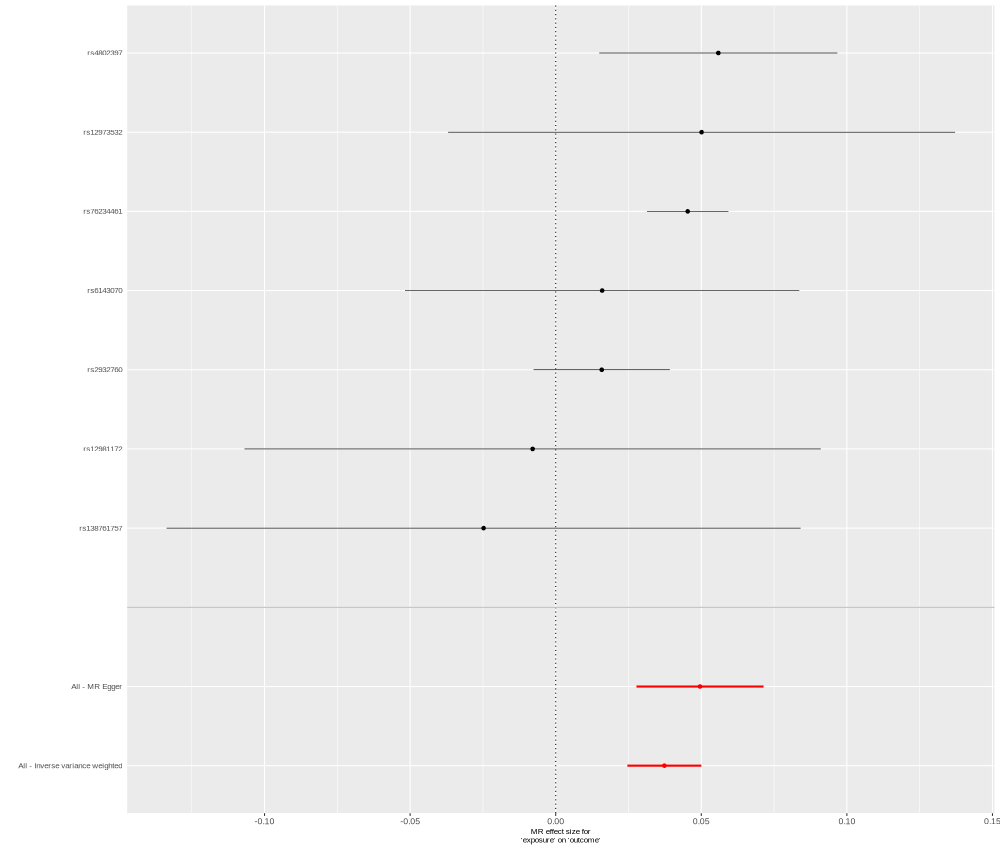

### THOP1_P52888_OID20194_v1_Cardiometabolic.png

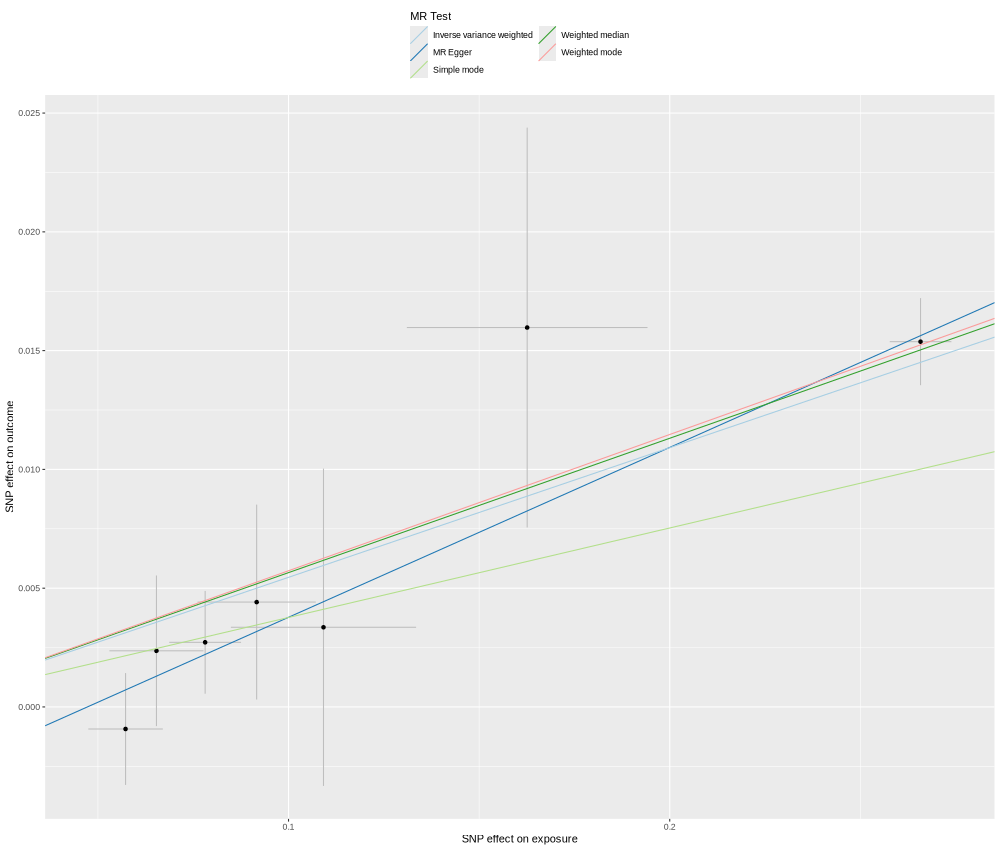

### TIMD4_Q96H15_OID20375_v1_Cardiometabolic.png

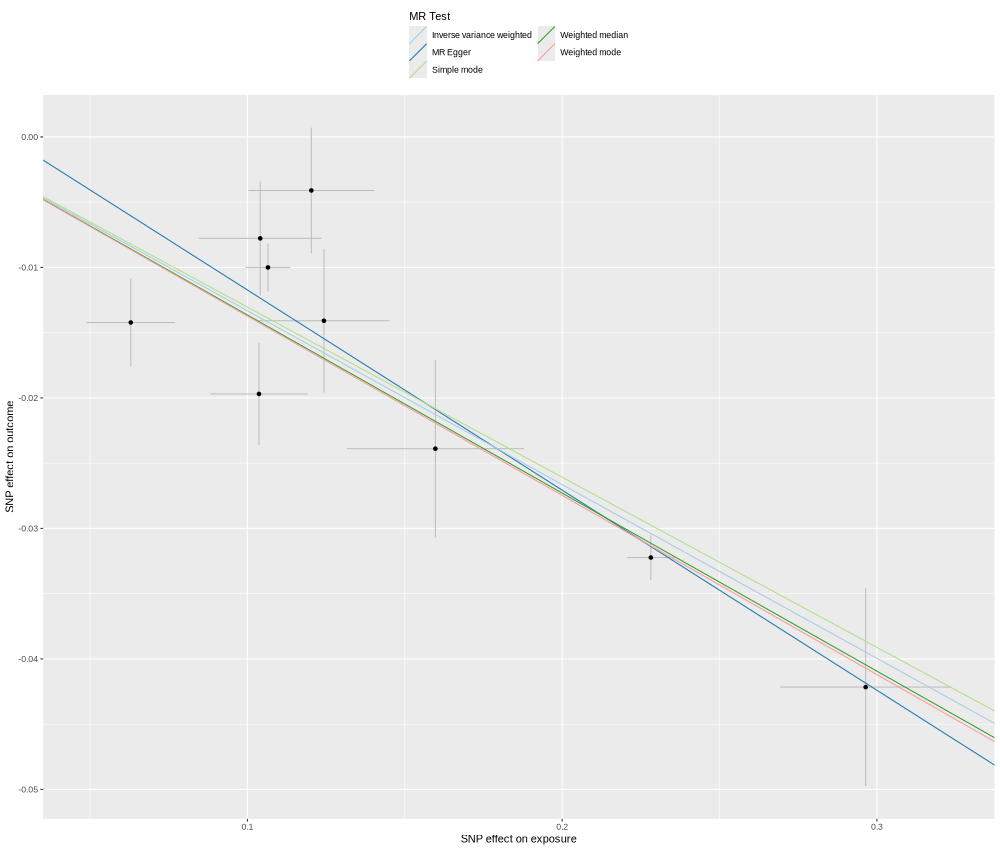

### TIMD4_Q96H15_OID20375_v1_Cardiometabolic_singleSNP.png

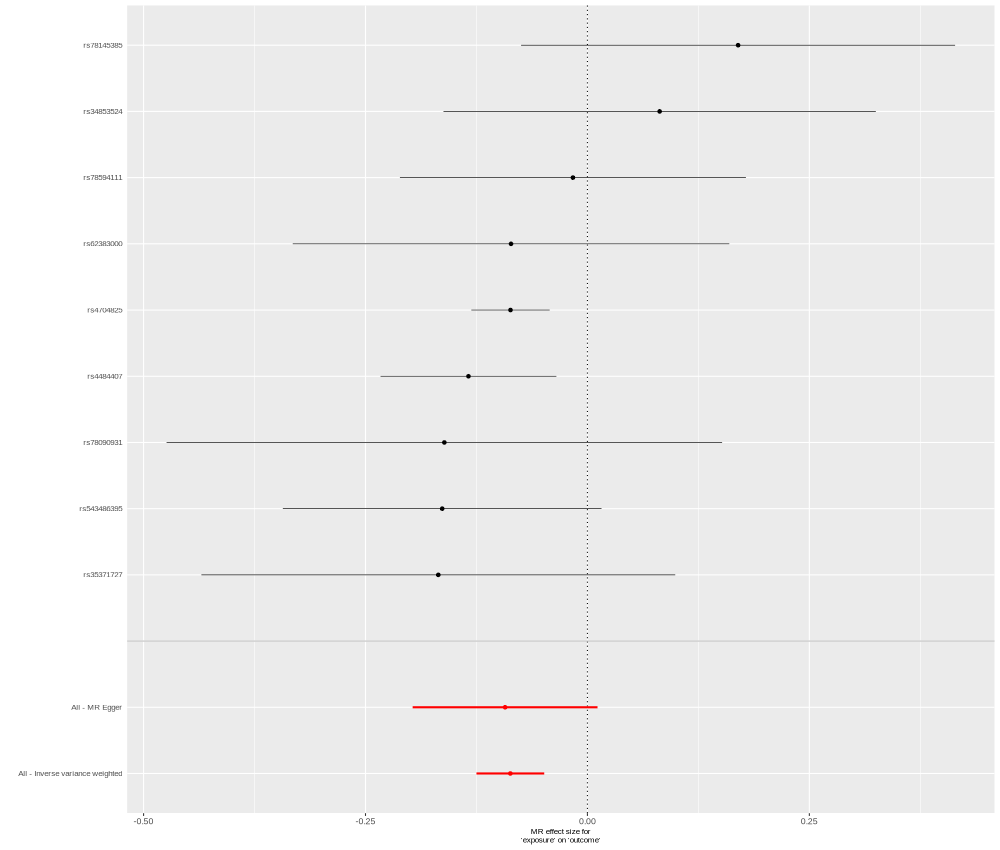

### TIMD4_Q96H15_OID20375_v1_Cardiometabolic_singleSNP.png

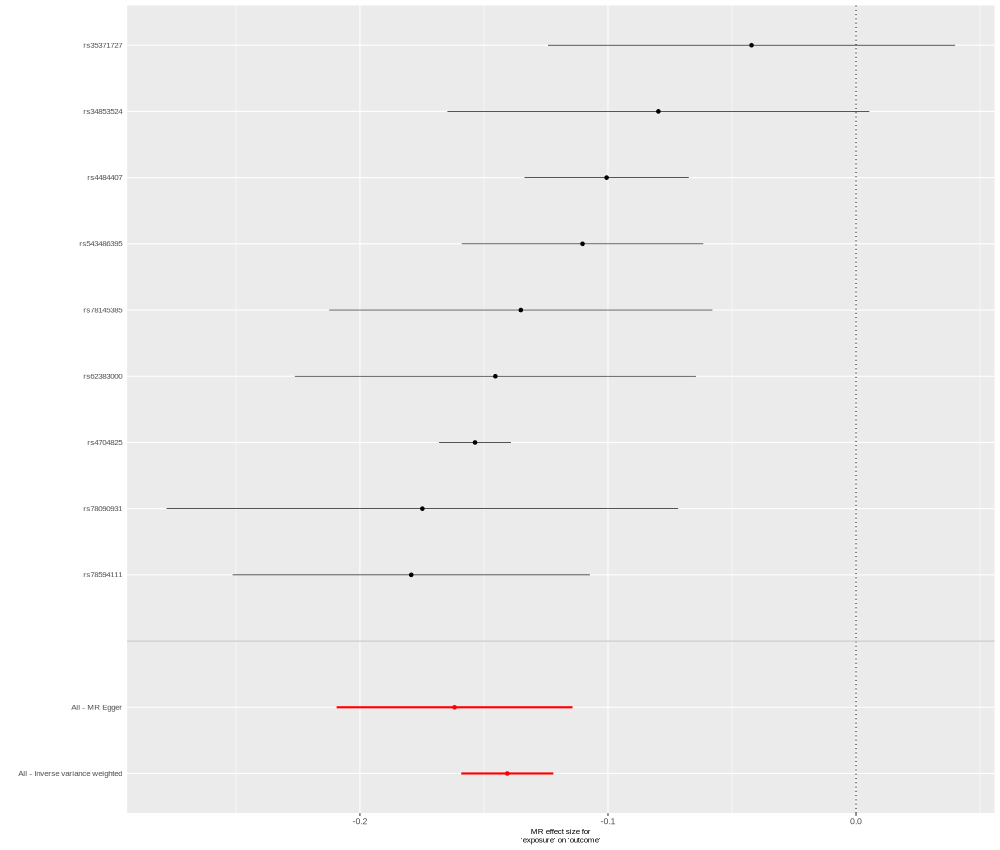

### TNC_P24821_OID20359_v1_Cardiometabolic.png

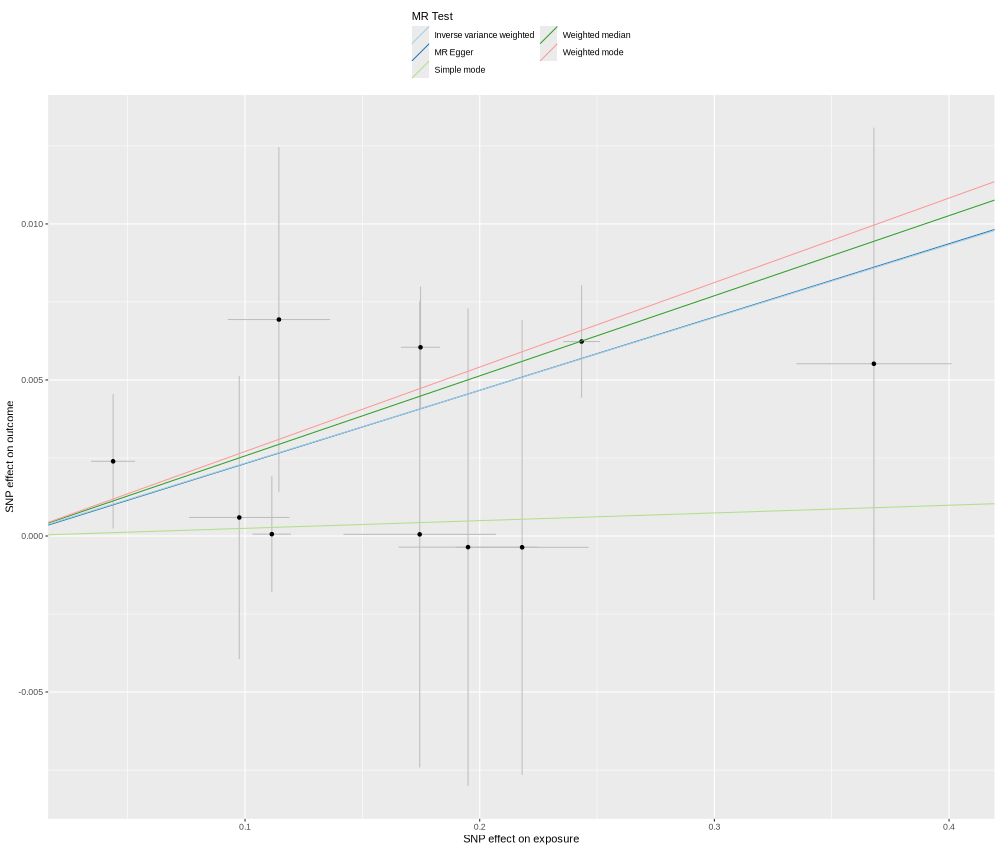

### TNF_P01375_OID20074_v1_Cardiometabolic_singleSNP.png

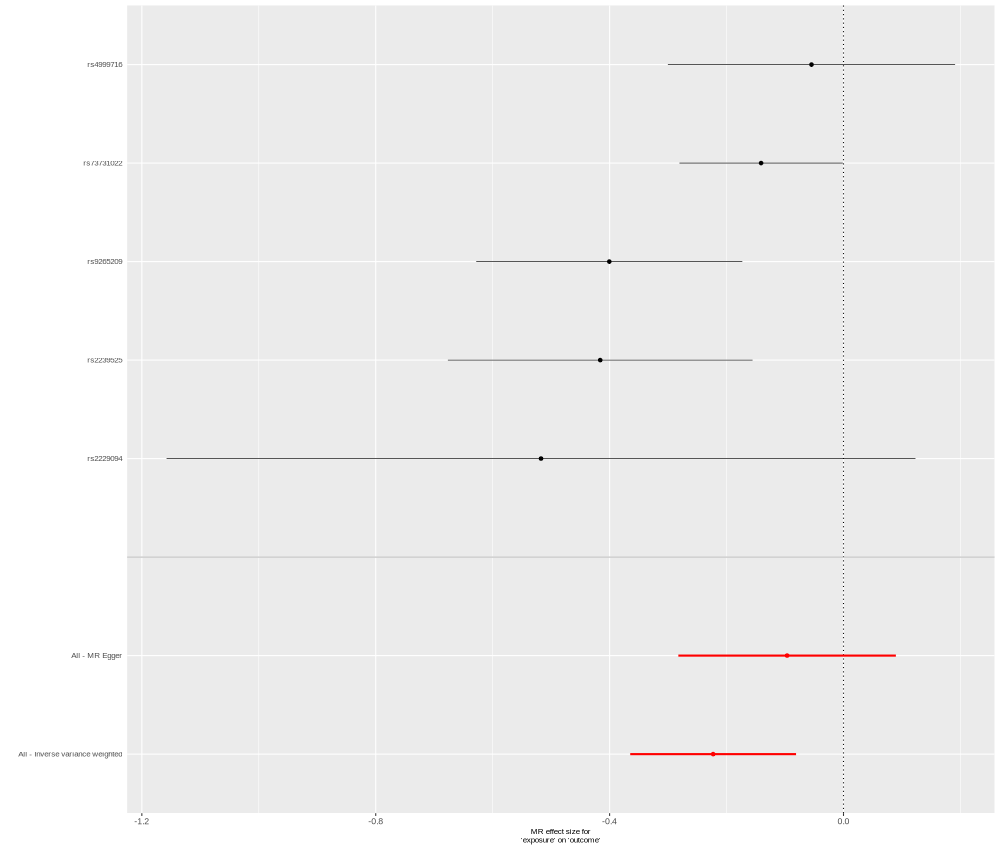

### TNF_P01375_OID20074_v1_Cardiometabolic_singleSNP.png

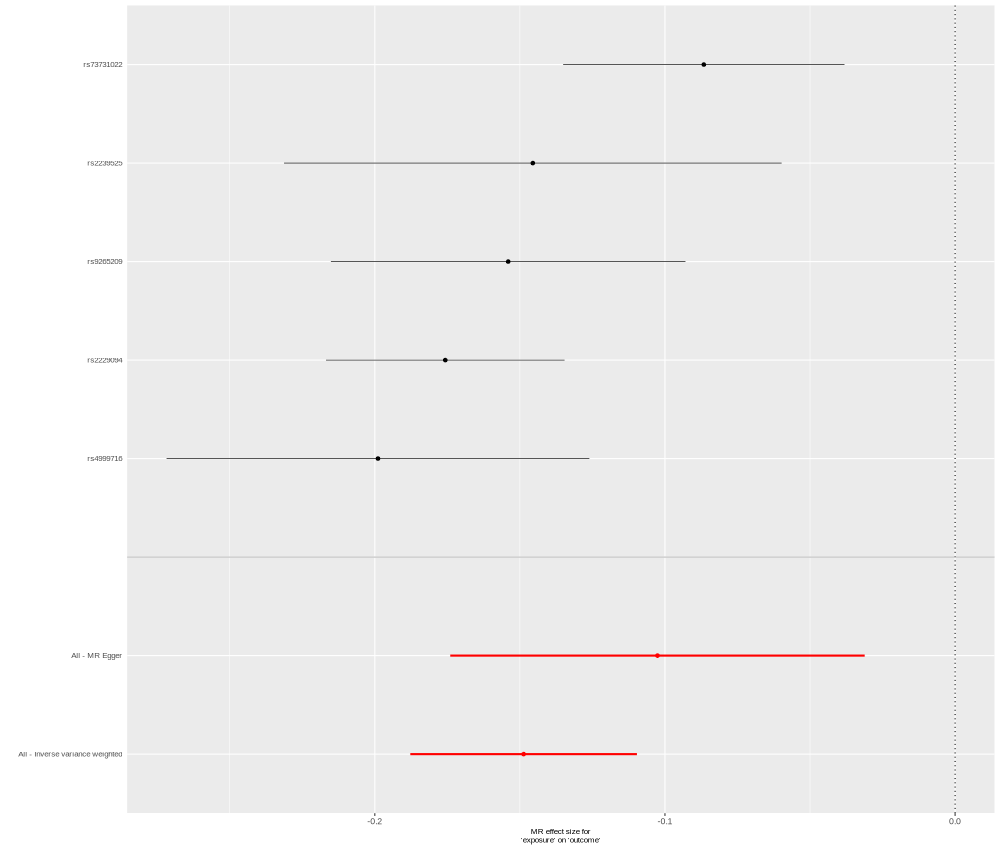

### TNN_Q9UQP3_OID30412_v1_Cardiometabolic_II.png

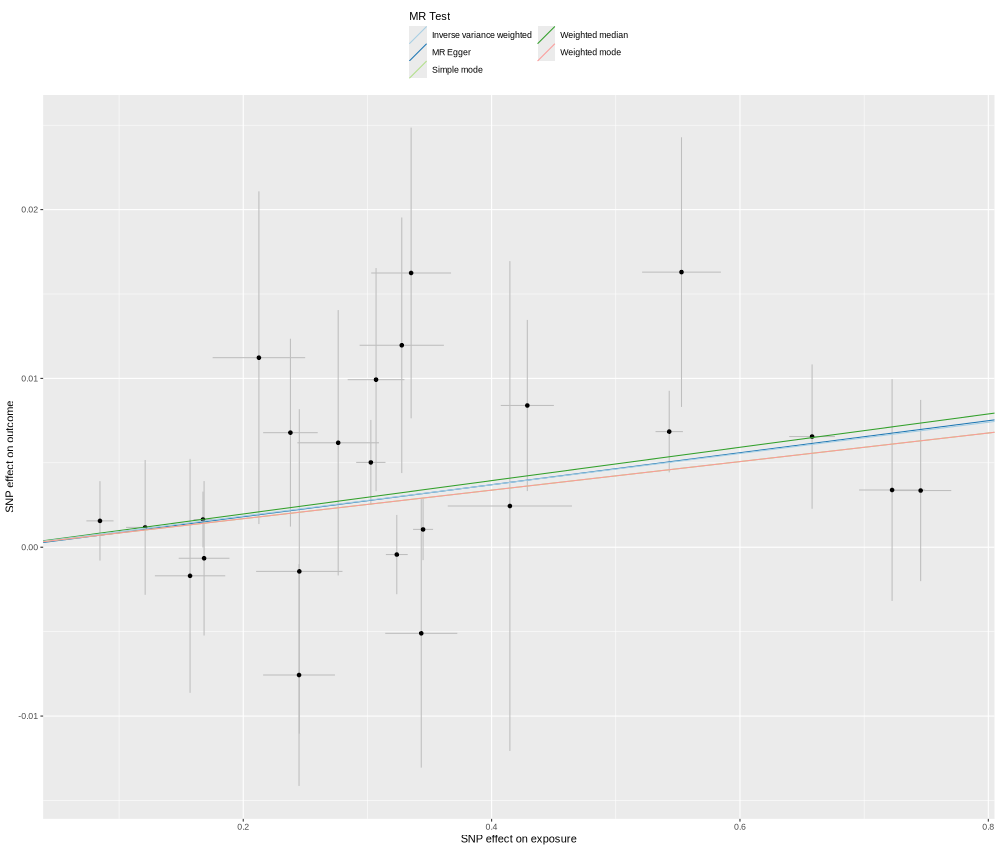

### TRDMT1_O14717_OID31335_v1_Oncology_II.png

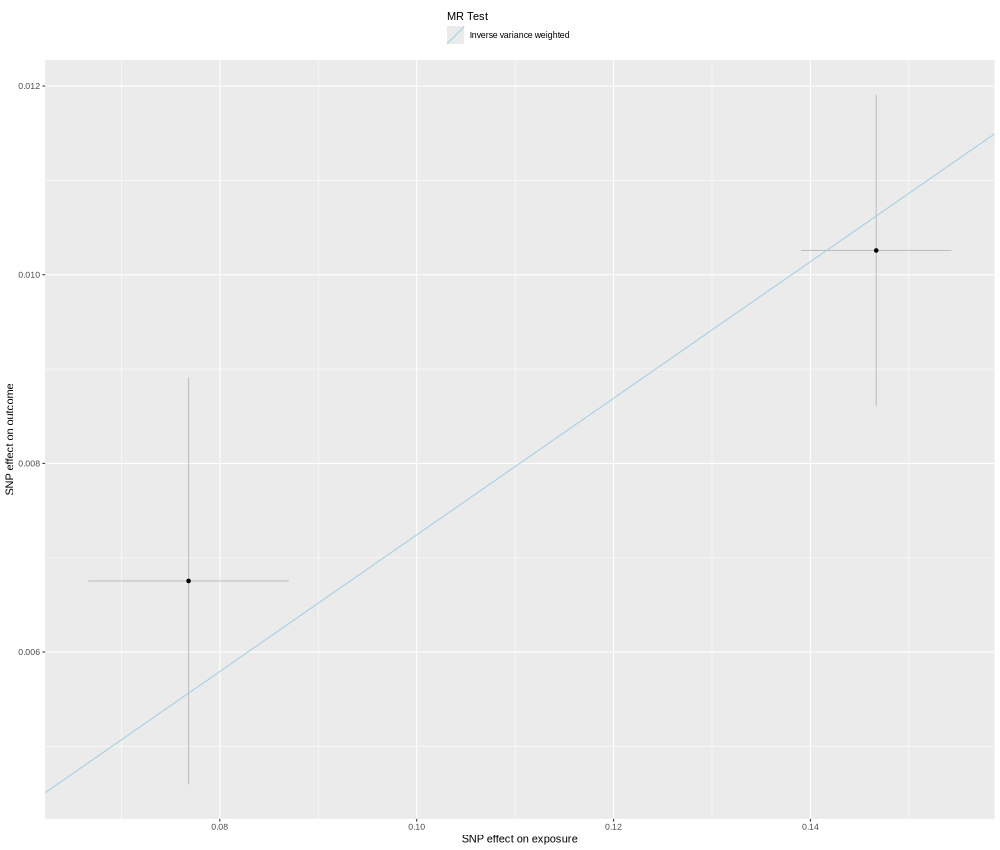

### TXNDC15_Q96J42_OID21495_v1_Oncology_singleSNP.png

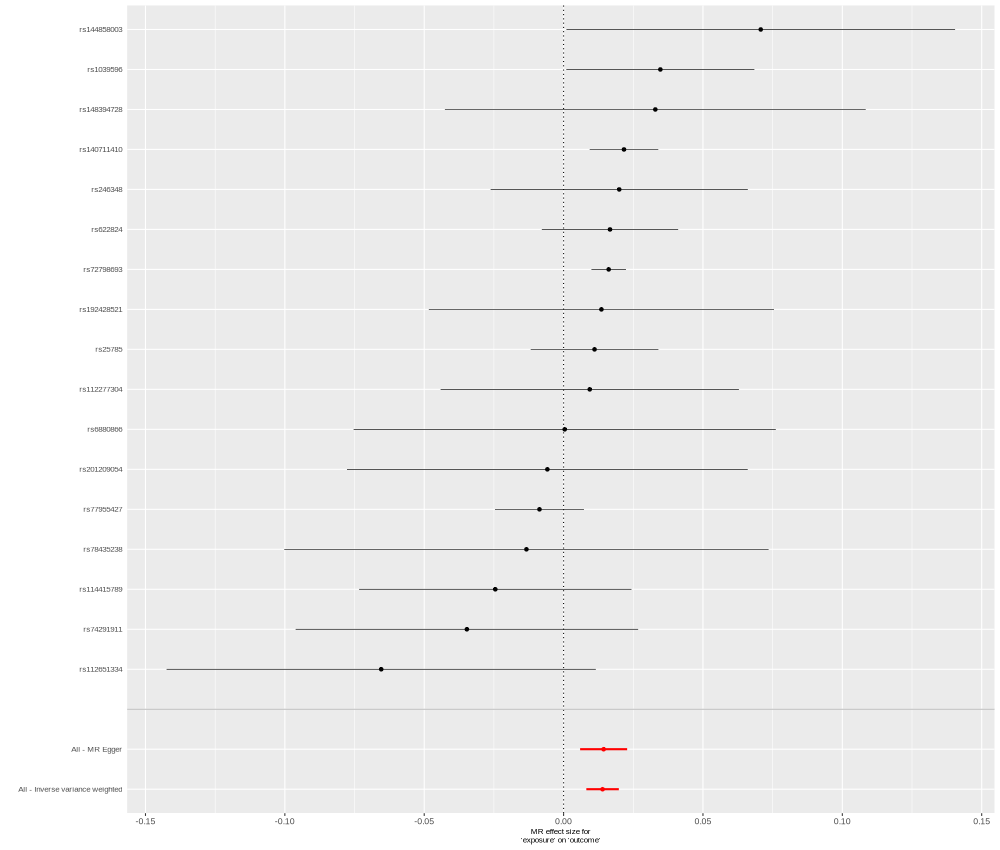

### TYRO3_Q06418_OID20161_v1_Cardiometabolic_singleSNP.png

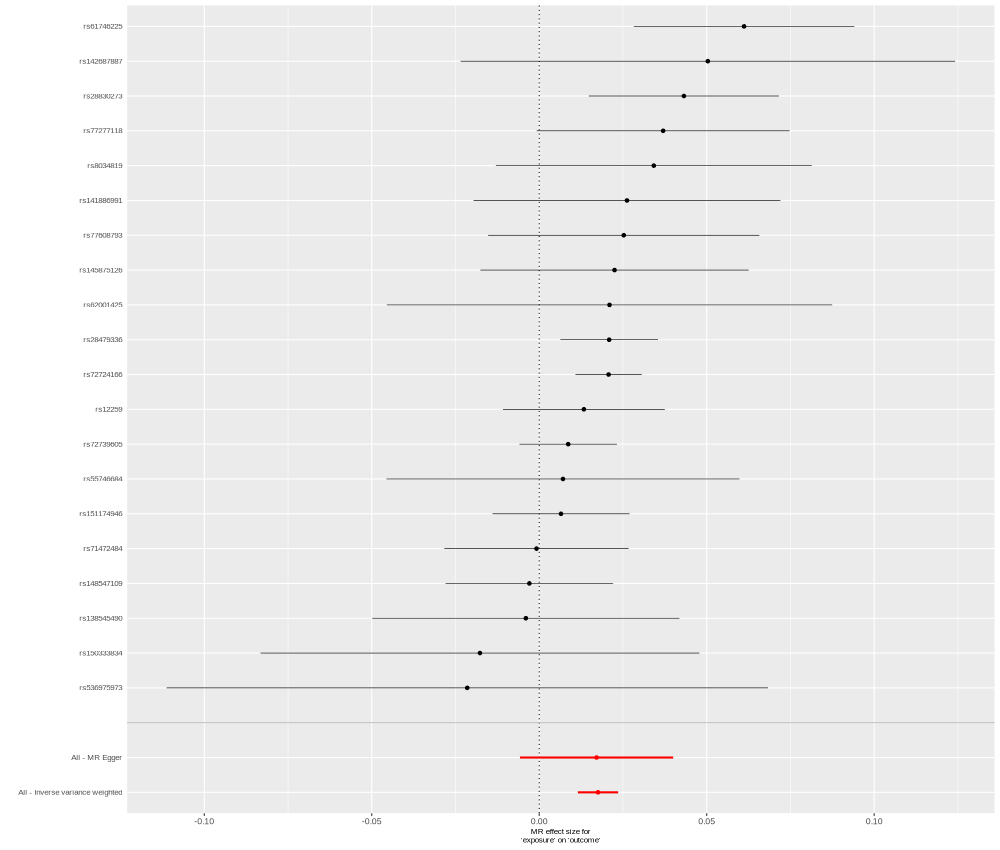

### ULBP2_Q9BZM5_OID20919_v1_Neurology.png

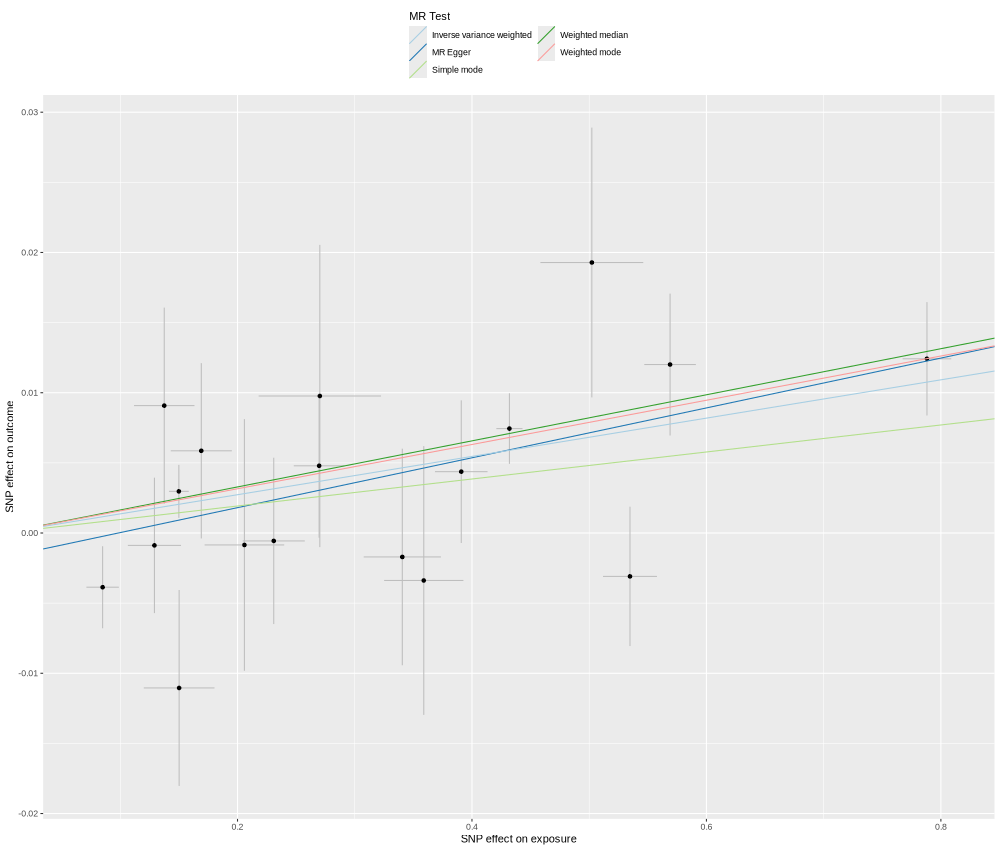

### UROD_P06132_OID30603_v1_Inflammation_II.png

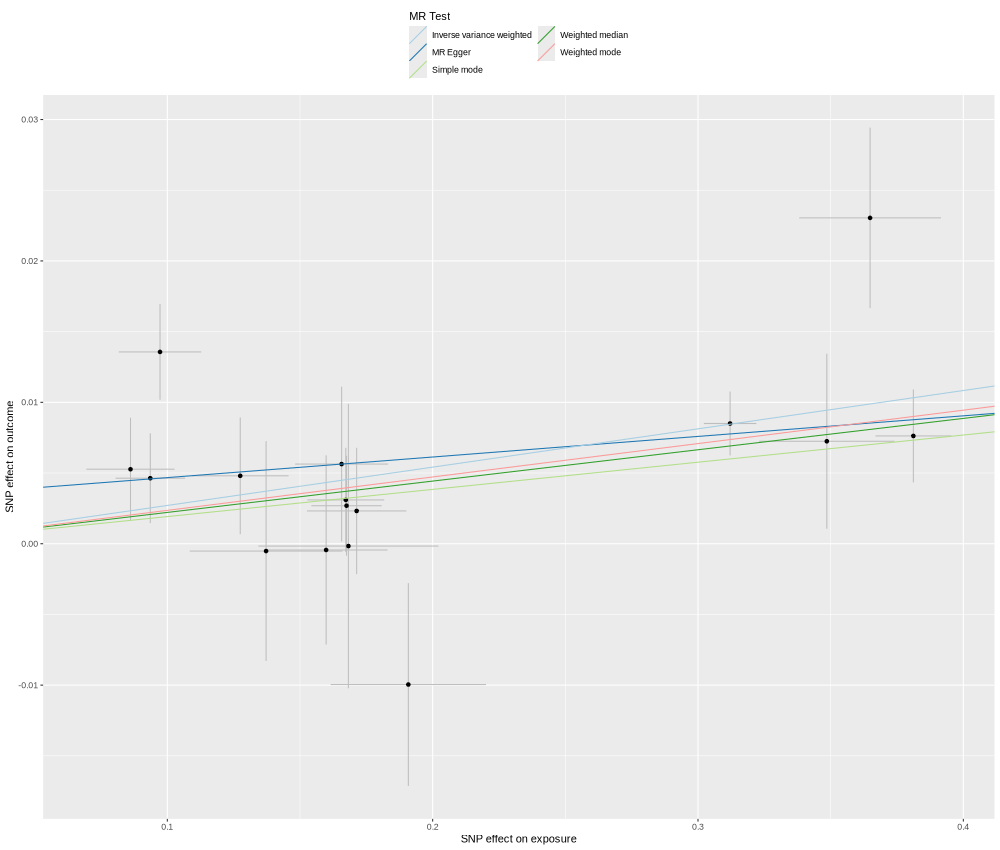

### VMO1_Q7Z5L0_OID21381_v1_Oncology.png

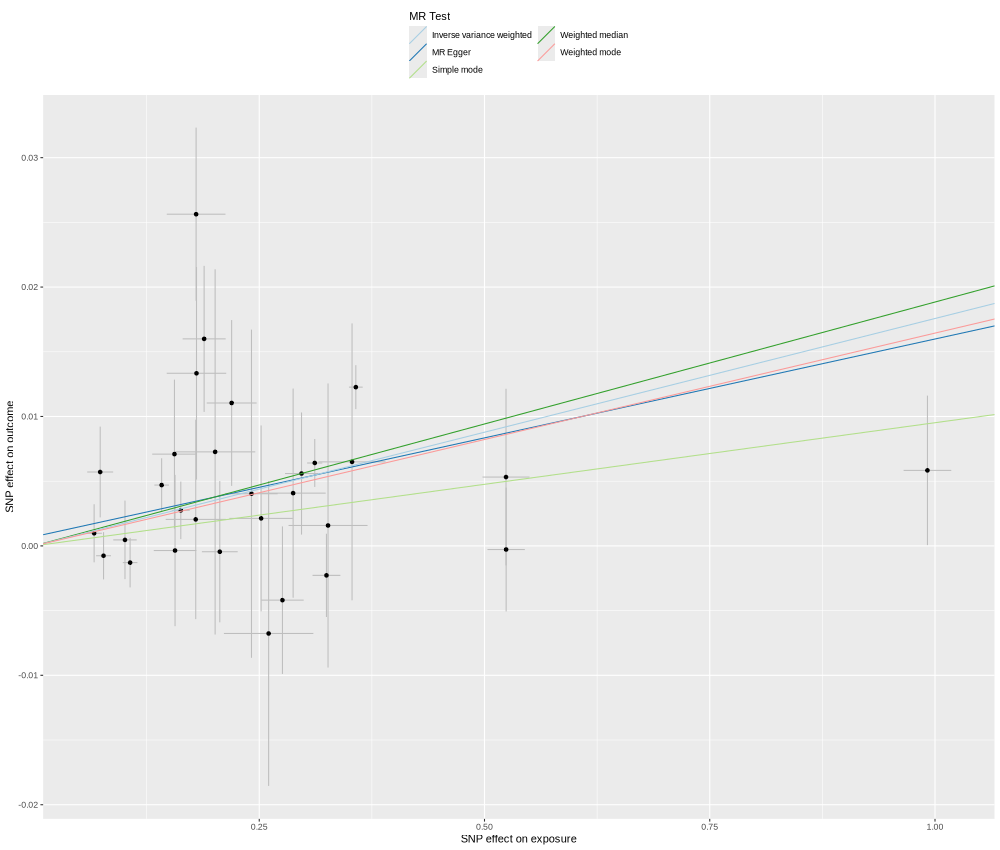

### WFIKKN2_Q8TEU8_OID20785_v1_Inflammation.png

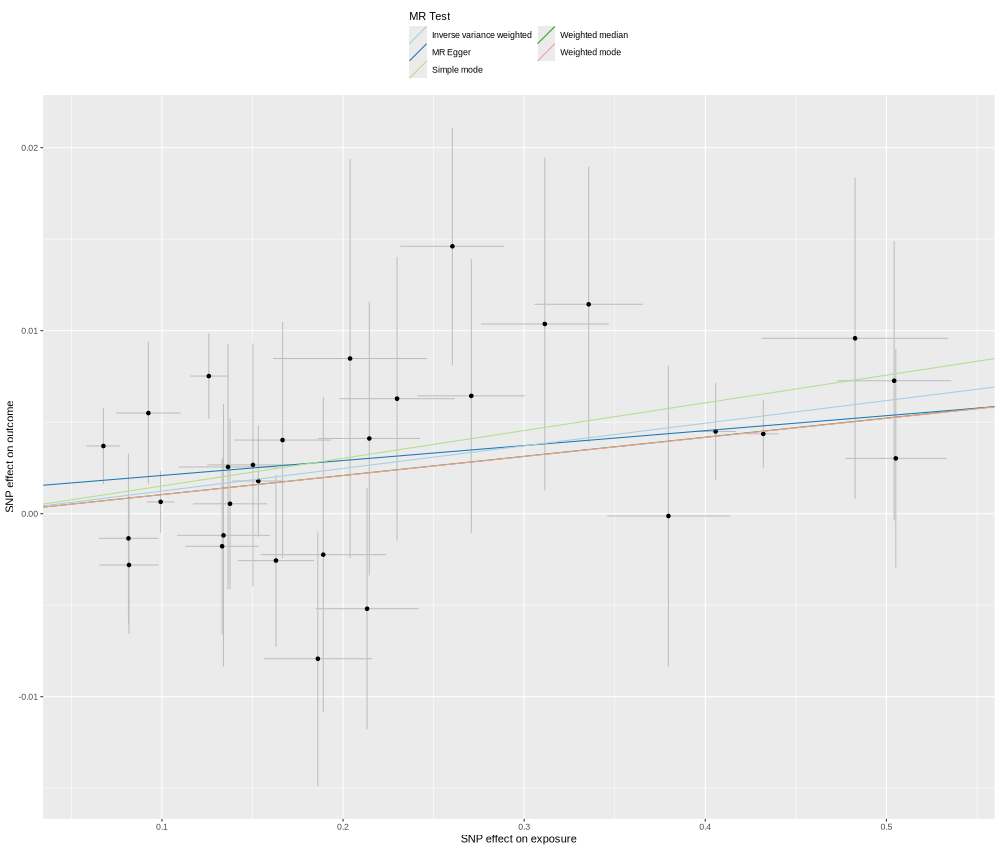
